# Elevated Blood Glucose Levels as a Primary Risk Factor for the Severity of COVID-19

**DOI:** 10.1101/2021.04.29.21256294

**Authors:** Emmanuelle Logette, Charlotte Lorin, Cyrille Favreau, Eugenia Oshurko, Jay S. Coggan, Francesco Casalegno, Mohameth François Sy, Caitlin Monney, Marine Bertschy, Emilie Delattre, Pierre-Alexandre Fonta, Jan Krepl, Stanislav Schmidt, Daniel Keller, Samuel Kerrien, Enrico Scantamburlo, Anna-Kristin Kaufmann, Henry Markram

## Abstract

SARS-CoV-2 started spreading towards the end of 2019 causing COVID-19, a disease that reached pandemic proportions among the human population within months. The reasons for the spectrum of differences in the severity of the disease across the population, and in particular why the disease affects more severely the aging population and those with specific preconditions are unclear. We developed machine learning models to mine 240,000 scientific papers openly accessible in the CORD-19 database, and constructed knowledge graphs to synthesize the extracted information and navigate the collective knowledge in an attempt to search for a potential common underlying reason for disease severity. The literature repeatedly pointed to elevated blood glucose as a key facilitator in the progression of COVID-19. Indeed, when we retraced the steps of the SARS-CoV-2 infection we found evidence linking elevated glucose to each step of the life-cycle of the virus, progression of the disease, and presentation of symptoms. Specifically, elevations of glucose provide ideal conditions for the virus to evade and weaken the first level of the immune defense system in the lungs, gain access to deep alveolar cells, bind to the ACE2 receptor and enter the pulmonary cells, accelerate replication of the virus within cells increasing cell death and inducing an pulmonary inflammatory response, which overwhelms an already weakened innate immune system to trigger an avalanche of systemic infections, inflammation and cell damage, a cytokine storm and thrombotic events. We tested the feasibility of the hypothesis by analyzing data across papers, reconstructing atomistically the virus at the surface of the pulmonary airways, and performing quantitative computational modeling of the effects of glucose levels on the infection process. We conclude that elevation in glucose levels can facilitate the progression of the disease through multiple mechanisms and can explain much of the variance in disease severity seen across the population. The study proposes diagnostic recommendations, new areas of research and potential treatments, and cautions on treatment strategies and critical care conditions that induce elevations in blood glucose levels.

**Highlights:**

- Patients with severe COVID-19 commonly present with elevated blood glucose levels.
- Elevated blood glucose impacts numerous biochemical pathways that can facilitate many steps of the SARS-CoV-2 infection.
- Elevated blood glucose increases glucose in the pulmonary airway surface liquid (ASL), which breaks down the primary innate antiviral defenses of the lungs and facilitates viral infection and replication.
- Elevated blood glucose causes dysregulations of the immune response that facilitates the cytokine storm and acute respiratory distress syndrome (ARDS).
- Elevated glucose levels act synergistically with SARS-CoV-2-dependent inactivation of angiotensin-converting enzyme 2 (ACE2) to escalate the disease to multi-organ failure and thrombotic events.

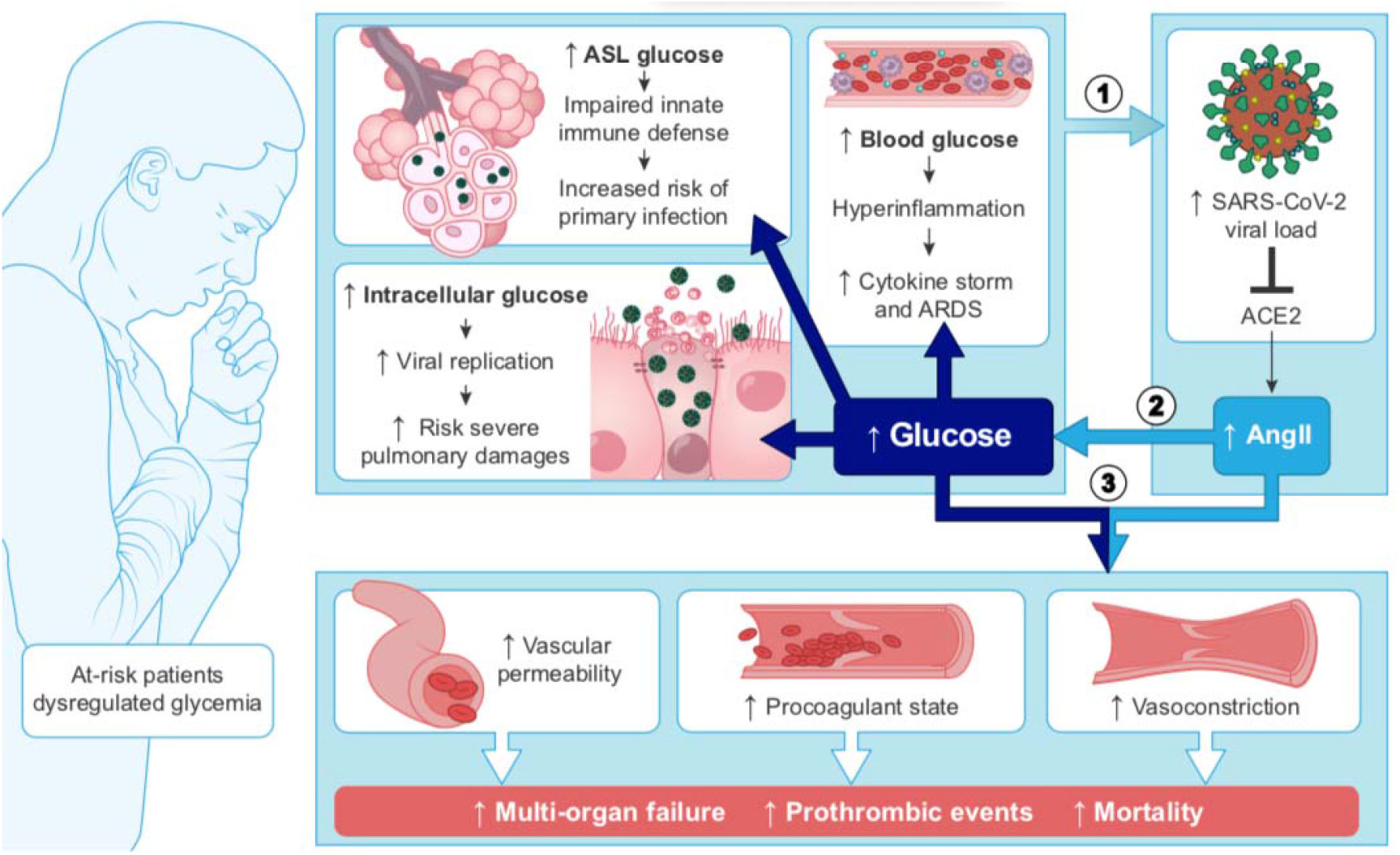

## 1. Introduction

SARS-CoV-2, a novel coronavirus closely related to its predecessor SARS-CoV-1 that was responsible for an outbreak in 2003, emerged towards the end of 2019 in China and reached pandemic proportions, probably within a month (1, 2) causing the disease COVID-19. The actual average mortality rate is lower than the current 2-3% of all confirmed infections because this coronavirus also causes asymptomatic infections in a larger proportion of the population (3, 4). Nevertheless, even with an order of magnitude more asymptomatic than symptomatic infections, this virus would cause over a 100 million hospitalizations and tens of million deaths if allowed to penetrate the world population fully. There are also increasing reports of persistent symptoms and various long-term sequelae from COVID-19 (5–8), warning of an even deeper health crisis. Containment strategies, and lockdown when these fail, slow down full penetration of the world’s population allowing nations time to prepare a public health strategy, improve treatments and develop vaccines. Vaccinating a major fraction of the world population could eradicate the virus, but this would need to be performed in a globally coordinated manner and fast enough to drastically cut transmission rates before the virus can mutate sufficiently to necessitate restarting a global vaccination program with a new vaccine. This is a major challenge since the current rate of infections is still in the hundreds of thousands per day, which provide ideal conditions for the virus to mutate. The disease has thus become endemic in the world and will most likely remain a health crisis for many years to come. It is thus of paramount importance to gain deep insight into the factors responsible for the progression of the disease, to improve disease management, and to develop new treatment strategies.

The main symptoms of COVID-19 are fever, cough, fatigue, dyspnea, myalgia, and chest pain, with diarrhea included among the less common symptoms (9–13). In addition, anosmia and a loss of taste are other early and long-lasting symptoms (14, 15). In 70-80% of known cases patients present with mild to moderate symptoms and the disease is manageable without hospitalization, with patients recovering within a few days or weeks. However, in about 15% of known infections the disease progresses to a severe form, with pneumonia as the primary complication often requiring hospitalization. Lung capacity decreases significantly and blood oxygen levels drop dangerously low, requiring nasal oxygen, and in more severe cases, intubation using mechanical ventilators. In 4-7% of known cases the disease becomes life threatening, requiring intensive care (16), with acute respiratory failure in around 20% of these cases (17).

The substantial amount of patient data that has become available has allowed the identification of groups of people at higher risk of the disease progressing to a severe form and with a higher mortality rate. Of all COVID-19 deaths, more than 50% are patients over 80 years old (Supplementary Figure 1A). Indeed, the case fatality rate (CFR; the percentage of deaths among positively diagnosed infections) increases sharply with age: from <1% below the age of 50 years, to 2-3% around 60 years, and as much as 10-20% above the age of 80 (Supplementary Figure 1B). The main risk factors that add to this age-related CFR include hypertension, cardiovascular diseases, diabetes mellitus (DM) and severe obesity (18–24), with varying impact depending on the country (25). The precedent SARS-CoV-1 showed a similar clinical profile and also affected more severely the elderly and those with diabetes and hypertension (26–28). In fact, the mortality rate (MR; the percentage of deaths among all people) increases with age for many other diseases as well, and diabetics and patients with hypertension or cardiovascular disease are also more susceptible to succumbing to a range of diseases (29, 30), including even seasonal influenza infections (31, 32).

A puzzling aspect of COVID-19 is why the disease becomes so severe with age and preconditions, and in some apparently healthy or young patients. Most of these critical cases seem to be associated with a “cytokine storm” in the lungs (33, 34), an exaggerated immune response that produces high levels of cytokines that damages the airway epithelium, leading to acute respiratory distress syndrome (ARDS), requiring ventilation or intensive care with intubation, which is fatal in 20%-50% of cases (24, 35–38). Survivors of the cases that require invasive ventilation also need long-term rehabilitation (39). The 20%-50% deaths in intensive care units (ICU) is due to respiratory failure, multi-organ failure and/or septic shock (40, 41). It has furthermore emerged that the virus affects blood coagulation, leading to micro- or macrovascular thromboses often associated with acute pulmonary embolism and cardiac injury (42–46). Data from China indicates that 53% of deaths were related to respiratory failure, 7% to septic shock, 33% to both and 7% to unclear mechanisms (39). Importantly, disseminated intravascular coagulation (DIC) is observed in 71% of fatalities, but only in 0.6% of surviving patients (47).

Several biomarkers predict a poor outcome of the disease, including increased levels of IL-6 (interleukin 6), serum ferritin, CRP (C-reactive protein), LDH (lactate dehydrogenase), D-dimer and fibrinogen (11, 12, 48–50), as well as reduced levels of antithrombin (45) and lymphopenia (51). Fasting plasma glucose (FPG) level at admission has more recently emerged as an additional strong risk factor for COVID-19 mortality (52–56). However, there is still no definitive treatment strategy or biomarker(s) that can help reduce the mortality rate sufficiently to resolve the health crisis. Treatments with many drugs such as lopinavir-ritonavir, interferon, hydroxychloroquine, remdesivir or corticosteroids were investigated (57) however have shown no or controversial efficacy (58, 59). Other treatments that try to dampen down the cytokine storm (such as anti-IL-6 antibody tocilizumab (60) or the anti-IL-1 anakinra (61) among others (62–64) were also investigated with variable success. Thus far, only the use of anticoagulants seems to consistently improve the outcome for patients (65, 66).

This pandemic has accelerated the development of a large number of vaccines on an unprecedented timeline (67–69). Several vaccines, based on different strategies (vector-based, mRNA-based or protein-based) and delivery systems (lipid nanoparticles, attenuated viruses) with proven efficacy and safety, are now available (70–74). The vaccination campaign has started in many countries, but the time required to get enough people vaccinated worldwide to eradicate the virus, is still too slow to stop the spread of the virus. The majority of the world population needs to be vaccinated to sufficiently reduce global transmissions and lower the risk of new variants emerging and having to restart the global vaccination campaign with a new vaccine. With global travel mobility, virus variants may require vaccination boosters or complete restarts in nations previously fully vaccinated. Other uncertainties include the period of immunity and efficacy of the vaccines in the various groups at risk (75) and hence, investigations into the pathophysiology of SARS-CoV-2 and new treatments must continue in parallel and with urgency.

Understanding why some groups are naturally protected while others are vulnerable (76) may improve management of this disease. All the known preconditions (i.e., aging, DM, obesity, hypertension) are commonly accepted to be associated with chronic inflammation and a weaker immune system, which could explain the higher sensitivity and complications of the disease (77–81). Another association with severe cases that is emerging is hyperglycemia (54, 56, 82, 83), and it is now well accepted that a tight control of glucose levels is important in the management of COVID-19, not only in diabetic patients (84–88) but also in general (89). However, the role that glucose plays in the progression of the disease and the importance of managing glucose levels in the aging population, in diabetic people and in apparently healthy groups, is unclear.

In 2020, the White House launched the CORD-19 database (COVID-19 open Research Dataset), a dataset of full text articles on COVID-19, SARS-CoV-2, and related coronaviruses that has been made open access to facilitate global collaboration in understanding and management of the pandemic and to accelerate development of treatments. Since it is humanly impossible for any researcher to read all these papers, let alone synthesize all the results, findings and knowledge, we developed natural language and machine learning tools to automatically mine the contents and constructed a knowledge graph to synthesize the data and navigate the knowledge. We found strong support for blood glucose as a fundamental risk factor that could explain most of the variance in the severity of the disease across age groups and groups with the identified preconditions. The knowledge graph allowed us to navigate deep into numerous biochemical, homeostatic and metabolic mechanisms of action of glucose in the context of this disease and find papers that implicate glucose at virtually every step of the SARS-CoV-2 infection.

The literature shows that elevated blood glucose impairs the first level of innate immune defense in the lung and creates ideal conditions for the virus to access, enter and replicate in target cells. It also shows that elevated glucose facilitates the development of multiple complications of the disease such as the hyperinflammation and pro-coagulation. A case for impaired glucose metabolism as the primary pre-condition for the severity of COVID-19 becomes even more compelling when the data published in the CORD-19 dataset is combined with established knowledge of glucose biochemistry, metabolism, and homeostasis, and with the role of glucose in related pathologies.

A clinical hypothesis must be tested in well controlled clinical trials before it can be used to take any medical actions. If clinically validated, the hypothesis of reduced glucose metabolic capacity as a pre-condition underlying age dependency and other pre-conditions of disease severity, and induced elevations of glucose as favoring disease progression, would have implications for diagnostic measures during admission. These include measurement of postprandial glucose (PPG) combined with HbA1c (glycated hemoglobin A1c) and not only FPG measurements. In addition, other measurements of insulin metabolism that not only aim at detecting diabetes, but any possible dysregulation of glucose metabolism such occurring in pre-diabetes, acute hyperglycemia, impaired glucose tolerance (IGT), or stress induced hyperglycemia would be indicated. This hypothesis also has implications for disease management where assisted control of blood glucose levels during hospitalization, prevention of hyperglycemia during critical care, and avoiding high levels of intravenous glucose in ICU becomes important. Glucose tolerance screening of those not yet infected by the virus could predict those groups with the highest risk of severe disease and enable improved prioritization for vaccinations.

## 2 Results

### 2.1 Analysis of the CORD-19 dataset

For our analysis, we used the CORD-19v47 dataset that contained, at the time of the study, over 240,000 scientific articles (see Methods). Given that it is humanly impossible to read this number of papers, we developed machine learning models to extract the most frequent entities mentioned in the context of respiratory viral infections, coronaviruses in general, and SARS-CoV-2 in particular. We then constructed a knowledge graph of these entities to synthesize the data and navigate the subset of knowledge that specifically relates to a potential role of glucose in the progression of COVID-19 (Figure 1).

**Figure 1:**
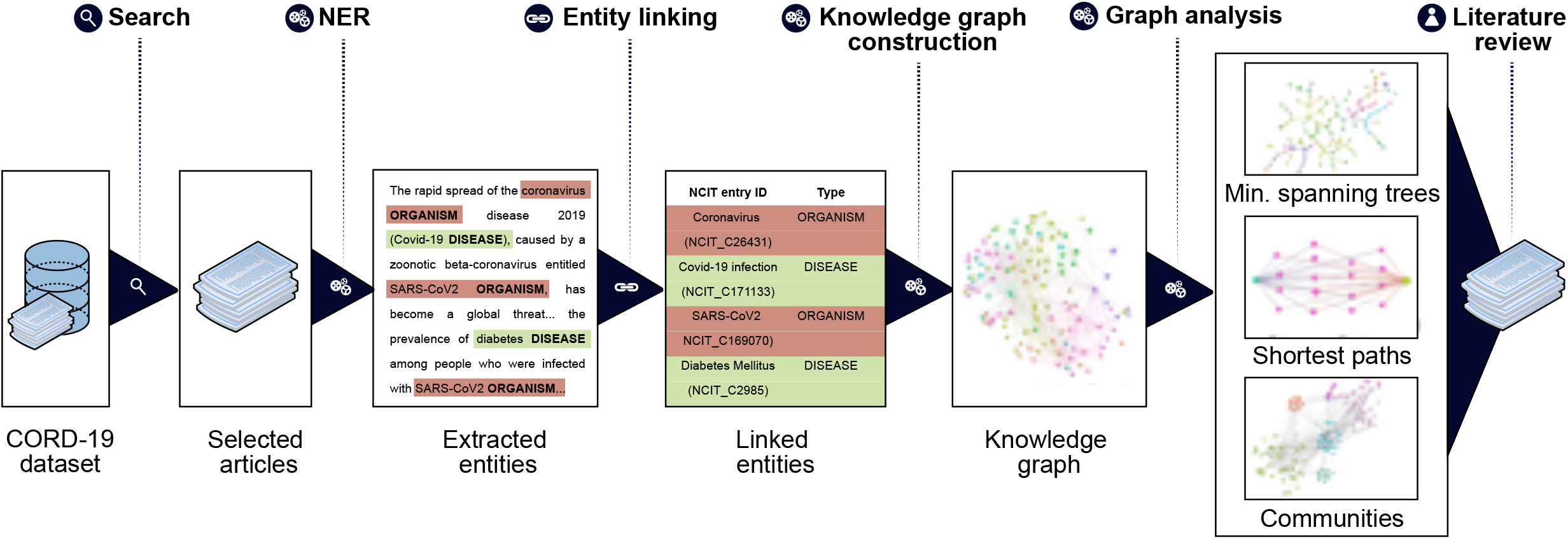
Knowledge graph construction and analysis pipeline. Entities of interest are extracted using the named entities recognition (NER) techniques on the entire CORD-19 dataset, or on a subset of papers selected for matching user-specified query (Search). The extracted entities are then linked to the NCIT ontology terms and passed to the knowledge graph construction stage. On this stage we build a knowledge graph using extracted and linked terms as nodes and their co-occurrences as edges. Finally, we perform graph analytics that allows us to navigate the knowledge graph and reveal connectivity patterns and subgraphs carrying the most important term relations (Minimum spanning trees, BMIPs, communities). Data from the graphs are then used to guide further literature review (see Methods for details).

We began by extracting entities using named entity recognition (NER) models trained to recognize nine selected entity types (see Methods, Entities Extraction). Each extracted entity was mapped to a term in the National Cancer Institute Thesaurus (NCIt) ontology allowing to resolve most of the ambiguities of lexical variations as well as synonyms, aliases and acronyms (see Methods, Entity linking). Linking to the NCIt ontology also enabled access to standardized semantics of the entities, their human-readable definitions, and their hierarchical structure within the ontology. This approach yielded over 400,000 unique and relevant entities.

Next, we constructed a knowledge graph by creating a node for each extracted entity and building a link when entities were co-mentioned. The importance of a node was computed as the weighted degree centrality and the strength of a link was computed using mutual information techniques (see Methods). In this context, the weighted degree centrality can be interpreted as the relative importance of the entity in the dataset, an edge between a pair of entities as the presence of some association between them, and the corresponding edge weight as a quantification of the strength of the association. The links do not on their own represent the type of association, which rather emerges from the overall structure of the graph. The network was then partitioned into the nine entity types to obtain a first high level view of the contents of the CORD-19 dataset (Figure 2). The entity types *protein and symptom/disease* are the most represented entities in the CORD-19 dataset (27% and 21% respectively), whereas c*ell compartment* is the least common. The six remaining entity types are roughly equally represented (between 6% and 11%). This rather trivial analysis does provide a first high-level view of the distribution of different entity types found in the dataset.

**Figure 2:**
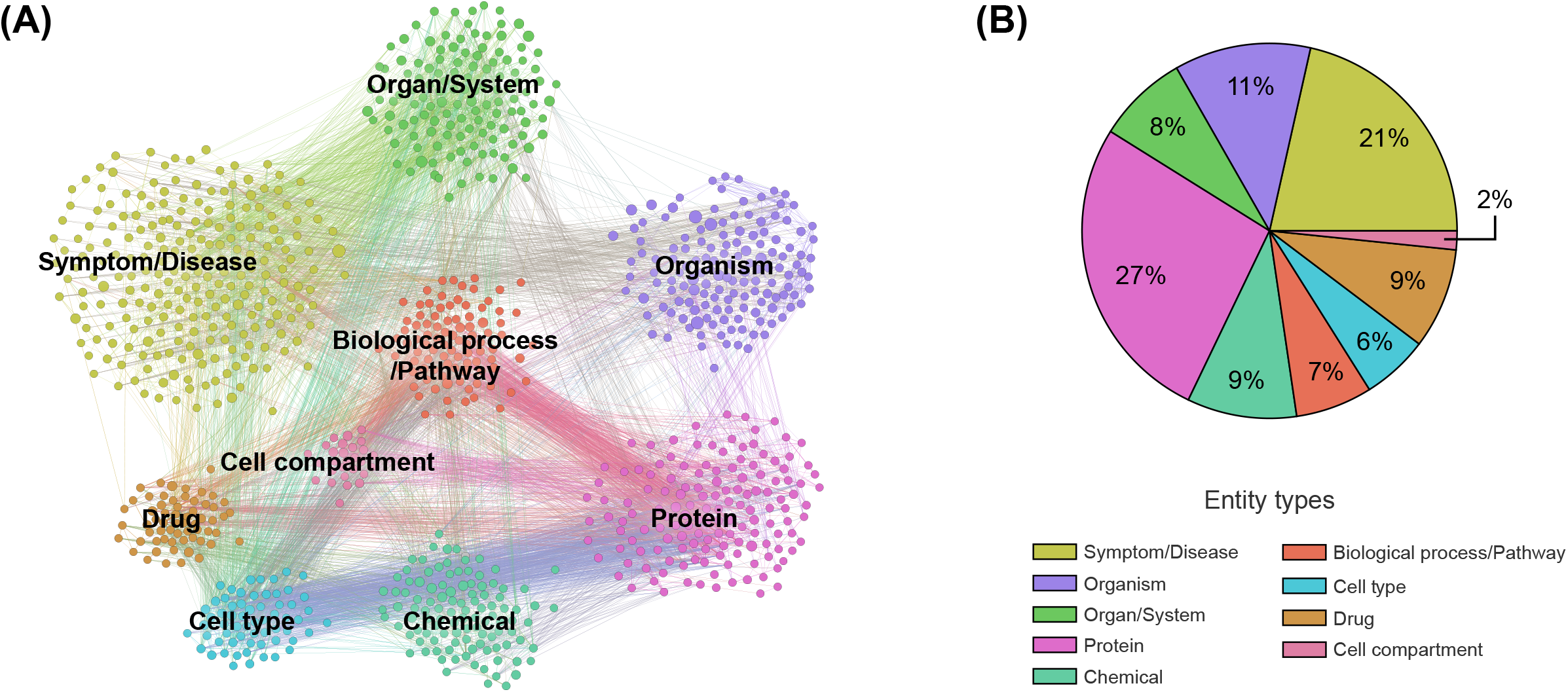
Overview of co-mention graph of high-level entities. (**A)** Sample of a knowledge graph containing ∼1,000 nodes representing the most frequent high-level entities and those with edges with the highest mutual information (see Methods). (**B)** Distribution of the entity types detected. Different entity types are colored according to the legend. A zoom into the co-mention subgraphs of each entity type is available in Supplementary Figure 2.

To validate that the associations between entities are semantically meaningful (as opposed to incidental), we applied community detection methods to objectively partition the knowledge graph into clusters of strongly associated entities (see Methods, Community detection). The emergent communities that were automatically detected, revealed five different conceptually coherent topics (biology of viruses, diseases and symptoms, immune response, infectious disorders, and chemical compounds) supporting some degree of relevance of the associations (Supplementary Figure 3).

#### 2.1.1 Presence of the entity *glucose* in the CORD-19 database

To obtain a next deeper level view of the contents of the dataset, we measured the frequency of entity mentions in each paper. COVID-19 is indeed the most frequently mentioned entity providing a minimal validation of the automatic entity extraction by the ML models (Table 1). The entity *glucose* is found in 6,326 of the 240,000 papers, making it the 179^th^ most frequently mentioned entity among more than 400,000 entities extracted. It is also the 17^th^ most frequently mentioned entity in the entity type *chemical* (over >20,000 chemical entities extracted) (Supplementary Table 2B), indicating the extent to which glucose is present in the CORD-19 database. Of these chemicals, the entity *glucose* in the one biochemical with the deepest and broadest association with all stages of the virus infection (see below).

**Table 1:** Entity Ranking. The entity *glucose* (the term glucose and any semantics of the term) rank (179^th^) mentions in the CORD-19v47 database following COVID-19-related entity-types recognition. The list of the 100 most frequent entities is available in Supplementary Table 2A.

| Rank | Entity | Frequency | Entity type | Rank | Entity | Frequency | Entity type |
| --- | --- | --- | --- | --- | --- | --- | --- |
| 1 | COVID-19 | 110,145 | Symptom / Disease | 174 | spike protein | 6,438 | Protein |
| 2 | virus | 75,012 | Organism | 175 | fatigue | 6,397 | Symptom / Disease |
| 3 | infectious disorder | 73,574 | Symptom / Disease | 176 | asthma | 6,370 | Symptom / Disease |
| 4 | coronavirus | 67,945 | Organism | 177 | CO2 | 6,356 | Chemical |
| 5 | human | 61,816 | Organism | 178 | hepatitis virus | 6,343 | Organism |
| 6 | viral | 52,801 | Organism | 179 | glucose | 6,326 | Chemical |
| 7 | SARS-COV-2 | 49,386 | Organism | 180 | immunocompromised | 6,310 | Symptom / Disease |
| 8 | SARS coronavirus | 41,463 | Organism | 181 | vomiting | 6,275 | Symptom / Disease |
| 9 | blood | 33,970 | Organ / System | 182 | erythrocyte | 6,254 | Cell Type |
| 10 | person | 33,893 | Organism | 183 | pain | 6,233 | Symptom / Disease |
| ... | ... | ... | ... | ... | ... | ... | ... |

#### 2.1.2 Knowledge graph of “glucose in coronaviruses infection”

In order to obtain the context in which glucose is mentioned in the dataset, we performed a mutual information-guided search of the paths that are formed by the links of the knowledge graph from “glucose” to “SARS-CoV-2”. Since there are a large number of potential paths between any two entities, we filtered them by the best mutual information pathways approach (see Methods “BMIPs Search”), and then aggregated the entities according to their BMIP. Examination of the clusters revealed five coherent coronavirus-specific topics (comorbidities in high-risk group, SARS-related symptoms and complications, SARS-related drugs, SARS disease biomarkers and inflammation, and coronavirus receptors and RAAS (renin–angiotensin–aldosterone system), see Figure 3), showing that *glucose* is mentioned in the context of numerous stages of the coronavirus infection: from high-risk groups through to disease development and complications. In addition, three entities directly associated with glucose (*glucose transport*, *glucose uptake* and *glucose tolerance test*) were found in the BMIPs (Figure 3, red nodes), indicating that glucose is also mentioned in the context of glucose metabolism.

**Figure 3:**
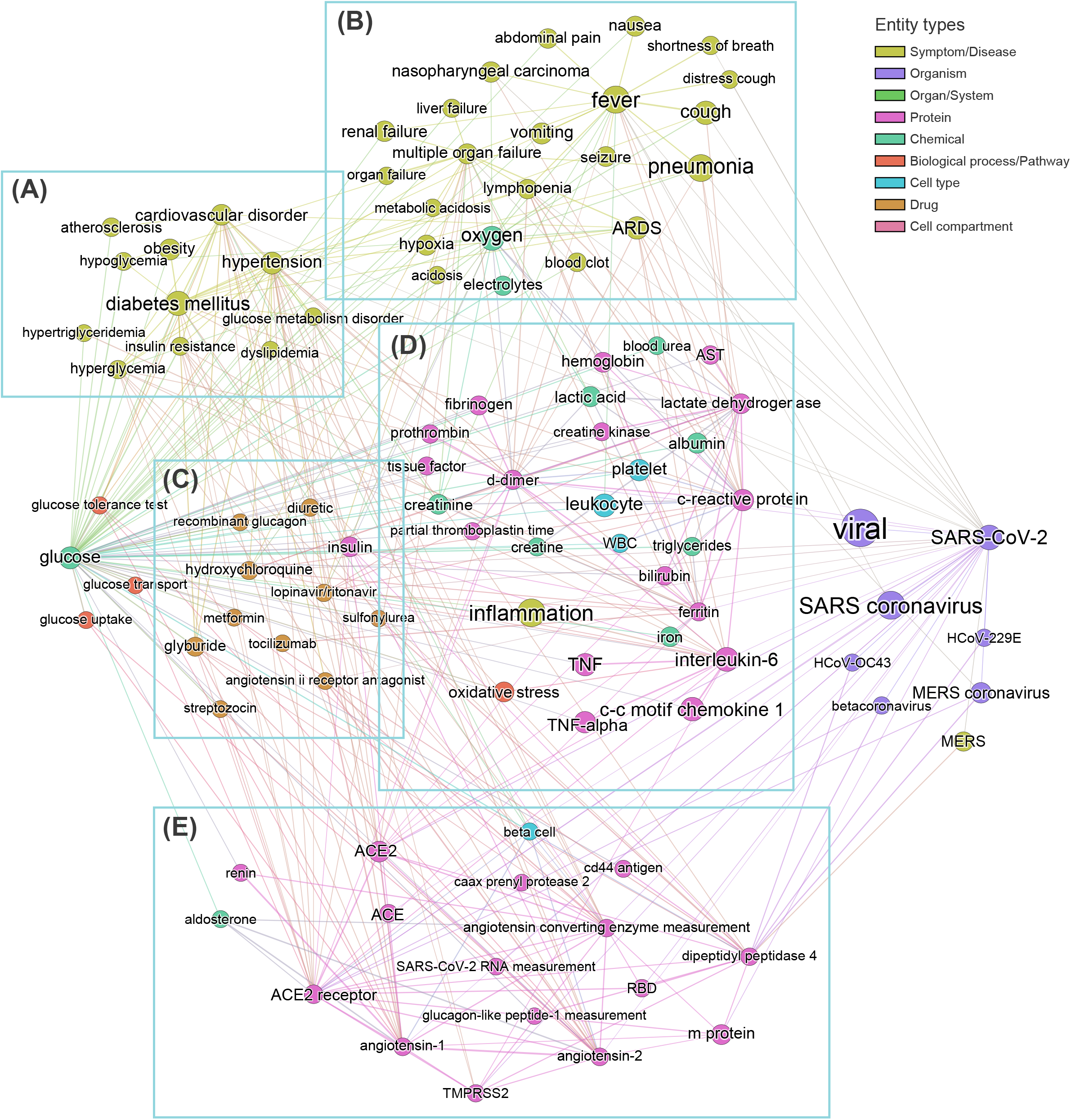
Subgraph obtained by aggregating the 20 Best Mutual Information Pathways from “glucose” to “SARS-CoV-2”. Node sizes are proportional to the weighted degree of nodes and node colors represent different entity types. Analysis of entities encountered during the mutual information guided shortest path search (see Methods) from ‘glucose’ to ‘SARS-COV-2’ allows us to identify five groups of entities, each related to a specific field in coronavirus infection: (**A**) comorbidities in high-risk group, (**B**) SARS-related symptoms and complications, (**C**) SARS-related drugs, (**D**) SARS biomarkers and inflammation, **(E)** coronavirus receptors and RAAS system. No entities from c*ell compartment* or *organ/system* entity types were detected in this analysis.

#### 2.1.3 Knowledge graph of “glucose in COVID-19”

The first level of analysis thus far shows that *glucose* is extensively covered in the CORD-19 dataset and is associated with numerous key events in the infection process of coronaviruses in general. Our next level of analysis aimed to understand to what extent, and how, glucose is associated specifically with COVID-19. First, we extracted the 3,000 of the most relevant articles in the CORD-19 database using a customized ML semantic search with the phrase “glucose as a risk factor for COVID-19,” and then subjected these articles to entity extraction, that yielded over 20,000 entities extracted. We then constructed a knowledge graph as above, but using only the 1,500 most frequent entities, from these 20,000 extracted. Since the resulting graph is still extremely dense (see CORD-19 Knowledge graphs in Methods), we constructed a minimum spanning-tree (see Methods) from the 150 most frequent entities (Figure 4) to allow focusing only on the most important associations between entities, in the context of “glucose as a risk factor for COVID-19”. The tree structure that emerged reflects those associations that survive the greedy minimum spanning tree algorithm, guided us to the most frequent entities linking glucose to various aspects of the disease. For example, we could identify *hyperglycemia* as the main entity that links *glucose* to all groups at risk for COVID-19 (i.e. DM, obesity, hypertension and cardiovascular disorder) in this dataset. It also shows links from *glucose* to *immune responses*, *inflammatory processes*, and *oxidative stress* in one part of the tree, *vascular system* and *thromboses* in another, and to *airways of the lung*, *ARDS*, *multi-organ failure* and *death* in another part, among other important entities (Figure 4).

**Figure 4:**
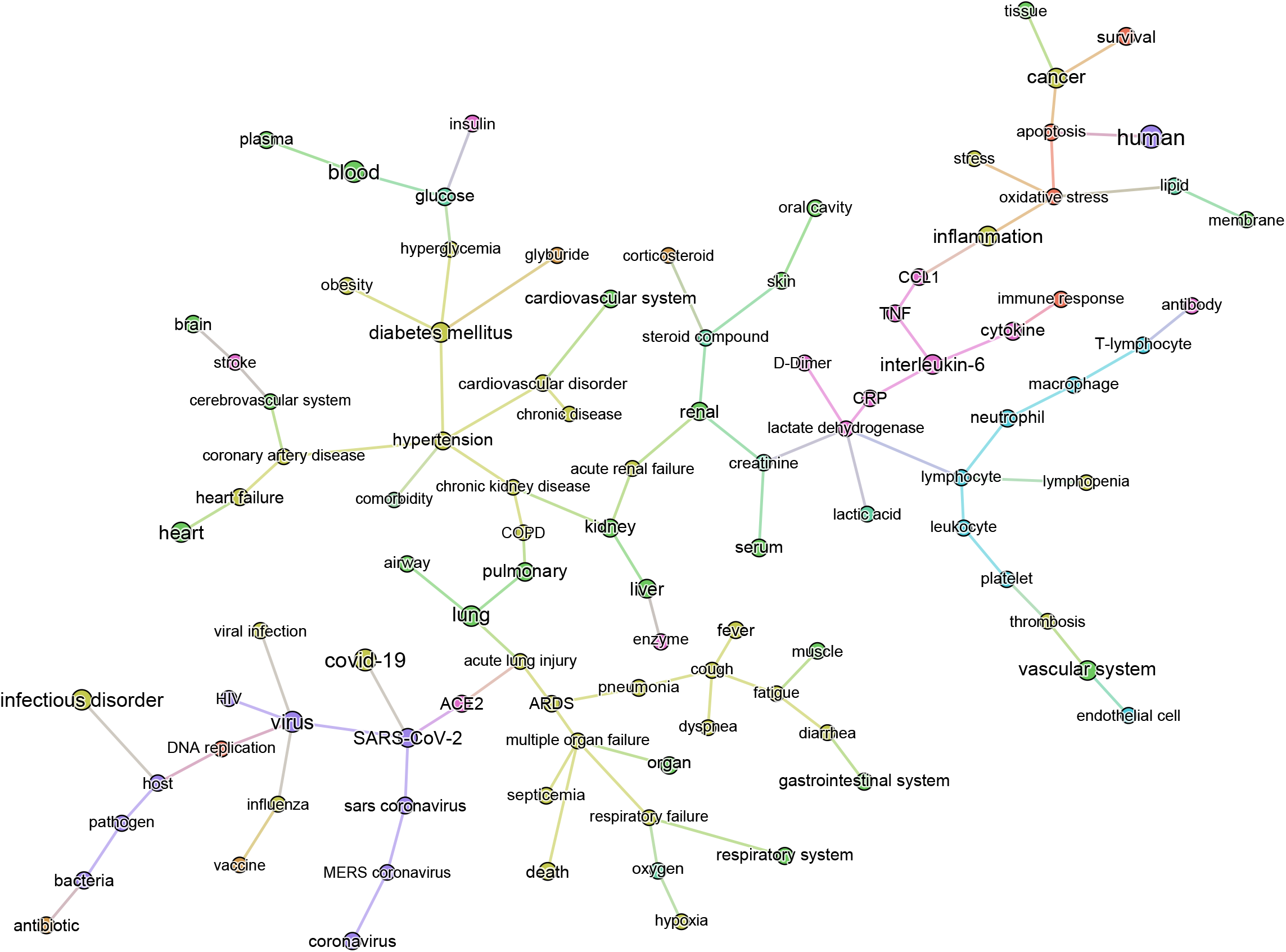
Minimum spanning-tree constructed from the knowledge subgraph containing the 150 most frequently mentioned entities. The knowledge graph is built from the 3,000 most relevant papers related to the query “glucose as a risk factor for COVID-19”. The spanning tree is obtained by minimizing an edge distance score based on mutual information (see Methods). The entity types are color-coded as presented in Figure 2.

From the graph created with the 1,500 most frequent entities, we next identified the top 25 BMIPs from *glucose* to *COVID-19* as further analysis. Figure 5 shows all the most important entities linking *glucose* to COVID-19 in this subset of the CORD-19 dataset. Most of the entities are pathologies and biomarkers known to be associated with COVID-19. The BMIP graph in Figure 5 shows the context in which glucose is found in the CORD-19 dataset obtained from a search for “glucose as a risk factor for COVID-19”. On the other hand, BMIPs subgraphs of entities linking glucose to COVID-19 according to each entity-type (as described in Figure 2), show the strongest associations with the immune defense of the lung through the entities *respiratory system*, *alveolar epithelium*, *innate immunity*, *alveolar cell type II*, *immune cells*, *interleukins*, *chemokines* among others (Figure 6). The subgraphs also show strong associations between *glucose* and the entities that concern viral entry and replication: entities such as *glycosylation*, *glycolysis*, *glucose uptake*, *lactic acid* or *lactate dehydrogenase*. Finally, the subgraphs show associations with *COVID-19 symptoms* and complications through the entities *inflammation*, *CRP*, *ARDS*, *cardiovascular complication*, *thrombosis* and associations with the vasculature by the entities *vascular system*, *fibrinogen*, *D-dimer*, *ferritin*, *platelet* or *endothelial cells*.

**Figure 5:**
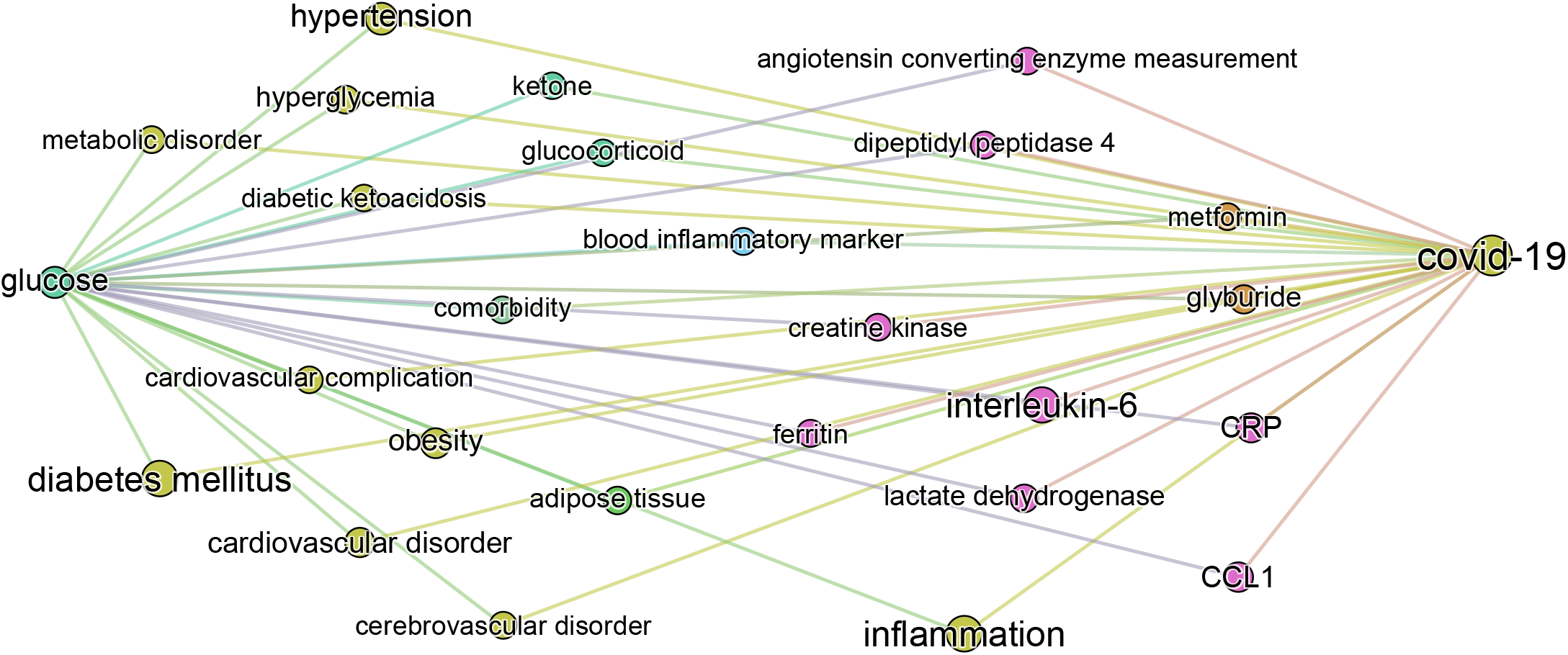
Top 25 BMIPs from glucose to COVID-19 in the 1,500 most frequent entities extracted from the query “glucose as a risk factor for COVID-19”. The size of a node indicates the frequency of the corresponding entity in the COVID-19 literature (measured by the *weighted degree centrality* of the node), whereas the thickness of the edge indicates the strength of association between a given pair of entities (corresponding to the non-negative pointwise mutual information, see Methods for more details). Their position and distance have no significance.

**Figure 6:**
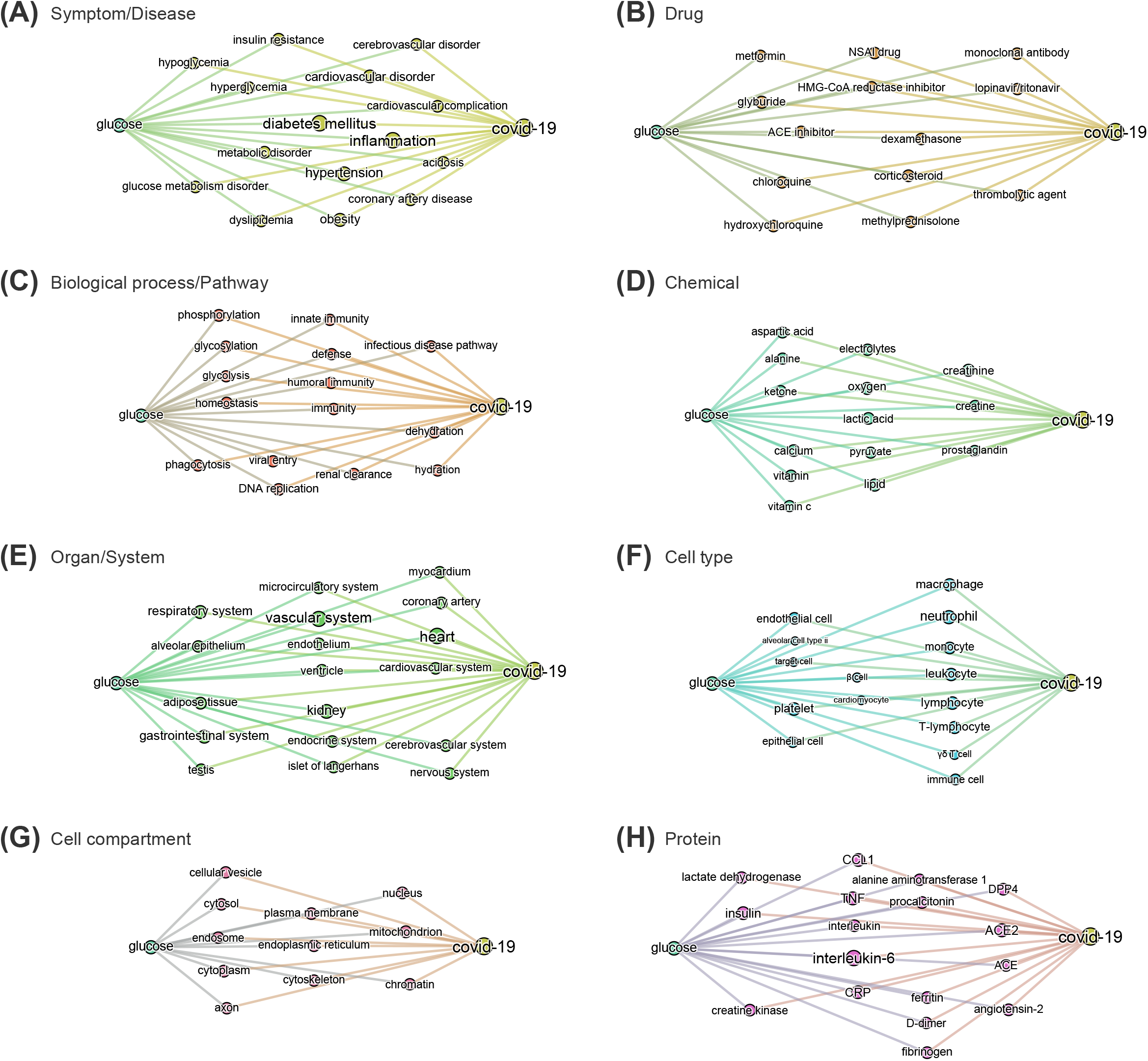
**BMIPs subgraphs from glucose to COVID-19 in each entity type,** such as presented in figure 5.

The subgraphs linking *glucose* to COVID-19 generated according to entity types additionally guided us to and through the specific symptoms, drugs, pathways, chemicals, organs, cell-types, cell compartments, and proteins where the strongest associations with glucose exist in the dataset (see Figure 6A-H). For example, the associations found in the context of the phrase “glucose as a risk factor in COVID-19” in the o*rgan/system* entity type, includes all organs known to be affected in COVID-19. In the entity type p*athways*, we find homeostatic, immune, infectious pathways as well as other biochemical and metabolic pathways where glucose is known to be involved.

The minimum spanning tree enables a different and deeper way to navigate the knowledge contained in the dataset. We therefore again constructed a minimum spanning tree, but this time from the entire knowledge graph containing all the 1,500 most frequent entities. This allowed us to then zoom into specific entities and navigate to deeper associations in the dataset (Supplementary figure 4 and Figure 7). For example, a zoom-in on the entity *glucose* reveals the entities *prediabetes*, *glucose tolerance test*, *homeostatic process*, *HbA1c, insulin* or *beta-cells* as key entities associated with the groups at risk to COVID-19 (Figure 7A). Zooming in on *lung* and *alveolar epithelium,* close entities in the spanning tree, reveals *airway*, *mucociliary clearance*, *airway surface liquid*, *mucus*, *alveolar macrophage*, *lung surfactant*, *surfactant protein-D*, *phagocytosis* and *alveolar cell-type II* as key entities associating glucose in the lung (Figure 7B-C). Zooming in on viral entry (Figure 7D) reveals *viral load*, *DNA replication*, *S protein*, *glycoprotein*, *carbohydrates*, *lectin* or *glycosylation* as key entities in the viral entry process.

**Figure 7:**
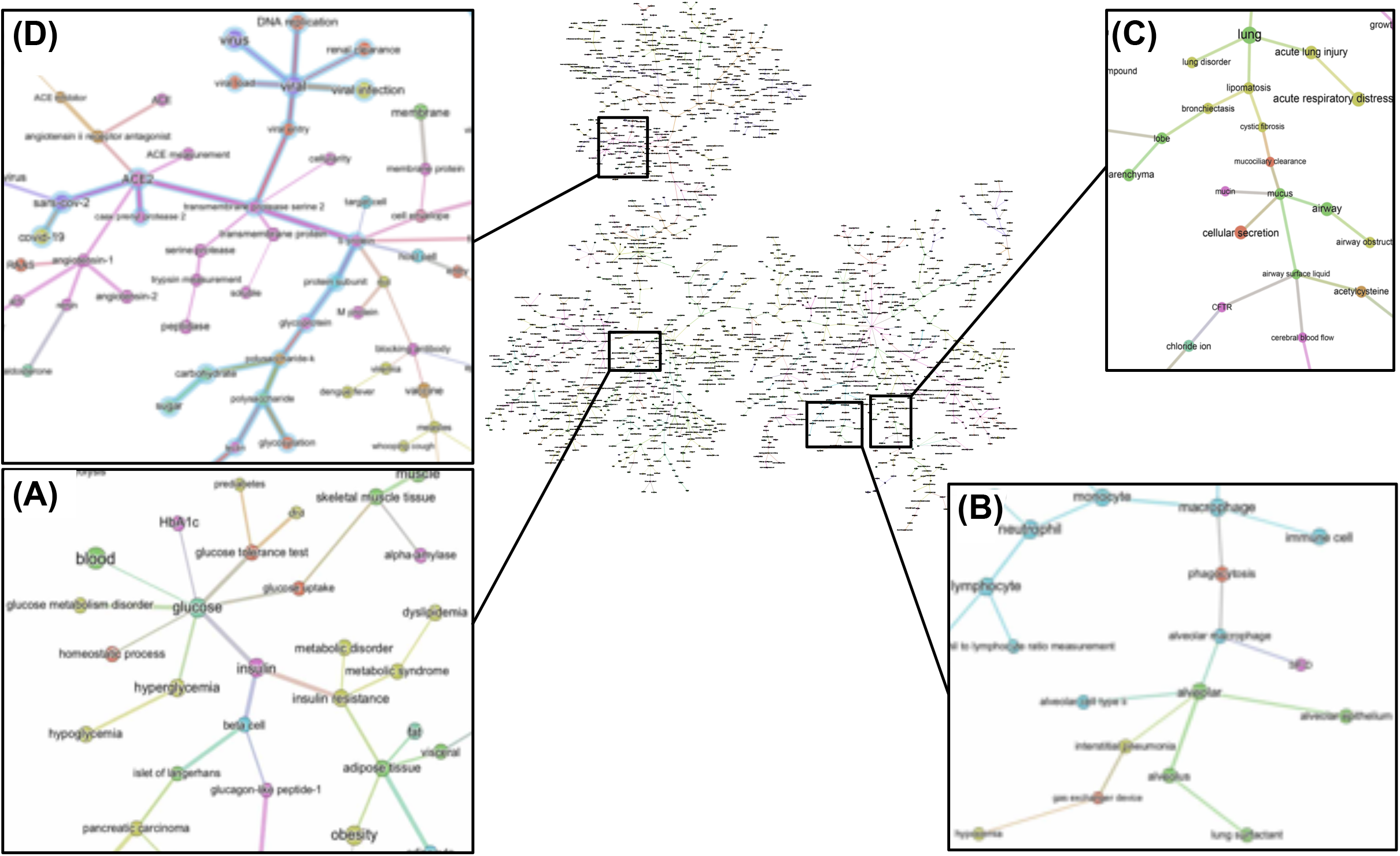
Minimum spanning-tree and zoom-in. Minimum spanning-tree is constructed from the 1,500 most frequent entities extracted from the 3,000 most relevant papers in the CORD-19 database, according to the query “Glucose as a risk factor for COVID-19” (see also supplementary Figure 4). Selected zoom-in for “glucose” **(A),** “alveolus” **(B),** “lung” **(C)** and “viral entry” **(D)** are shown. A high-resolution pdf version of the spanning-tree is available (Figure S4 high res).

To summarize, the knowledge graphs generated from the CORD-19 database enables navigation of the contents of the CORD-19 dataset in terms of entities, different associations between entities, and in the specific context of “glucose as a risk factor for COVID-19”, and enable instant access to the underlying paper(s) and the specific text where these entities are mentioned. We chose this way to construct the knowledge graphs because it finds all types of meaningful associations for an objective view of the dataset, rather than focusing our extraction on a predetermined subset of association types that may bias the view. The methodology used delivers unbiased access to all entities and their associations in over 240,000 scientific papers that are relevant to a potential role played by glucose in the infection. We complemented this representative review by research in the general literature, analyses and computational modeling of specific parameters extracted from multiple papers and using atomistic reconstructions of the virus and its immediate environment to gain a deeper insight into the biophysical constraints that may need to be considered.

### 2.2 Overview of blood glucose metabolism in high-risk patients

Two measures are frequently used as indicators of glucose metabolism: FPG (fasting plasma glucose), that is the blood glucose concentration after a minimum fasting period of 8 hours, and PPG (postprandial plasma glucose), the blood glucose concentration one or two hours after a meal or ingestion of a bolus of glucose. Under normal conditions, FPG values range from 4.4 to 6.1 mmol/L (79 – 110 mg/dL) (average of 5.5 mmol/L), and PPG values should be lower than 7.8 mmol/L (<140 mg/dL). Hyperglycemia is generally diagnosed when FPG is >7 mmol/L (>126 mg/dL) or PPG >11 mmol/L (>190 mg/dL). Such a high FPG value is sufficient to diagnose chronic hyperglycemia, however normal or modestly elevated blood glucose (FPG ranging 6.1 to 7 mmol/L or PPG ranging 7.8 to 11 mmol/L), called impaired fasting glucose (IFG), could reveal an impaired glucose tolerance (IGT) that leads to greater and more frequent glucose fluctuations than normal (91). Because there are no symptoms of IGT, many people with this condition are unaware of their condition. IGT is diagnosed following an oral glucose tolerance test (OGTT), the measure of blood glucose concentration two hours after ingestion of a standardized bolus of glucose (usually 75 g) to detect how quickly the body can clear the glucose from the blood. IGT is indicated when OGTT is between 7.8 to 11 mmol/L and could be a sign of pre-diabetes or other metabolic disorders.

As mentioned, aging (>60 y.o.), hypertension, cardiovascular diseases, DM and obesity are strong risk factors for more severe symptoms and higher death rates upon SARS-CoV-2 infection. We find that the literature strongly supports abnormal FPG, IGT or hyperglycemia in all these conditions as described below.

#### 2.2.1 Aging

A hallmark feature of COVID-19 is its preferential impact on the elderly, but the reason is not clear. One of the many changes that occur with aging, is a steady increase in FPG and PPG, increased rate of IGT (92–94), as well as an increase frequency of asymptomatic hyperglycemia (95, 96). FPG reflects the steady-state of blood glucose, while PPG reflects how well perturbations in glucose levels are tolerated, or the capacity to clear sudden elevations in glucose. To examine the variations in blood glucose metabolism with age, we compiled data from several papers (Supplementary Table 5) and plotted the average trajectories of FPG and 2h PPG (following an OGTT test) within different age ranges (See Figure 8A-C). Figure 8A shows that FPG concentration increases linearly by approximately 0.165 mmol/L per decade starting from around 30 years old, with a non-significant difference between gender (Figure 8B). On the other hand, 2h PPG concentration only changes marginally until the age of around 60, but then starts increasing markedly by around 0.64 mmol/L per decade (Figure 8C).

**Figures 8:**
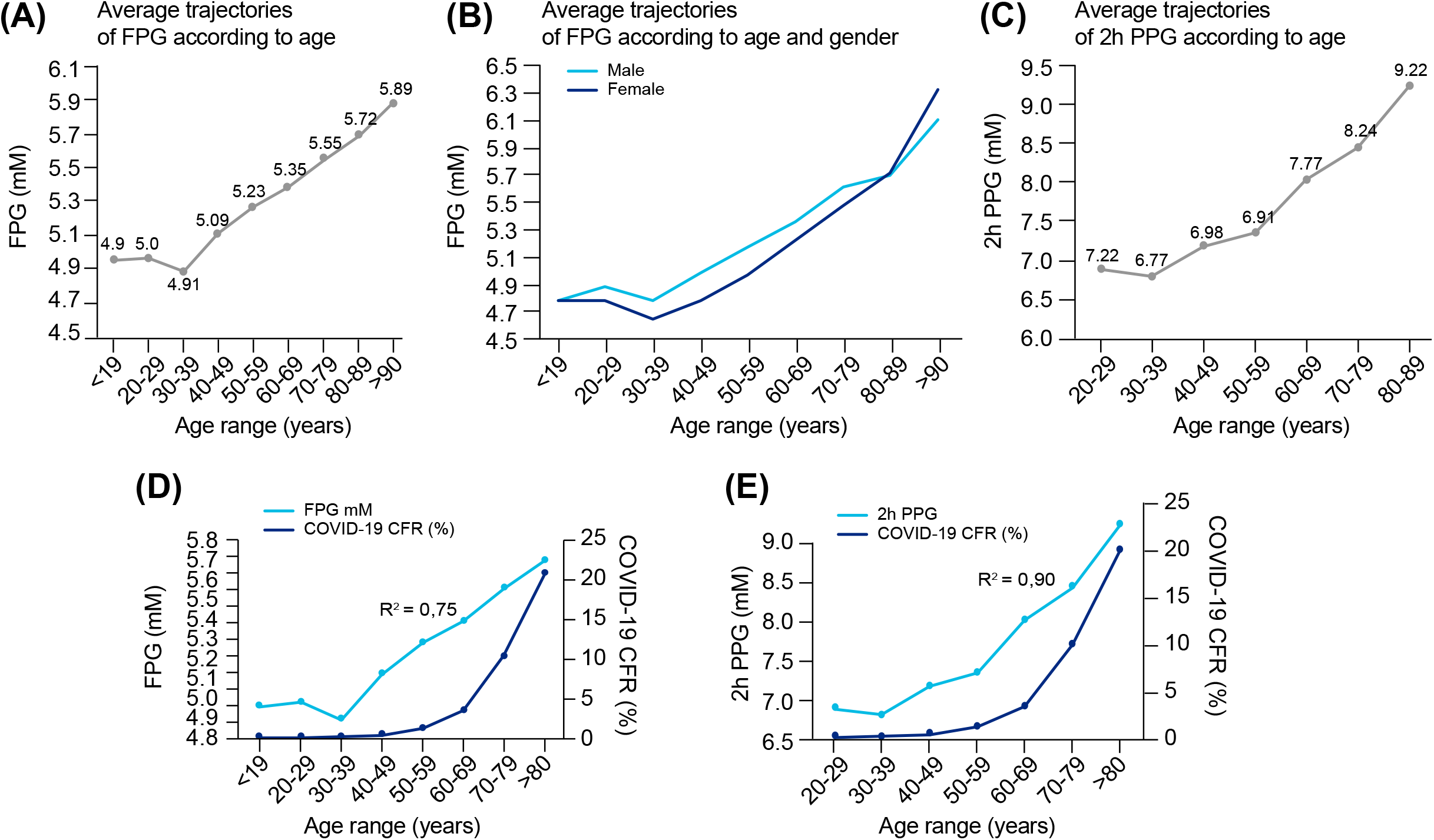
Blood glucose concentrations according to age and gender and correlation with COVID-19 fatality. **(A)** Average FPG values per age range and **(B)** according to gender. **(C)** Average 2h-PPG values after an OGTT test per age range. Data sources are available in Supplementary Tables 5. **(D)** Overlay of COVID-19 CFR and FPG in function of age ranges (from data reported in panel 8A and Supplementary Figures 1B). (**E)** Overlay of COVID-19 CFR and 2h-PPG in function of age ranges (from data reported in panel 8C and Supplementary Figures 1B). In **(D)** and **(E)**, the correlation coefficient “R” is calculated between the two respective series of values for the same age range, then R^2^ = R^2.

The fatality rate for COVID-19 with age of the patient is well characterized by an exponential increase and also increases dramatically after the age of 60 (Supplementary Figure 1B). We then evaluated the correlation between age-related COVID-19 CFR and blood glucose values according to age range. The correlation coefficient between age-related COVID-19 CFR, is 0.87 (R^2^ = 0.75) with age-related changes in FPG (Figure 8D), and 0.95 (R^2^ = 0.90) with age-related changes in 2h PPG (Figure 8E). While these correlations are high, they are not identical, suggesting that the correlation with reduced capacity for glucose metabolism alone is not sufficient to explain the exponential increase in CFR. Indeed, the exponential increase in CFR is likely due to the convergence of multiple factors, including age-related comorbidities. However, if a compromised capacity for glucose metabolism underlies the contribution of the age-related comorbidities, then elevated glucose levels could be considered a fundamental indicator of CFR. We therefore next reviewed the extent to which dysregulation of glucose metabolism is common among some of the known comorbidities of COVID-19.

#### 2.2.2 Diabetes mellitus (DM)

DM is one of the comorbidities that is a clear risk factor for COVID-19 mortality since around 50% of DM patients, independent of age (Supplementary Figure 5A), succumb to the disease. A persistent hyperglycemia (FPG > 7 mmol/L or PPG > 11 mmol/L) is the hallmark of DM. Because hyperglycemia could also be acute, glycated HbA1c (glycated hemoglobin), a measure of the glycemic variation over the past 2-3 months, is an additional marker that is used in the diagnosis of DM (97). Hence, FPG > 7 mmol/L with HbA1c > 6.5 % is the definite indicator of DM, whereas FPG > 7 mmol/L with normal HbA1c (< 6%) reflects an acute hyperglycemia without DM. Indeed, non-diabetic acute hyperglycemia is often asymptomatic and therefore undiagnosed, but could mask an impaired glucose tolerance (IGT). Finally, HbA1c ranging between 6 - 6.4% is a sign of pre-diabetes. Figure 9A shows the average values of FPG, 2h-PPG and Hb1Ac in the diabetic population.

**Figure 9:**
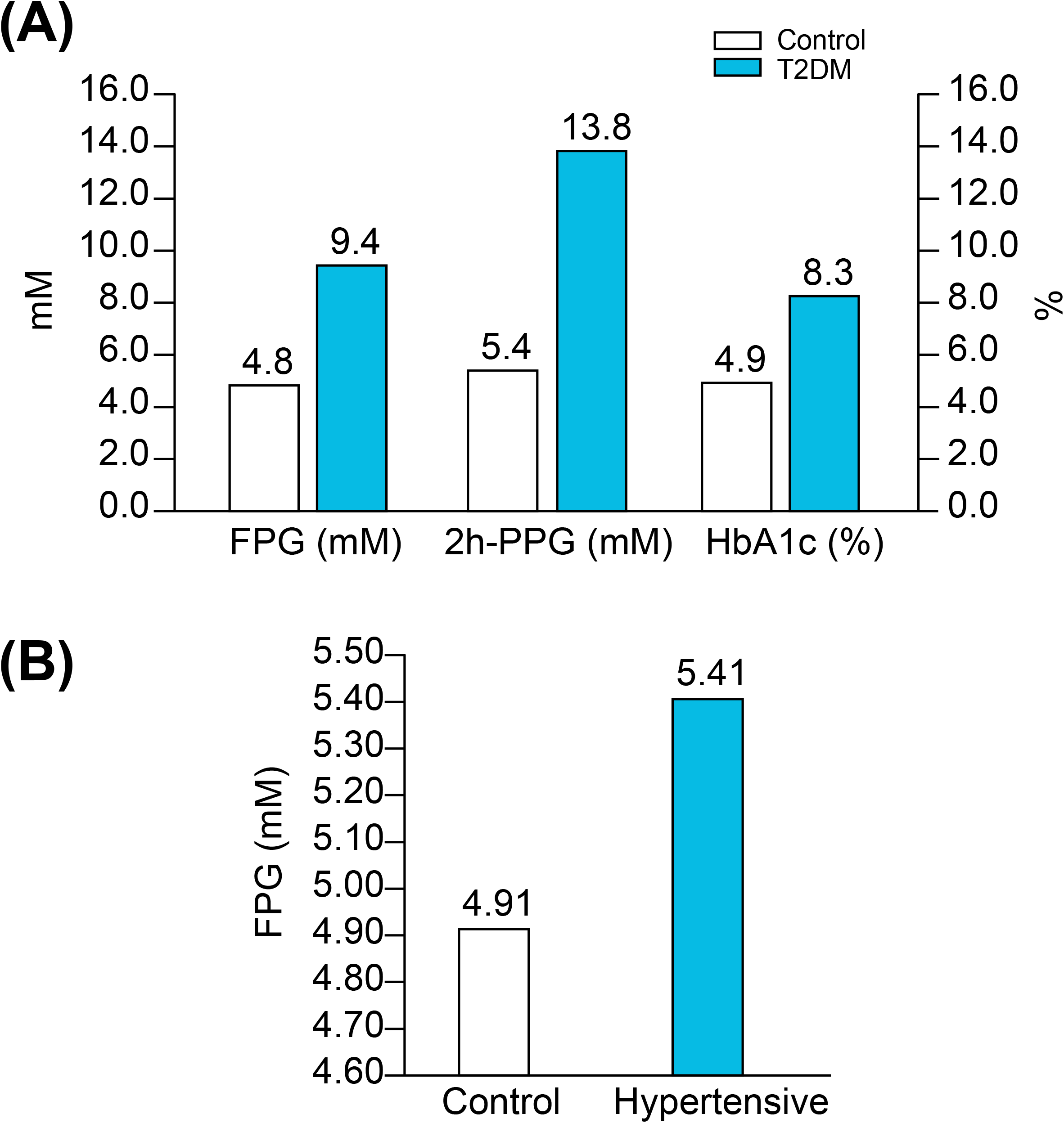
Blood glucose in diabetes and hypertension. **(A)** Average values of FPG, 2h-PPG and HbA1c in diabetic people compared to controls (See detailed references in Supplementary Table 4A). **(B)** Average values of FPG in hypertensive patients compared to controls. (See detailed references in Supplementary Table 4B).

#### 2.2.3 Hypertension

Hypertension is the second most frequent comorbidity in COVID-19 related deaths (25) and is also correlated with age (Supplementary Figure 5A); in patients < 44 years, 35% of the deaths are associated with hypertension, but in patients > 75 years, the association rises up to 70% of the deaths. Hypertension is one of the most prevalent conditions found in the general population (from 20 to 45% depending on the country (98), and is positively correlated with advancing age (99, 100). In addition, hypertension frequently coexists with the other risk factors such as DM, overweight and obesity (96, 101–103). Indeed, a high proportion of COVID-19 patients present with both diabetes and hypertension (Supplementary Figure 5A). It is therefore difficult to separate hypertension as a risk factor by itself from its association with advancing age and other age-related comorbidities, and there is a real need to address if hypertension, by itself, is an independent risk factor for COVID-19 mortality (104, 105). However, what is clear in the literature is that hypertension is strongly associated with poor glucose metabolism.

Firstly, a hypertensive state is positively associated with increased FPG (Figures 9B and Supplementary Figure 6A) and in parallel, higher levels of glucose are considered to be one of the causes of hypertension (106). Secondly, in 70% of cases, hypertension is associated with a disturbance in glucose metabolism (previously known or newly diagnosed DM (25%), IGT (22%), insulin resistance (9%) or IFG (11%) (107)). Thirdly, a study on 63,443 men (ages 21 to 60 years) showed that IFG increases more with age if blood pressure is also elevated (100) (Supplementary Figure 6B). Dysfunction in glucose metabolism in hypertensive patients is therefore frequent and often undiagnosed because an OGTT test to detect IGT is not commonly conducted in the management of hypertension (108). Additionally, some β-blockers, the first drugs prescribed in the management of hypertension, have the common side effect of inducing acute hyperglycemia (109, 110).

#### 2.2.4 Obesity

Overweight and obesity are risk factors for COVID-19 complications and mortality (111, 112). Overweight is defined as a condition where the body mass index (BMI) is between 25-30, while obesity is indicated when BMI > 35, and severe obesity when the value exceeds 40. BMI is positively correlated with FPG levels (113–115) (Supplementary Figure 6A). Mild or severe obesity is directly correlated with hyperglycemia and the incidence of diabetes (93), and IGT is a common finding in obese patients (116). Additionally, it was shown that the incidence of IGT and DM increase proportionally with BMI (i.e. the study shows 20% and 1% IGT and DM incidence respectively for BMI >21; 29% and 6% for BMI ranging 25-26.9; and up to 55% and 20% for BMI >31 (from Figure 1 in Rosiek et al, 2015 (117)).

#### 2.2.5 Intensive care

Patients in ICU have a high risk of hyperglycemia, independent of a history of diabetes, due to the stress of the disease and/or hospitalization (termed “stress hyperglycemia”) (118), or due to the enteral or parenteral feeding that is commonly rich in glucose (95, 119–121); and hyperglycemia has been reported to predict a poor prognosis for diverse critically ill patients (122–124). Additionally, common drugs, that are sometimes used for the treatment of severe viral infection such as catecholamine vasopressors, and some immunosuppressants and corticosteroids, can predispose patients to hyperglycemia (95, 125, 126).

A review of the literature thus far shows that different groups known to be at risk for severe COVID-19 are all likely to present with some level of hyperglycemia, impaired fasting glucose (increased FPG), or IGT (Figure 10), suggesting that reduced glucose metabolic capacity and/or induced elevations in blood glucose could explain why the known preconditions are risk factors for COVID-19 complications and mortality. Findings on the role of glucose during the previous SARS-CoV-1 and MERS outbreaks and preliminary reports on COVID-19 pathogenesis further support this hypothesis. Firstly, even a mild increase in FPG (5.78-7.9 mmol/L) was linked to increased morbidity and mortality in SARS-infected patients during the 2003 outbreak (27). Secondly, corticosteroids, with hyperglycemia as its most common side-effect (127), may exacerbate the severity of the disease and were contra-indicated drugs for the treatment of MERS and SARS-CoV infections (128). Thirdly, it was reported in China, in a small cohort, that 52% of patients presenting clinical characteristics of COVID-19 were hyperglycemic (12). Finally, numerous more recent studies showed that increased FPG is associated with a poor prognosis and increased risk of death from COVID-19, whereas well-controlled FPG is associated with a better outcome (Table 2). More importantly, not only diabetes or hyperglycemia, but IFG specifically has been associated with a higher risk of poor outcome and mortality (53), suggesting that even a modest increase in FPG is a prognostic indicator.

**Figure 10:**
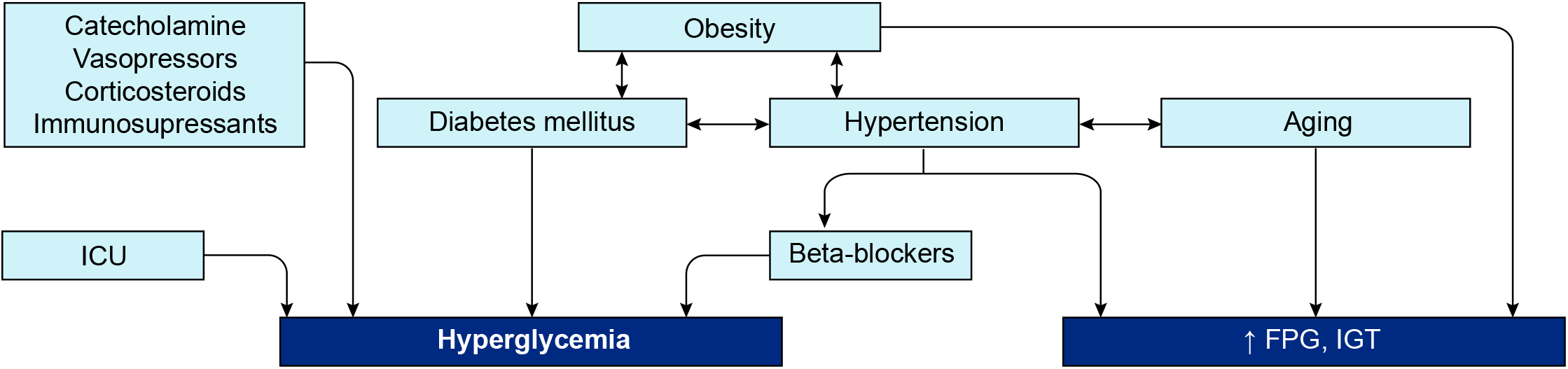
Interconnection of groups at-risk and their link with dysregulation of blood glucose metabolism.

**Table 2:** Overview of the recent studies showing the association between increased FPG and a poor prognosis and/or increased risk of death from COVID-19.

| Reference | Country | Sample size | Patients characteristic | Follow-up duration | Outcomes | Finding |
| --- | --- | --- | --- | --- | --- | --- |
| Bode et al, 2020 | U.S. | n = 1,122 | DM or uncontrolled hyperglycemia vs. non-DM or no uncontrolled hyperglycemia | 37-days follow-up | Mortality | Uncontrolled hyperglycemia is linked to higher mortality rate |
| Li et al, 2020 | Wuhan, China | n = 453 | Normal glucose, hyperglycemia, newly diagnosed DM, known DM | 29.5-days follow-up | Mortality, ICU admission and IMV* | Newly diagnosed diabetes (i.e uncontrolled) had the highest risk of mortality before known diabetes (i.e controlled), hyperglycemia and normal glucose |
| Wang et al, 2020 | Wuhan, China | n = 605 | Non-diabetic | 28-days follow-up | Mortality | FBG is an independent risk factor for COVID-19 mortality |
| Zhang et al, 2020 | China | n = 461 | Diabetic and non-diabetic | 30-days follow-up | ARDS, ICU admission, septic shock or death | FBG is an independent predictor for 30-days poor outcome |
| Zhang et al, 2020b | Wuhan, China | n = 312 | Diabetes, IFG*, NFG* | ND | IMV*, ICU admission or death | IFG is associated with higher risk of poor outcome |
| Wu et al, 2020 | Wuhan, China | n = 2,041 | Non-diabetic | 30-days follow-up | IMV*, ICU admission or shock | FBG is an independent risk factor for COVID-19 progression in critical case or death |
| Liu et al, 2020 | Wuhan, China | n = 255 | All types | From admission (Feb 1-24, 2020) until March 31, 2020 | ICU admission or death | High FPG at admission is a strong predictor for poor outcome |
| Smith et al, 2020 | U.S. | n = 184 | Patients with severe disease | ND | Need for intubation | Impaired glucose metabolism due to DM or obesity is associated with severe COVID-19 |
| Zhu et al, 2020 | Hubei, China | n = 7,337 | Diabetic vs. non-diabetic | 28-days follow-up | All-cause mortality | Well-controlled blood glucose correlated with lower mortality in infected patients |
| Li et al, 2020c | Anheui, China | n = 65 | Moderate case at admission | ND | Transition to severe case | BG is a biomarker for transition to severe case |
| Linli et al, 2020 | Hunan, China | n = 192 | Critically ill patients | Up to death or discharge | Mortality | Higher BG is an independent predictor for mortality |

\*IMV = Invasive mechanical ventilation; IFG = Impaired fasting glucose; NFG = Normal fasting glucose; BG = Blood glucose

In summary, the literature supports the hypothesis of glucose dysregulation as a common factor within all known groups at risk. To now understand whether this is a mere correlation or a cause, we investigated potential mechanisms of action of glucose in the life cycle of the infection. To do so, we traced the various steps of the pathogenesis of COVID-19 that were highlighted by the analyses of the knowledge graphs.

### 2.3 Glucose in the SARS-CoV-2 life cycle

SARS-CoV-2 belongs to the coronavirus family, whose name comes from the shape that the structural spike gives to the virion; protruding as spikes at the surface of the envelope and forming a crown (129). The S protein holds a receptor binding domain (RBD) at the termini of the ectodomain that allows the recognition and binding to its host receptor angiotensin-converting enzyme 2 (ACE2) (130, 131). Each spike is a homotrimer of the S protein, but only one RBD acquires the so-called *up-conformation* to allow the binding to ACE2 (132, 133). Subsequently, a complex sequence of cleavages by host proteases (membrane TMPRSS2 (134) and furin (135)) allows conformational changes of the spike necessary for the subsequent fusion of the virion with the host cell membrane, the cell entry and genome delivery inside the cell for further replication (16, 136). Once inside the cell, the virus relies entirely on the host for energy, and must hijack the cellular machinery of the host to produce more copies of virions. Glycosylation and glycolysis are two key pathways necessary for viral entry and replication and therefore hijacking these metabolic processes is of critical importance for the infection.

#### 2.3.1 Glycolysis as a key mechanism for viral replication

Viruses are nonliving entities and, as such, do not have their own metabolism. Hence, viruses need a supply of nucleotides for genome replication, amino-acids for new protein synthesis, fatty-acids for their membrane, as well as adenosine triphosphate (ATP) for the viral packaging process (137). For this purpose, most viruses have evolved to modify the cellular metabolism of host cells upon entry to increase the availability of energy and nutrients for their own reproduction. One of the most common modifications is the switch to glycolysis as the main metabolic pathway, a fast process for providing the virus with ATP without requiring oxygen, but needing an increase in the uptake of extracellular glucose.

To achieve this, viruses induce glucose transporter expression, glucose uptake, glycolytic enzymes expression (hexokinase 2) and lactic acid production (138, 139), as early as 8-12 hours post infection. The activation of any one of these metabolic pathways is dependent on the cell type infected and on the type of virus (140, 141). The correlation between glucose availability and viral replication is well known, especially for the influenza virus. For example, Reading et al,(142) showed that viral replication of influenza in the lung is proportional to blood glucose concentration. Kohio and Adamson (143) also showed that *in vitro* exposure of pulmonary epithelial cells to elevated glucose concentrations significantly increased influenza virus infection and replication, whereas the treatment of cells with glycolysis inhibitors significantly suppressed the viral replication. Similarly, glucose reduction during infection reduces viral replication (137, 144). Importantly, SARS-CoV-2 replication in monocytes was shown to rely entirely on ATP produced by glycolysis (145). Glucose supply and glycolytic efficiency are therefore crucial parameters for viral replication.

#### 2.3.2 Glycosylation as a key process in viral pathogenesis

Glucose is not only an essential energy and carbon source for viral replication, it is also the precursor for glycan trees synthesis, a key process in viral pathogenesis. N-glycosylation, that consists of the addition of glycan trees at N(X)T/S consensus sites of proteins, is a post-translational modification that affects more than 50% of mammalian proteins, most importantly membrane proteins (146). This modification has a crucial role in ensuring the correct structure and function of the proteins, the regulation of protein-protein interactions, cell signaling, and pathogen-host recognition (147, 148). Glycan trees are hydrophilic structures also conferring a high solubility to secreted proteins. They consist of assemblies of monosaccharides (sugar molecules such as glucose, galactose, N-acetylglucosamine, N-acetylgalactosamine, glucuronic acid, xylose, mannose, fucose or sialic acids; (149)) and can be divided into three main types: 1) the oligomannose types (or high-mannose (HM)), considered to be under-processed glycan trees, that exclusively contain mannose residues and are rarely found in mammalian membrane proteins; 2) the complex types, that are bulky, but flexible trees, containing multiple branches with any number of the other type of saccharides mentioned, and 3) the hybrid types which are composed of one branch of mannose residues and a second branch with complex residues (150). Importantly, glucose, the main monosaccharide in carbohydrate metabolism, can be converted into all the types of sugars required to build glycan trees.

Glycosylation is key in multiple biological mechanisms of viruses (infectivity, virulence, immune interactions among others (151), and implicated in species-to-species transmissibility (152). Transmission of zoonotic viruses into humans are accompanied by drastic changes in glycosylation, as exemplified by the human influenza H3 hemagglutinin where the number of glycosylation sites have doubled since the 1968 pandemic while its amino acid sequence has remained 88% unchanged (153). Glycosylation is essential for particular mechanisms such as maintaining the structural shape of the viruses, recognizing the host cells and binding sites, as well as for cell entry (154–156). It is also used to evade the immune system; indeed, it allows the virus to deceive the humoral and adaptive immune system of the host by imitating its glycosylation coat (in a process called molecular mimicry), and shield its immunogenic epitopes from antibody recognition (152, 154, 156–159). The glycan coat of SARS-CoV S proteins is however relatively sparse compared to strong immune evaders such as HIV or Ebola (152).

Viral glycoproteins are thought to be more heavily glycosylated than host glycoproteins, and the glycan composition can differ from host compositions, and from host to host. The under-processed HM type is, for example, rarely found on the host cells, but frequently found in enveloped virus protein. This is explained because the distribution of oligo-mannose or complex glycans is determined by the accessibility and crowding of the carbohydrate chains, more than the protein sequence itself (160). Indeed, densification of glycans over a protein sequence results in inhibition of glycan processing and poorer conservation of glycan trees across viral copies (161). In SARS-CoV-2, the S protein forming the spike is particularly highly glycosylated, with 22 sites of N-glycosylation per monomer, holding mostly complex-type glycans, and ∼30% oligomannose-type (132, 133, 162, 163).

We used an atomistic visualization tool (BioExplorer, see Methods) to reconstruct the glycosylation profile of the SARS-CoV-2 S protein, in order to obtain a realistic view of the organization of the different types of glycans on the different domains of the spike. Several groups have reported the glycosylation profiling of the protein S (159, 162, 164), with some discrepancies in the reports. This is likely because the glycosylation profile of a protein can differ from cell type to cell type, and because of glycosylation microheterogeneity, i.e. the inherent variation of glycan structure at a specific site (165). In our study, we considered data reported in Watanabe et al, 2020 (162), considering only the most frequently represented glycan type (HM, complex or hybrid) for each specific site, without including microheterogeneity (see Methods, Glycan types and position). The resulting distribution of glycans is schematically represented in Figure 11A (detailed in Methods). This atomistic representation of the glycosylated spike shows the extent to which the spike is physically shielded by glycan trees (Figure 11B-E) making the virus appear as a large sugar molecule to the host.

**Figure 11:**
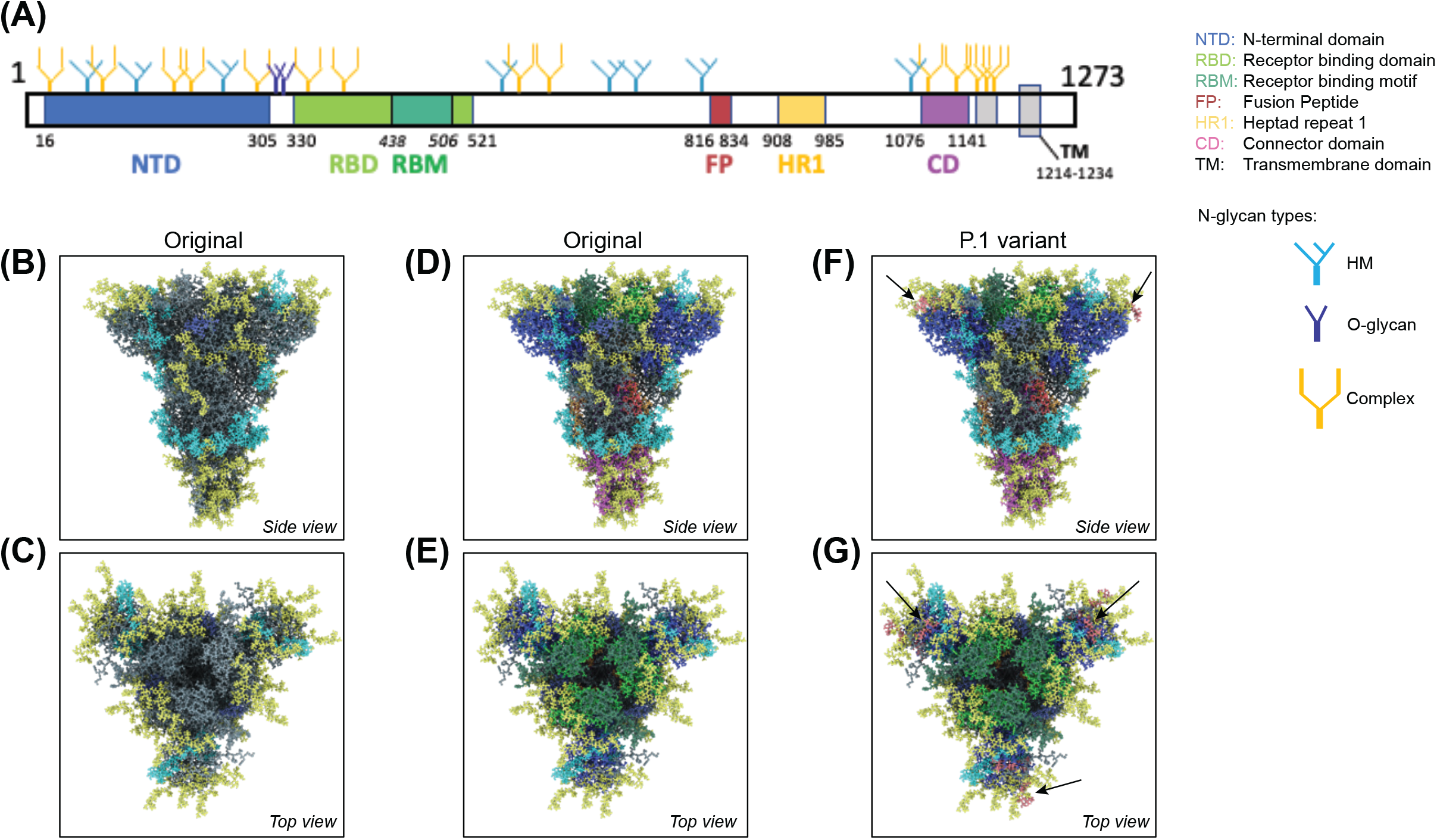
Atomistic reconstruction of the glycosylated SARS-CoV-2 spike. (**A)** Schematic of the primary structure of the SARS-CoV-2 S protein with the different types of glycan-tree positioned on the different domains. (**B), (C)** Side and top view of the SARS-CoV-2 glycosylated spike in closed conformation. The different types of glycan trees are colored according to the legend. (**D)**, (**E)** Side and top view of the SARS-CoV-2 glycosylated spike in closed conformation, with the different domains colored according to the legend used in panel A. (**F)**, (**G)** Side and top view of the modeled glycosylated spike from the Brazilian P.1 variant in closed conformation. The potential additional glycan-tree in position N20 is colored in red and indicated with arrows. Generation of the spike, positions and sources of the glycans are detailed in methods.

Whereas the complex glycans are mostly localized at the extremities of the spikes and around the connector domain (CD), the HM glycans are concentrated around the central core of the spike (in a ring-like formation), and only rarely localized at the extremities (Figure 11A-E). We can reasonably hypothesize that the bulky complex glycans, mimicking the host cell glycan types, are exposed at the extremity to help hide the spike from detection by the immune system. In contrast, under-processed HM glycans, that require less enzymatic processing, could be sufficient to cover less exposed immunogenic domains such as the fusion peptide (Figure 11A, D and E). In addition, the HM types, which are the glycans recognized as foreign by the innate humoral immune system, are logically less exposed than the host-like complex types (159). Complex glycans on the RBD are surrounding the RBM (receptor binding motif), that is itself completely glycan-free to allow binding to its receptor (Figure 11C and E). These complex glycans may also serve a different function such as aiding the recognition and binding to the receptor. Indeed, ablation of two N-sites of the RBD (N331 – N343) drastically reduces infectivity (166). Glycans located on the N-terminal domain (NTD) could also be involved in receptor recognition as molecular dynamics simulations have suggested that, apart from the shielding, glycans at two sites, the N165 and N234 in the NTD, may provide conformational stability of the receptor-binding domain during recognition of ACE2 (167). ACE2 is also glycosylated, holding six putative N-glycosylation sites ((168) and Methods). It has been reported that ACE2 glycosylation does not affect its expression on the cell surface, but it is required for the binding to SARS-CoV-2 glycosylated spike and for fusion with the membrane (169, 170). To gain insight into the involvement of glycans in the spike-receptor interaction, we represented the interaction of the glycosylated spike in its open conformation with glycosylated ACE2 (Figure 12). The domains on the spike and ACE2 involved in the interaction (binding domains) are highlighted in green and blue respectively, showing the accuracy of the models. Interestingly, one can observe that both binding domains are almost exclusively surrounded by complex glycans (Figure 12) that seem to be connected. The complex glycans on the spike might therefore not only serve to protect the ACE2 binding domain when in closed conformation or to stabilize the interaction, but could also enable the conformational change of the protein S into its up-position, required to be able to bind to the receptor. This is in agreement with a recent proposal reported by Casalino et al, (167), that the glycan composition of SARS-CoV-2 spike is crucial for the RBD up/down conformational changes. Similarly, the complex glycans of ACE2, almost all concentrated near the spike interacting domains, may serve to allow and stabilize the interaction with the spike.

**Figure 12:**
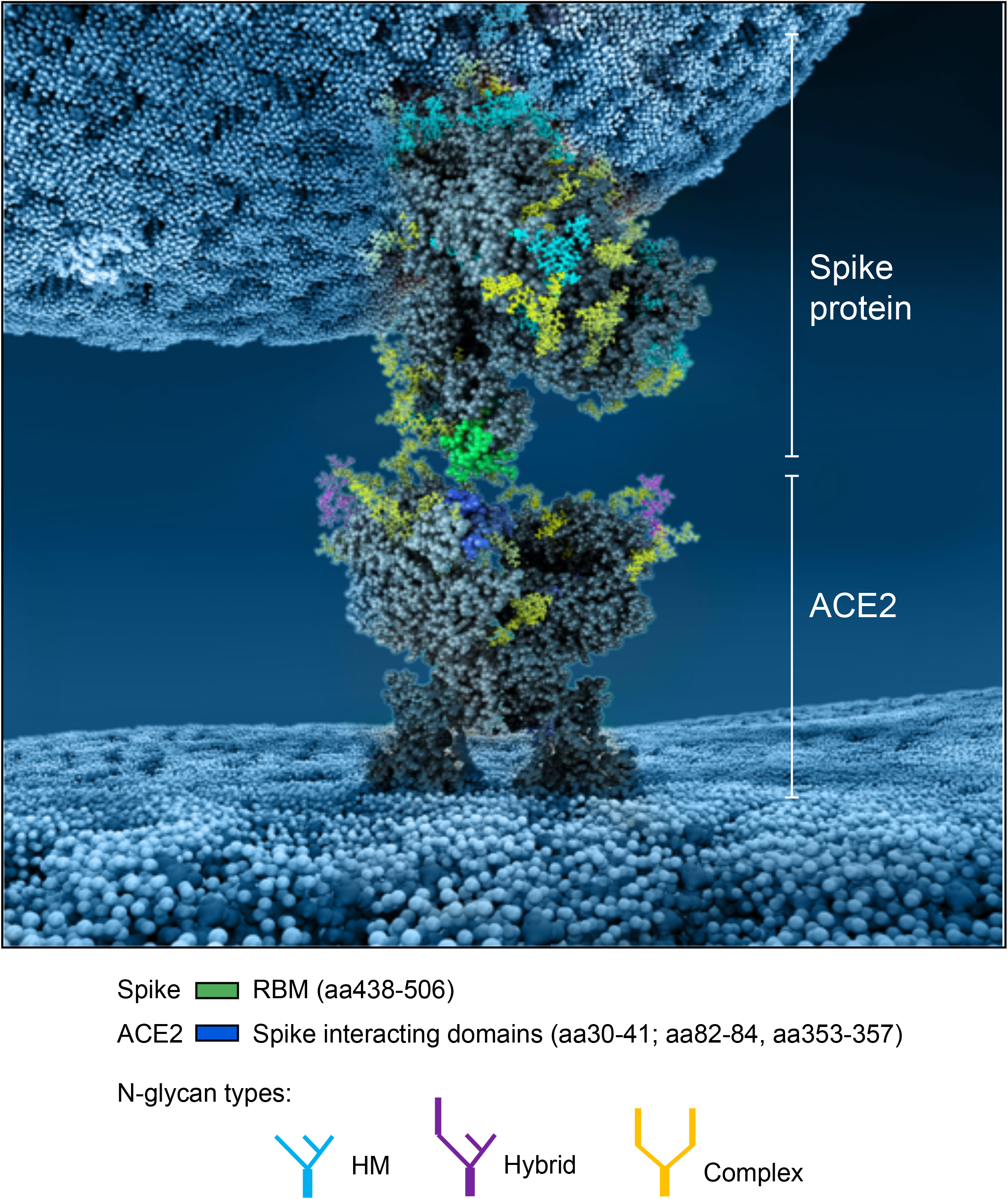
Atomistic reconstruction of the glycosylated spike of SARS-CoV-2 interacting with its glycosylated receptor ACE2. The interaction between a spike in open conformation and ACE2 is represented. The binding domains in the spike and ACE2 involved in the interaction are colored in green and blue respectively. Note that not all glycans are present on the open spike (see Methods for details).

More than an effect in receptor binding and infectivity previously mentioned, mutations of some glycosylation sites are known to render the virus resistant to neutralizing antibodies (171). The S protein is the major antigen responsible for the adaptive immune response (133, 172–174); it is therefore natural to direct vaccines at the spike protein (175). Several amino acid changes in the S protein could affect viral infectivity, transmissibility and efficacy of neutralizing antibodies. If they involve glycosylation sites, then the virus can change its glycan coat needed for infection, transmission, and deceive the host’s immune system. Three variants of SARS-CoV-2 are of particular concern; the variant B.1.1.7 (or 501Y-V1) first emerged in the UK, the variant 501Y-V2 first emerged in South Africa (SA), and the P.1 variant first emerged in Brazil - all with mutations identified in the sequence coding for the S protein (see https://www.ecdc.europa.eu/en/publications-data/covid-19-risk-assessment-spread-new-variants-concern-eueea-first-update). All three variants share the N501Y mutation, located in the RBM, responsible for a more infectious phenotype with higher infectivity but apparently little change in severity (176, 177), due to increased binding affinity for its receptor ACE2. The SA and Brazilian variants hold an additional mutation (E484K), which may be an immune evader mutation (178, 179). They all have undergone additional mutations, but none of these impact glycosylation sites. However, the Brazilian variant, from which there is less information on infectivity, severity, immune evading, possesses a T20N mutation, a mutation located at the beginning of the NTD region, which could potentially become a functional glycosylation site according to the NetNGlyc 1.0 software (http://www.cbs.dtu.dk/services/NetNGlyc/).

In order to better understand the potential impact of this additional glycan site, we modeled the glycosylation profile of the P.1 variant using the BioExplorer tool we developed. Interestingly, the additional 20N glycan would be localized at the very top of the spike (Figure 11F), adding to the shielding of the RBM surrounding region (Figure 11G), which may suggest that it would be better at evading the immune system. In addition, it is localized very close to the N331 and N343 glycans sites (see Supplementary Figure 7 and (166)) shown involved in receptor binding, which may also increase efficacy of receptor binding and account for transmission with lower viral loads. Overall, this potentially new functional N-site, in addition to the well-described N501Y and E484K mutations, could render the P.1 variant more infectious and even a stronger immune evader.

### 2.4 Glucose in the antiviral defense of the lung

SARS-CoV-1 and SARS-CoV-2 are respiratory viruses that mainly invade the human body through droplets first inhaled into the upper airways, where they infect host cells by binding to the host receptor ACE2, and then may migrate to the lower airways where more cells can be more easily infected (180). ACE2 is expressed in many different tissues, but mainly found in lungs, pancreas, kidneys, as well as the gastrointestinal tract and endothelial cells (181–184). Because ACE2 is the entry point for the virus, several studies have focused on the role of the ACE2-spike interaction, the expression level of ACE2, and the glycosylation status of ACE2 to explain the severity of the disease. The data is however inconsistent with no clear correlation between ACE2 expression levels and disease severity (185–189). However, before reaching the lower airway where it can bind to ACE2, the virus has to break through the first non-specific anti-pathogen defense system of the lung formed by the pulmonary epithelium and the airway surface liquid (ASL). This defense system is the first-line protective barrier from constant exposure to bacteria, fungi, viruses and toxic particles (190).

#### 2.4.1 The pulmonary epithelium and the ASL as the first-lines of defense against pathogens

The non-alveolar epithelium of the respiratory zone is composed of many types of secretory cells that produce cytokines, antimicrobial agents as well as mucins forming the mucus (Figure 13) (191–193). This epithelium possesses a high number of ciliated cells with hair-like projections that beat rhythmically, propelling pathogens and inhaled particles trapped in the mucus out of the airways. This process, called mucociliary clearance, is the very first defense that starts in the upper airway and that attempts to expel the pathogen before it can reach the epithelial cells (194, 195). Some pathogens may get through and reach the lower alveoli, where the epithelium is mainly composed of alveolar epithelial cells type I and II (AECI and AECII) along with numerous resident macrophages (193). The thin AECI cover 95% of the alveolar surface area and are largely devoid of organelles since they specialize on passive gas exchange (196), whereas the cuboidal AECII secrete surfactant, a fluid composed of a mixture of proteins and lipids involved in both the maintenance of surface tension, to avoid the collapse of the alveoli, and alveolar protection (197–199). The AECII pneumocytes are the cells of the respiratory tract showing the highest expression of ACE2 as compared to lower levels of ACE2 that are found on the clara cells, the ciliated airway cells and the epithelial cells of the nasal cavity (200).

**Figure 13:**
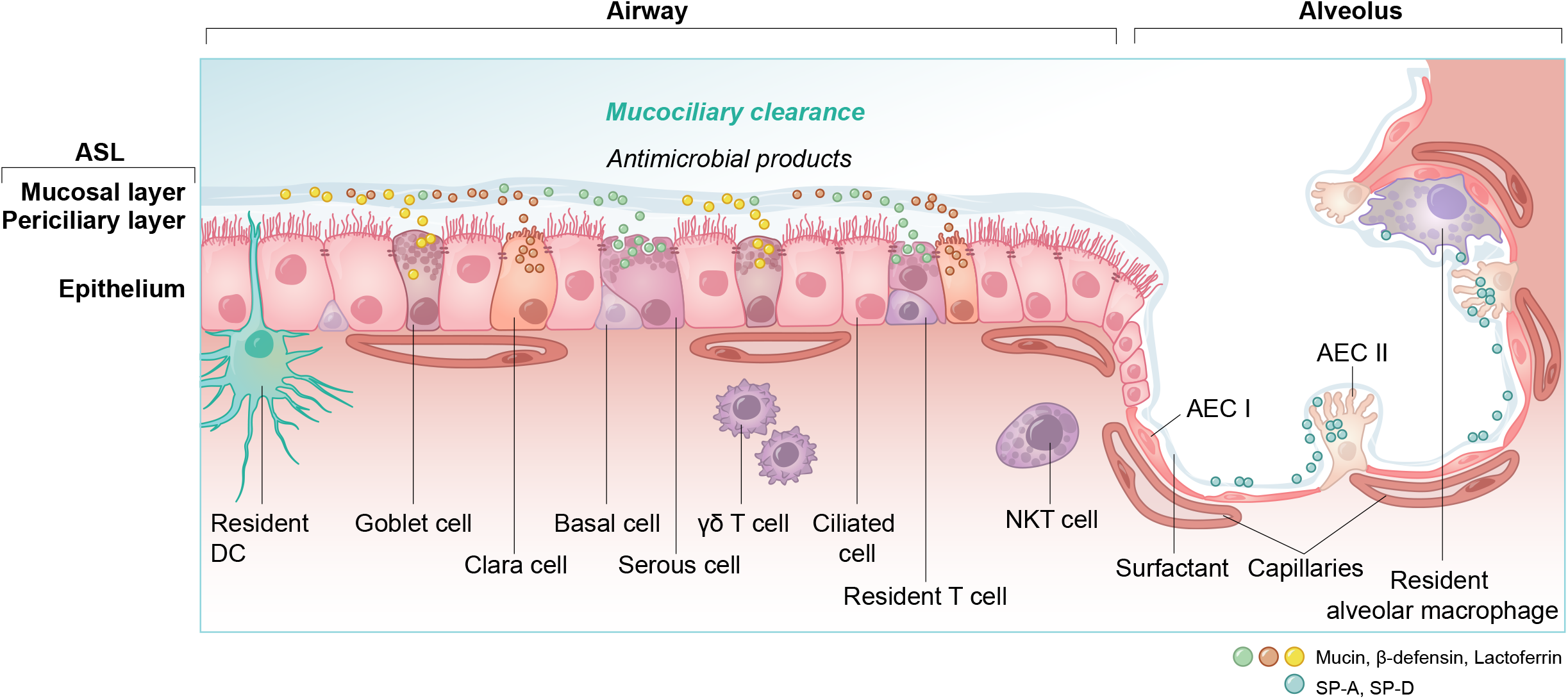
Overview of cell types and innate immunity in the epithelium of the lung. DC = dendritic cell, NKT = natural killer T cell, γδ T = gamma delta T cell, AEC I and AEC II = alveolar epithelial cells, SP-A, SP-D = surfactant protein A and D.

The ASL is composed of a periciliary layer and the overlying mucosal layer, and lines most of the respiratory tract (Figure 13). The mucosal layer is composed of mucins, large glycosylated proteins secreted by the specialized mucosal and goblet cells (191, 201) that form a physical barrier to trap inhaled particles or pathogens. The periciliary layer has a lower viscosity to allow the ciliary beating for mucociliary clearance. The ASL volume, depth and hydration level are critical for the functioning of the mucociliary escalator and these parameters are therefore homeostatically regulated by an intricate orchestration of mucin production and expression of a complex combination of ion channels, exchangers and pumps (see (202) for an extensive review). Na^+^ absorption and secretion of HCO_3_^-^ and Cl^-^ are mediated through the specific transporters ENaC and CFTR (203, 204). Importantly, the deeper alveoli in the lungs are lined with a thin surfactant layer to permit efficient gaseous exchange (198), which contains several other molecules, including amino-acids, proteins, lipids and glucose, all of which are under strict homeostatic control to avoid conditions that would support bacterial growth (205), while ensuring a proper functioning of the ASL. The glucose concentration in the ASL is especially carefully regulated (206).

##### 2.4.1.1 Regulation of glucose concentration in the ASL

Glucose is 10-12 times less concentrated in the ASL than in blood (207). This low concentration of glucose (0.4 mmol/L +/− 0.2 in normal condition) is necessary to maintain the proper functioning and the sterility of the ASL (208). Glucose is exclusively supplied to the airways from the circulating blood, reaching the basolateral side of epithelial cells, where uptake of glucose can occur through glucose transporters (GLUT). The low concentration of glucose in the ASL is tightly regulated by homeostatic mechanisms that include paracellular passive diffusion controlled by tight junction barriers, and facilitative transcellular epithelial glucose transport (Figure 14); the paracellular diffusion being the primary mechanism (209). The transcellular transport of glucose is mediated by the facilitative transporter GLUT, expressed at the basolateral membranes, and by GLUT or SGLT1 at the apical membrane of the airway and alveoli respectively (Figure 14 left panel and (207, 210). Glucose normally moves through GLUTs by passive diffusion down a concentration gradient generated by the activity of hexokinases, which phosphorylate intracellular glucose to maintain a low-intracellular concentration of glucose (209). In contrast, transport via SGLT is driven by sodium (Na^+^) and glucose gradients. This co-transport of Na^+^ in the alveolus would be advantageous for the maintenance of the low volume of fluid required for efficient gaseous exchange (211–213). Several pathological conditions lead to a disruption of glucose homeostasis in the lung and a subsequent increased glucose concentration in the ASL (210, 211). Indeed, defects in tight junction permeability or an increase in blood glucose concentration (hyperglycemia) could both lead to a rise of glucose in the ASL (Figure 14, right panel), with the greatest effect when they coexist. Any elevation is directly countered by apical reuptake by the epithelial cells through the GLUT and SGLT transporters, followed by rapid metabolism by hexokinase in the glycolysis pathway. Hence, the direct conversion of glucose to glucose 6-phosphate (G6P) allows the cells to maintain a steep gradient of glucose concentration needed for a strong driving force for the reuptake of glucose from the ASL.

**Figure 14:**
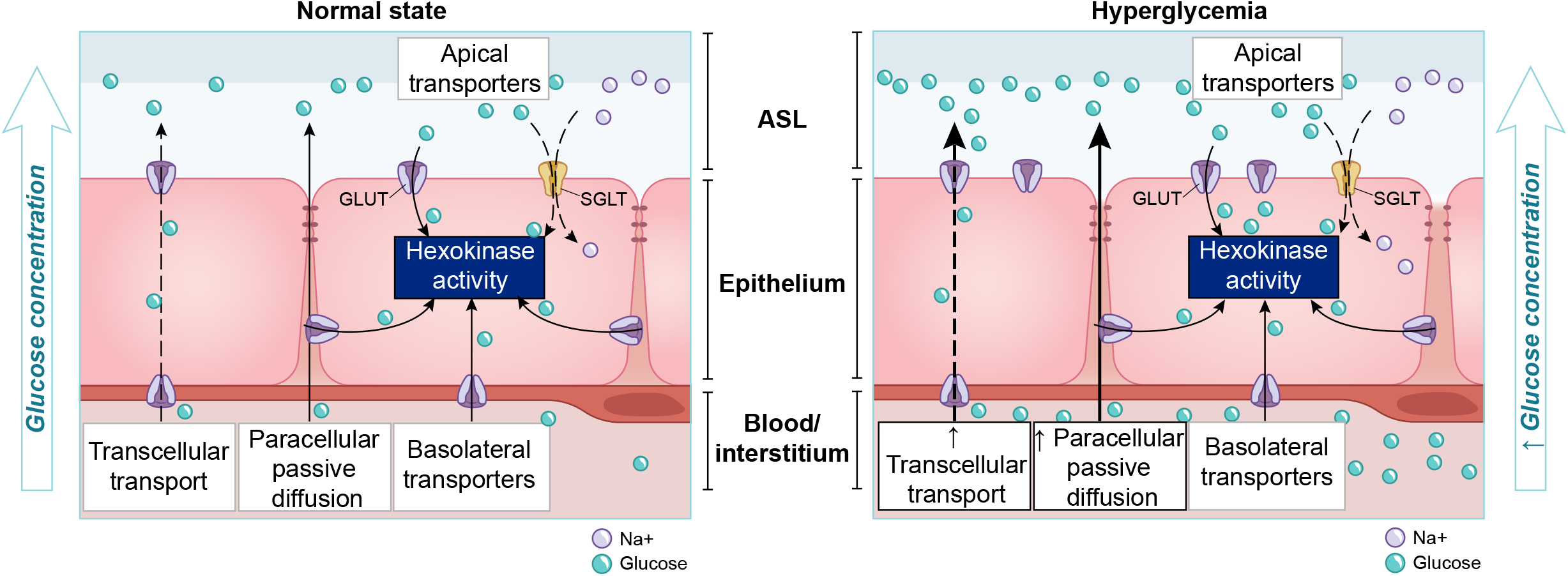
Overview of glucose transport in the lung epithelial cells.

##### 2.4.1.2 Elevated glucose in the ASL impairs primary lung defenses

A high concentration of glucose in the ASL has multiple effects that lead to general impairment in its defense capability (210), as summarized in Figure 15 and detailed below.

**Figure 15:**
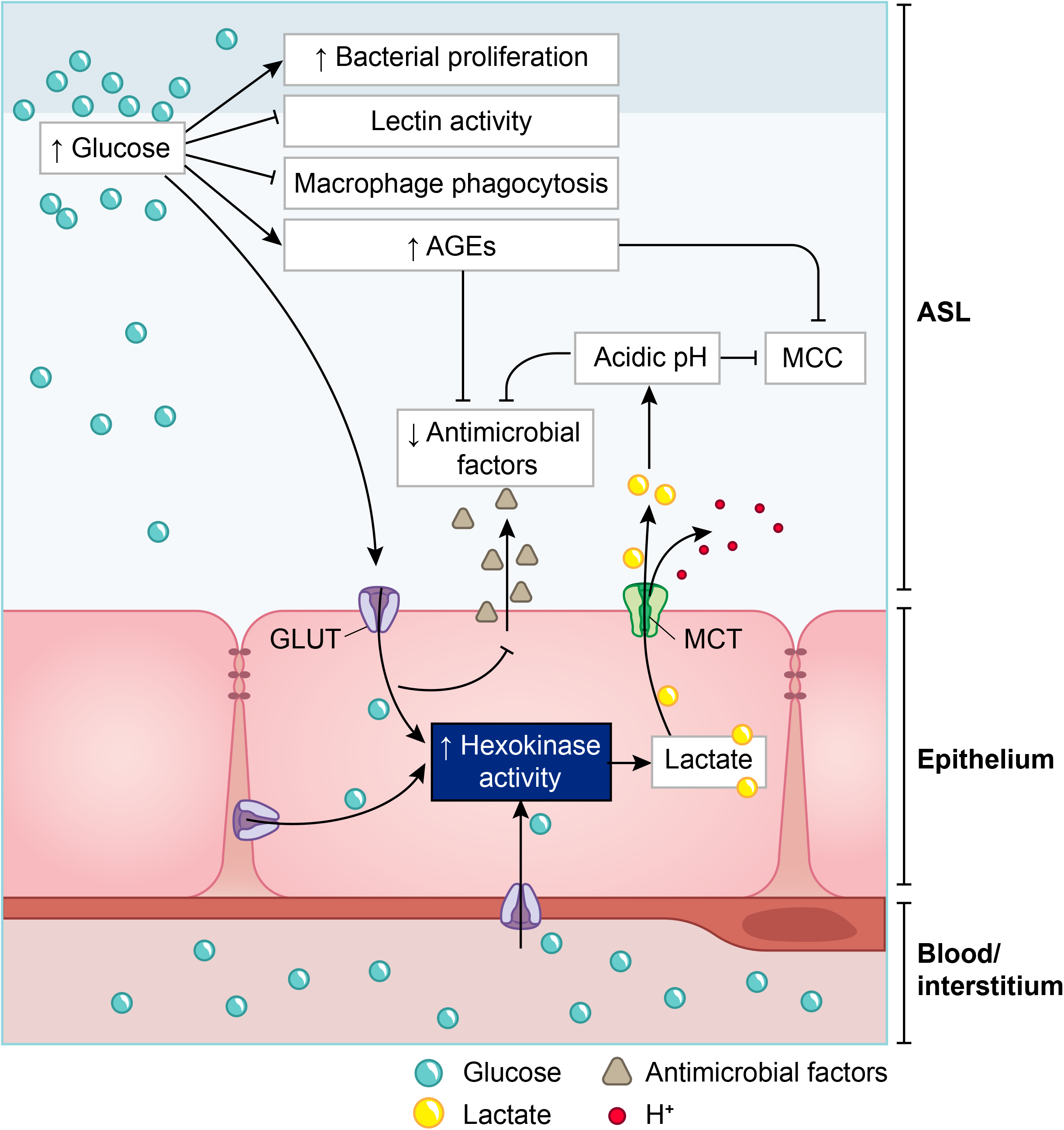
Schematic of the impacts of increase in ASL glucose on ASL functions. AGEs = Advanced glycation end products, MCC = mucociliary clearance. MCT = monocarboxylate transporter.

###### 2.4.1.2.1 Glucose impairs the humoral arm of lung defenses

As mentioned before, the airway epithelial cells secrete a wide range of antimicrobial agents (see Figure 13). The combined activity of these proteins is a crucial step in the first phase of the innate defense of the lung against infections by viruses, bacteria and fungi. Among them, enzymes (lysozymes, proteases), proteases inhibitors, and soluble factors (cytokines, lactoferrin, ß-defensin and LL-37 (cathelicidin-related peptide)) that are dedicated to humoral immunity against a variety of pathogens (190, 192, 214, 215). However, the protection against viruses is mainly mediated by the soluble C-type lectins SP-A and SP-D (surfactant protein A and D, pattern recognition molecules of the collectins family) (216–218), produced by the AECII cells and secreted in the distal alveolar airway (see Figure 13) (219, 220). In case of viral invasion, C-type lectins bind to the high-mannose glycans exposed at the surface of the enveloped viruses through their carbohydrate recognition domain (CRD) (221, 222), and exert their antiviral activity through two different mechanisms: first, by aggregating the pathogens, that physically impairs the binding to the receptors, and second, by recruiting and activating the resident alveolar macrophages, neutrophils and chemo-attracted phagocytes to phagocytose the aggregated viruses (216, 218, 220, 223, 224). SP-A and SP-D show significant differences in ligand preferences; in the case of SARS-CoV, it seems to be mainly targeted by SP-D recognition (218, 225). Using the BioExplorer, and the data reported in the literature, we reconstructed a model of the environment of SARS-CoV-2 in the ASL during primary infection under normal glucose concentration (Figure 16).

**Figure 16:**
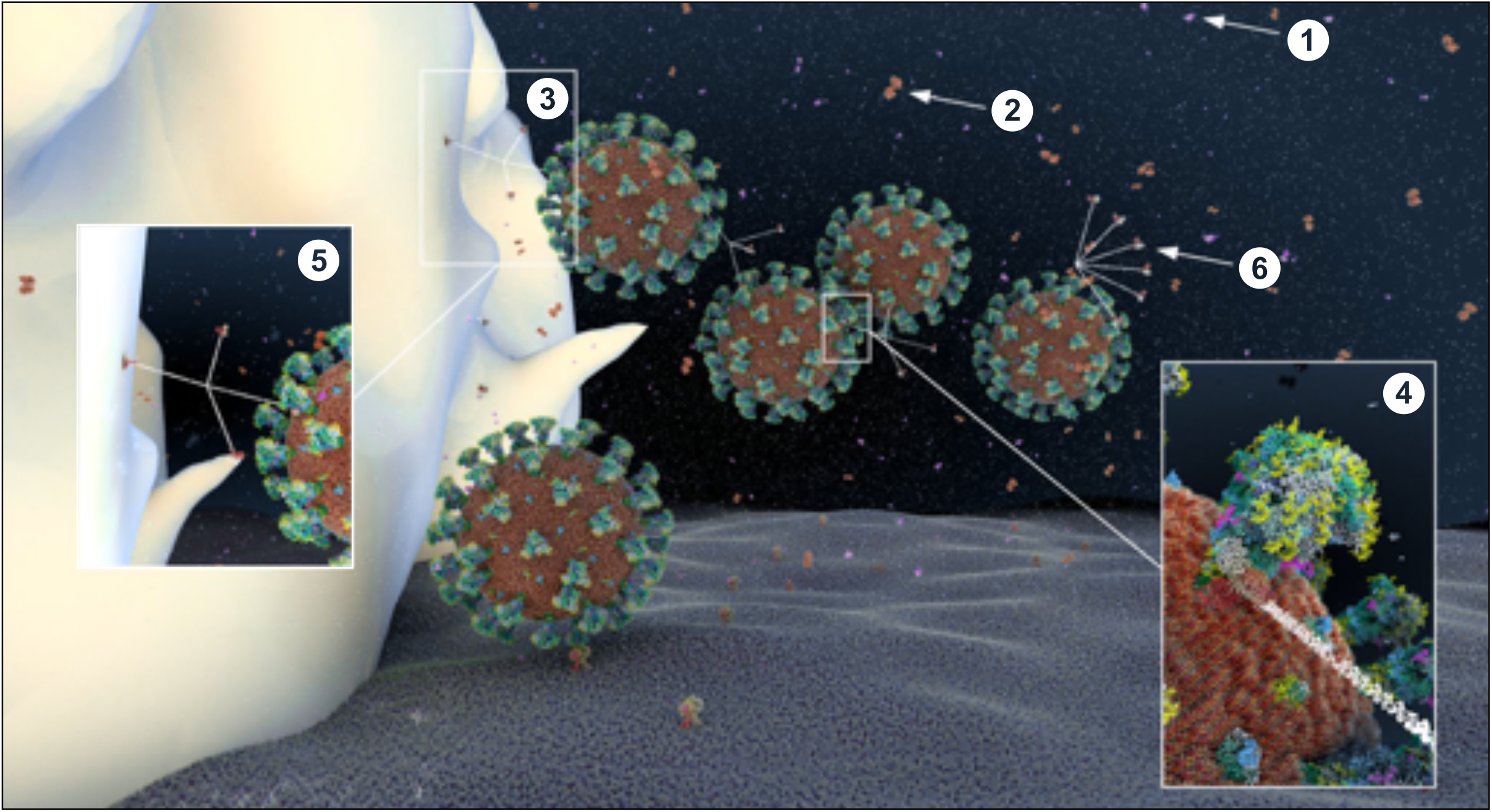
Atomistic reconstruction of SARS-CoV-2 virions in alveolar ASL with normal glucose concentration (0.4mM). In normal conditions, SARS-CoV-2 virions entering in the lung are fought by antimicrobial factors such as β-defensin (1), lactoferrin (2) or trapped by the cruciform shape SP-D collectin (3). The CRD domain of SP-D binds to high-mannose glycans of the spikes (4), trapping viruses in a mesh for alveolar macrophage phagocytosis ((5); receptors expressed on the macrophage surface for collectin recognition is not represented here). Some particles evade the defense system and reach the epithelial surface for ACE2 receptor binding. The fan shape of SP-A collectin (6) is probably not adapted for efficient virus recognition but may participate in apoptotic cell uptake and inflammation resolution (see methods for detailed ID of the components presented).

Importantly, the CRD domain of collectins recognize other varieties of carbohydrates with different affinities (217, 226). C-type lectin with an EPN tripeptide motif on their CRD, such as SP-D, show a high affinity for glucose (227). Hence, at high concentrations, glucose can bind to the CRD domain of these C-type lectin that competitively blocks the viral recognition (142, 228–230). In addition, glucose can indirectly impair ß-defensins and lactoferrins activities (see below on ASL acidosis). Finally, high ASL glucose concentrations could impair not only the activity, but also the secretion of these antimicrobial factors (210) (Figure 15).

###### 2.4.1.2.2 Elevated glucose impairs the cellular immunity in the lung

The alveolar phagocytes play a key role in the non-specific elimination of pathogens as well as in the orchestration of the adaptive immune system through crosstalk. Alveolar macrophages, interstitial macrophages and dendritic cells (DC) are some of the few cell types that reside in healthy airspaces ((231) and Figure 13). In brief, at resting state, alveolar macrophages participate in the homeostasis of the lung, mainly clearing apoptotic cells and recycling surfactant. Upon infection, alveolar macrophages recognize early alarm signals from infected cells, such as elevations in type 1 interferon□gamma (IFN□γ) and pathogen-associated molecular patterns (PAMPs), migrate to the site of infection, and initiate a pro-inflammatory response. These activated macrophages (M1) express various cell surface receptors, including surfactant-CRD receptors or pattern recognition receptors (PRRs), leading to pathogens recognition, phagocytosis and clearance (232, 233). They also secrete reactive oxygen species (ROS) to kill pathogens, as well as pro-inflammatory cytokines necessary for the chemoattraction of additional phagocytes and immune cells that migrate across the epithelium to access the site of infection. Additionally, they facilitate the clearance of infected cells to limit the propagation of the infection. In a second phase, guided by anti-inflammatory cytokines and surfactant proteins (such SP-A and SP-D that play a crucial immunomodulatory function; (234)), macrophages switch to the alternatively activated macrophage (M2) state to begin winding down the inflammatory response, phagocytose apoptotic cells, repair damaged cells, and restore homeostasis (224, 235). Increasing glucose above physiological concentrations is associated with a reduction in the chemotactic migration capacity of neutrophils and in their phagocytotic efficiency (79, 236, 237). Interestingly, aging and hyperglycemia are also two conditions associated with a decreased number of alveolar macrophages and DCs, with altered function of antigen presenting cells (APCs) (238–240), significantly impairing the cellular arm of innate defense.

###### 2.4.1.2.3 Elevated Glucose causes acidosis of the ASL

Regulation of the pH of the ASL, neutral under normal conditions (6.9-7), is also tightly controlled as it may affect the general capability of the innate immune defense of the ASL (202). As previously mentioned, elevation in the glucose concentration in the ASL is countered by apical reuptake by the epithelial cells and rapid metabolism by hexokinases in the glycolytic pathway. One main consequence is the production of lactate, in part released into the ASL (212, 241) through apical monocarboxylate transporters (MCT) that are lactate/H^+^ cotransporters. Secretion of lactate into airway secretions leads to an acidification of the ASL that inhibits numerous pH-dependent antimicrobial agents, such as lysozyme, lactoferrin, ß-defensin and LL-37 (242, 243). Acidic pH could also affect the activity of the surfactant protein (229).

The acidification is normally neutralized by secretion of HCO_3_^-^-rich fluid through CFTR channels (202). However, the accompanying secretion of a HCO_3_^-^-rich fluid leads to an imbalance of ions and water, impacting the ASL osmolarity and volume, resulting in increased viscosity of the ASL fluid, diminished beating of the cilia, and reduced mucociliary clearance of waste and pathogens (202, 244–246) — direct consequences of hyperglycemia (202, 241). Indeed, pathological conditions such DM, aging, and hypertension are associated with impaired mucociliary clearance (247, 248). Acidification additionally impairs immune cell migrations such as neutrophil chemotaxis and consequently the efficacy of innate phagocytosis (249).

###### 2.4.1.2.4 Elevated glucose leads to increased production of AGEs

In contrast to N-glycosylation, which requires a complex sequence of enzymatic reactions during protein synthesis in the endoplasmic reticulum (ER) and Golgi apparatus, advanced glycation end products (AGEs) are proteins and lipids modified from a non-enzymatic covalent linking through direct exposure to high amount of sugars (glucose, fructose and derivatives) (250). Glycation of proteins can interfere with their normal functions by disrupting molecular conformational changes, altering enzymatic activity, impeding protein-protein interactions and functioning of receptors. The normal physiological rate of AGE production is markedly increased in hyperglycemia (caused by diabetes for example, (251)), but also increases with advancing age, oxidative stress, and inflammation (252–256).

The presence of high concentrations of glucose in the ASL changes its overall glycation profiling with an increased expression of AGEs, leading to serious consequences for ASL function. First, the activity of lysozymes and lactoferrins, the most abundant antimicrobial peptides in the ASL (215), is significantly reduced (257, 258). Second, AGEs are known ligands for RAGE (receptors for AGEs) that are highly expressed in lung tissue, such as in AECI and AECII cells. RAGE is a pro-inflammatory mediator, with its main role being the amplification of the cellular inflammatory response by producing reactive oxygen species (ROS) (259). Hence, the presence of excessive AGEs in the ASL would lead to a pro-inflammatory status of the pulmonary epithelial cells. Indeed, RAGE was shown to be an important factor in respiratory viral infection, as RAGE-/- mice showed delayed mortality and accelerated viral clearance upon influenza A virus (IAV) infection (260). Finally, we hypothesize that the properties of mucins, highly glycosylated proteins, may also be altered by excessive glycation, leading to a disturbance of the mucus viscosity (261), affecting the efficacy of the mucociliary clearance.

In summary, high glucose in the ASL is associated with the impairment of multiple aspects of the innate antiviral defense of the lung, including the mucociliary clearance capacity, the lectin-mediated recognition of the virus, the general activity of the antimicrobial agents, as well as the number, the migration capacity and the function of the resident neutrophils and macrophages (Figure 15). Taken together, the overall efficiency of the early phase of viral elimination and clearance of infected cells could be seriously compromised by elevations of glucose in the ASL. The integrity of this early non-pathogen-specific phase is critical because if the virus breaks through these defenses, cascades of other pathogen-specific effects are initiated that make it increasingly more difficult for the immune system to protect the body from the virus, especially if it is a novel virus, as is the case of SARS-CoV-2.

#### 2.4.2 Modeling of ASL glucose concentrations in patients at-risk

As mentioned before, defects in tight junction permeability or hyperglycemia could both lead to a rise of glucose in the ASL, with the greatest effect when they coexist. Hence, DM, obesity or acute hyperglycemia, are pathological conditions known to induce an increased concentration of glucose in the ASL (119, 210, 262). For example, ASL glucose is reported 1.2 (+/-0.7) mmol/L in diabetic patients compared to 0.4 (+/-0.2) mmol/L in non-diabetic (263). Concerning epithelial permeability, a defect in tight junction resistance can be induced by exposure to toxic particles from air pollution or smoking, (264, 265), but also, and especially, by chronic inflammatory conditions associated with chronic lung diseases such as cystic fibrosis (CF), chronic obstructive pulmonary disease (COPD) or severe asthma (119, 265, 266). In such inflammatory conditions, glucose in the ASL has been reported to reach 1.6 (+/-0,1) mmol/L, or even 2 (+/-1.1) mmol/L depending on the pathology (211). Diabetic patients not only suffer from hyperglycemia, but they also often present with chronic inflammation (247, 267), aggravating the disruption of glucose flux from the blood to the ASL.

To attempt to quantitatively evaluate to what extent changes in blood glucose can change glucose levels in the ASL under various permeabilities of the tight junctions, we produced a computational model using data obtained from the literature (see Methods and Figure 17) and used this model to estimate the ASL glucose concentration for a control case (with normal blood glucose and epithelial resistance (Rt)) and a diabetic case (hyperglycemic and impaired Rt) (Figure 17). The model accurately reproduced the values of ASL glucose reported (263) for a control case (0.6 mmol/L versus 0.4 (+/− 0.2) mmol/L reported) and a diabetic case (1.6 mmol/L versus 1.2 (+/− 0.7) mmol/L reported). The model suggests, that even moderate increases in blood glucose, if combined with any impairment in paracellular lung permeability (impaired Rt), could lead to large increases in ASL glucose concentrations.

**Figure 17:**
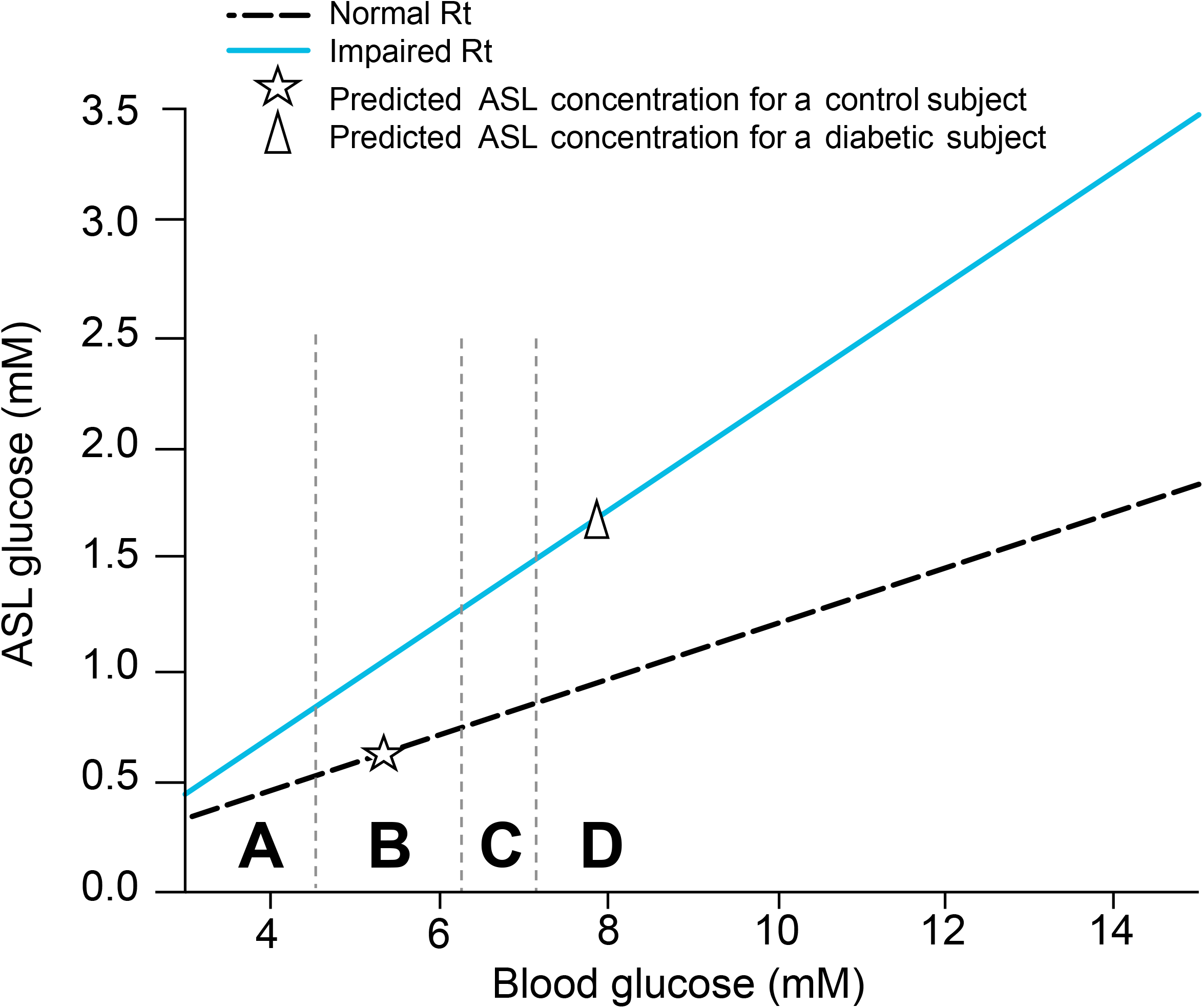
Simplified model of the combined effect of blood glucose concentration and tight junction permeability (normal or impaired Rt) on ASL glucose concentration. The model predicts the trajectory of glucose in the ASL in function of the concentration of glucose in the blood, in case of normal (dashed line), or impaired (blue line) epithelial resistance (Rt), such as reported in diabetic patients (see method). A = hypoglycemia, B = normal FPG, C = IFG, D = acute hyperglycemia or diabetes. star = predicted value of ASL glucose for a control subject, triangle = predicted value of ASL glucose for a diabetic subject.

Then, we used the model to predict ASL glucose concentration in the other group at-risk for COVID-19, for which there is no data available in the literature.

Aging is a condition that, in addition to reduced glucose metabolic capacity, is strongly linked to a general decrease in paracellular resistance in many tissues, including the lungs (268). We therefore used the model to infer an age-related increase of FPG (Figure 8A) based on reported increases in the paracellular permeability with aging (Figure 17). Indeed, the model predicts that the glucose concentration in ASL increases significantly with age, as expected, because FPG increases and epithelial resistance decreases with age.

Hypertension is associated with chronic inflammation (269), which could also be responsible for a general impairment of cellular epithelial resistance. We reviewed above how hypertension is linked to an increased FPG and a higher risk of developing IGT. Based on these known qualitative effects and the quantitative modeling, it is reasonable to assume that people with hypertension will also present with higher concentrations of glucose in their ASL. Importantly, higher glucose in the airway secretions has been observed in ventilated patients in the ICU (262, 270), not surprisingly correlated with stress hyperglycemia, and not necessarily only in those patients with a chronically compromised glucose metabolism. Hence, it is most likely that all groups defined at risk for COVID-19, present with a higher concentration of glucose in their ASL as summarized in Figure 18.

**Figure 18:**
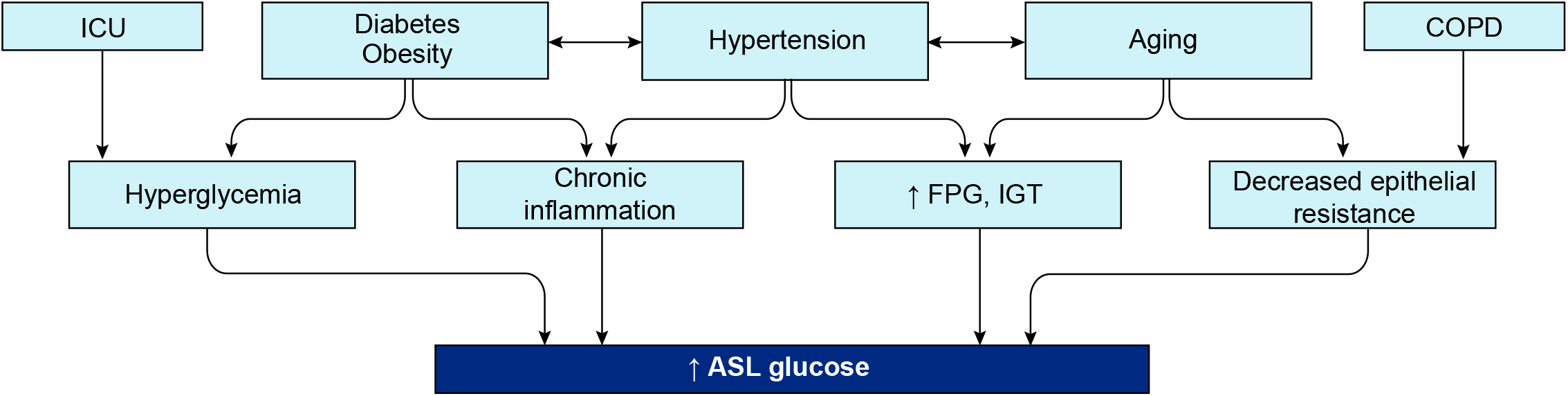
Mechanisms involved in increased ASL glucose concentration in the groups at risk for COVID-19.

It is also important to emphasize that viral infection itself is a condition known to affect the tight junction resistance (271), which would act synergistically to facilitate the infection if the virus breaks through the primary defenses of the lungs.

### 2.5 The multiple effects of glucose on SARS-CoV-2 infection

#### 2.5.1 Infection in healthy patients

In brief, when a healthy person becomes infected, droplets containing virions that reach the respiratory tract will activate the mucociliary clearance as well as a reflex cough to normally expel the virus. If some virions reach the deeper airways and alveoli, they become trapped and inactivated by a second layer of defense, namely the humoral defense of the ASL, composed of numerous antimicrobial peptides, such as the lactoferrin, ß-defensins or the SP-D proteins (see Figure 16), as previously mentioned. Beyond this barrier, those cells that do get infected quickly produce pro-inflammatory cytokines and type 1 IFN to alert the immune system and the neighbor’s healthy cells to protect themselves. The resident macrophages and DC cells, also activated by PAMPs, convert into the M1 phenotype and promptly release proinflammatory molecules, such as type 1 IFN, TNF-a and Il-1b (272), as well as a panel of chemokines to attract and activate more resident and circulating phagocytes and immune cells (neutrophils, DC cells and monocytes, as well as cytotoxic NK cells). SP-D proteins help drive the phagocytosis of the virus by the alveolar macrophages that also produce ROS to help clear the virus and the infected cells. Once at the infected site, immune cells themselves release a battery of pro-inflammatory and anti-inflammatory cytokines as well as ROS to further orchestrate an even more elaborate immune response (273). The infected AECII cells express damage-associated molecular patterns (DAMPs) on their plasma membrane, produce ROS and cytokines to activate their own phagocytosis and clearance by macrophages, converted to the M2 phenotype - all to limit viral propagation to the neighboring cells (273). AECII cells also secrete more surfactant proteins to further amplify the local innate defense and help drive the resolution of the inflammation through their immunomodulatory activity and their capacity to stimulate phagocytosis of apoptotic cells (219, 224). Viral replication is contained by this timely orchestration of non-pathogen specific humoral and cellular innate pulmonary defenses (274) and is therefore a determining step to avoid a deeper infection (192). Complete viral clearance is finally achieved through the adaptive immune response orchestrated by the T and B lymphocytes coming from the bloodstream (68), reaching the site of infection by diapedesis: but of course, even more effectively if the body has previously been exposed to the pathogen.

#### 2.5.2 Elevated glucose favors the primary infection and viral replication

##### 2.5.2.1 Elevated glucose impairs the primary non-specific defense of the ASL

We reviewed and showed above that all groups at high risk for COVID-19 are likely to present with higher glucose in their ASL (see Figure 18), which acts to impair numerous facets of the primary innate humoral and cellular defenses (detailed in Figure 15). Reduced capacity of the early innate immune response and in consequence reduced physical viral clearance by glucose in these patients may explain the general increased susceptibility to infection with respiratory viruses such as SARS-CoV and influenzae (see Figure 19). Using the BioExplorer, we have produced a movie showing the main impacts of high glucose in ASL on the primary step of infections in the lung, to explain the increased susceptibility to respiratory viruses in at-risk patients (https://www.youtube.com/watch?v=hkgqG0nzW9I).

**Figure 19:**
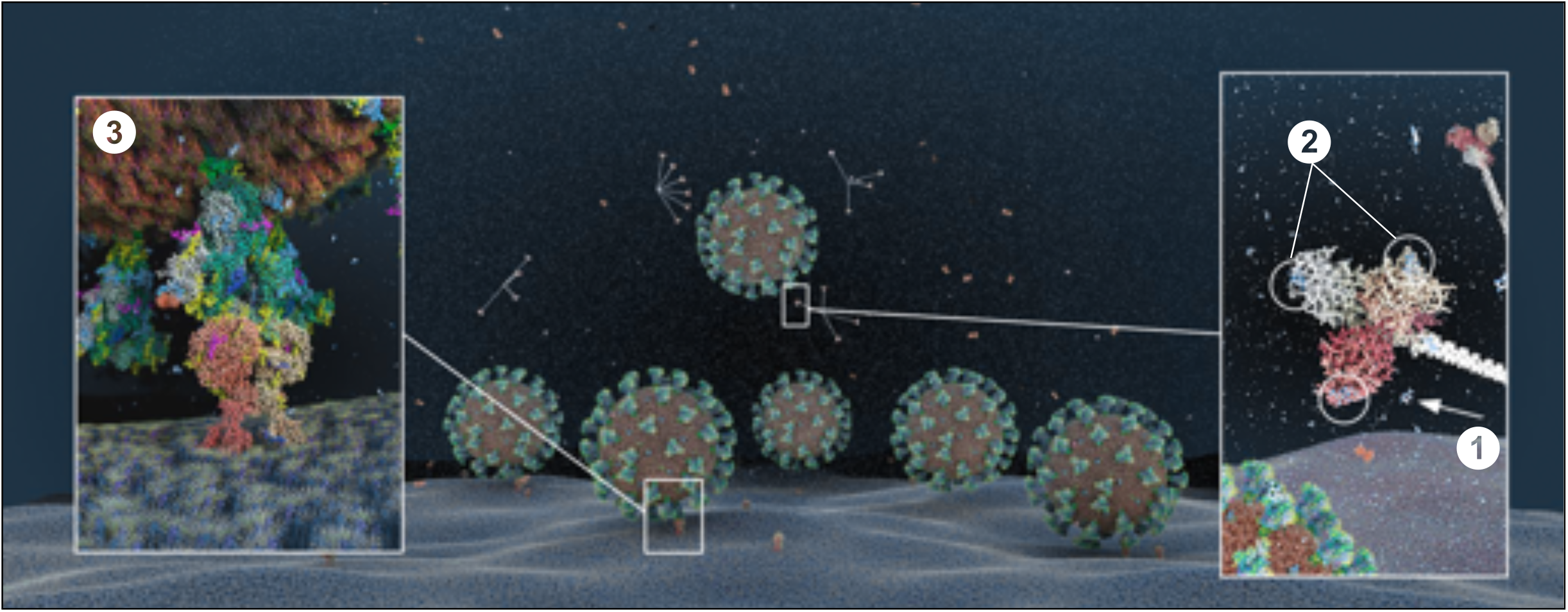
Atomistic reconstruction of SARS-CoV-2 virions in ASL with high glucose concentration (1.2 mM). In patients with high glucose in the ASL, the concentration of antibacterial agents (lactoferrin, β-defensin) is decreased. Glucose molecules (1) in excess bind to the CRD domain of the SP-D heads, ((2) two glucose molecules may bind each head of the trimer), competing for virus recognition. More virions reach the surface of the epithelial cells to bind their receptor ACE2 (3) for further endocytosis (see Methods for detailed ID of the components presented). As the number and migration capacity of alveolar macrophages are decreased in high glucose, we did not include the macrophage in the reconstruction.

##### 2.5.2.2 Elevated glucose levels facilitate ACE2 binding and cell entry

When the virus reaches the receptor, the RBD domain of the spike must move into the “up-conformation” to bind to the receptor, which triggers a sequence of cleavages by host proteases (membrane TMPRSS2 and furin) required for the virus to fuse with the host cell membrane and enter the cell. The virus must then strip off its coat to deliver the mRNA package inside the cell. We have described how elevated blood glucose can elevate glucose in the ASL and cause acidification of the ASL. Additionally, a study has shown that a mere bolus of glucose can also produce a more acidic intracellular environment (143, 279). A lower pH in the ASL is thought to help the SARS-CoV-2 spike conformational masking to avoid detection by the immune system while it binds to the receptor (275). According to the literature, we speculate that more acidic pH in the ASL increases proteases activity (246), that would facilitate the membrane fusion (276–278). In addition, lowered pH intracellularly could further help the uncoating of the virus (143–279). Taken together, elevated blood glucose can not only compromise the early physical and immune barriers making it easier for the virus to reach its target receptor, but can also facilitate all the main steps of the actual process of infecting the cells, such as receptor binding, membrane fusion and cell entry.

##### 2.5.2.3 Elevated glucose favors viral replication

###### 2.5.2.3.1 Increased glycolysis rate

Enveloped viruses have evolved the ability to reprogram carbon metabolism of cells and hijack the glycolysis pathway for their own replication (137, 144). As mentioned above, in the case of high glucose in the ASL, the epithelial cells have the capacity to uptake this glucose from the apical side to keep concentration low in the ASL. However, glucose levels (for example blood glucose reaching > 6.7-9.7 mmol/L) can exceed the capacity for re-uptake (119). As a consequence, not only the glucose concentration in the airway, but also the glucose level inside the cells rises, saturating the hexokinase capacity for glucose phosphorylation. This negative feedback could lead to an abrupt runaway elevation of glucose levels in the ASL as the increased concentration of unmetabolized glucose inside the cells lowers the driving force for the reuptake of glucose from ASL. Thus, once inside the cell, the virus has access to an abundant supply of glucose for producing the nucleotides, amino acids, lipids and the ATP needed for replication (145). Therefore, not only does elevated glucose allow more viral particles to access and enter the cells, but also provides an ideal environment for efficient and fast intracellular replication. This analysis is in agreement with a recent study showing that the use of the glucose analogue 2-deoxy-D-glucose blocks SARS-CoV-2 replication in Caco-2 cells (280).

###### 2.5.2.3.2 Efficient glycosylation process

A specific glycosylation coating is required for viruses to efficiently evade the immune system and invade cells (see above and (167)). Enveloped viruses have the capability to hijack the host cell N-glycosylation machinery to adorn their own glycoproteins with host glycans; however, to do so, a readily available glucose supply is essential. Glycosylation of some of the host proteins, such as ACE2 are also essential to allow more viruses to enter (170, 181). A large supply of intracellular glucose therefore provides the ideal environment to ensure maintenance of glycosylation profiles of both viral and host proteins throughout the progression of the infection.

###### 2.5.2.3.3 Exponential viral replication

High viral replication rates result in host cell damage and death, with numerous adverse effects. In the non-alveolar epithelium, damaged ciliated cells lead to reduced mucociliary clearance capacity (281, 282), and compromised integrity of tight junctions increases paracellular flux of glucose into the ASL (119, 207), escalating the damage through the numerous mechanisms described before (see Figure 20). This is in agreement with a recent publication on IAV infection showing that elevated glucose prior to viral infection increases virus-induced pulmonary barrier damage (283). Host cell damage together with elevated glucose turns negative feedback homeostatic mechanisms into a positive feedback loop of pathological processes. Increased production of ROS caused by cellular damages and hypoxia induces expression of the HIF1α (hypoxia inducible factor 1α) transcription factor (145) stimulating the expression of GLUT transporters (211, 212, 284, 285) and glycolytic genes (286, 287), that increase extracellular glucose uptake and glycolytic capacity, amplifying the viral replication (287–290). HIF1α also induces the expression of LDH (286) causing an increase in the conversion of pyruvate into lactate (a biomarker of a poor COVID-19 prognosis), further decreasing the pH, and further compromising innate defenses. Thus, the increased glucose levels in the ASL observed in high-risk patients, not only favors viral access to the cells, receptor binding, cellular entry, and the delivery of its genetic material, but also a vicious cycle of exponential viral replication depicted in Figure 20. This causes significant damage to the local alveolar epithelium and reduced capacity for gaseous exchange, likely correlating with the appearance of respiratory distress symptoms in the patient such as shortness of breath, dyspnea and fatigue.

**Figure 20:**
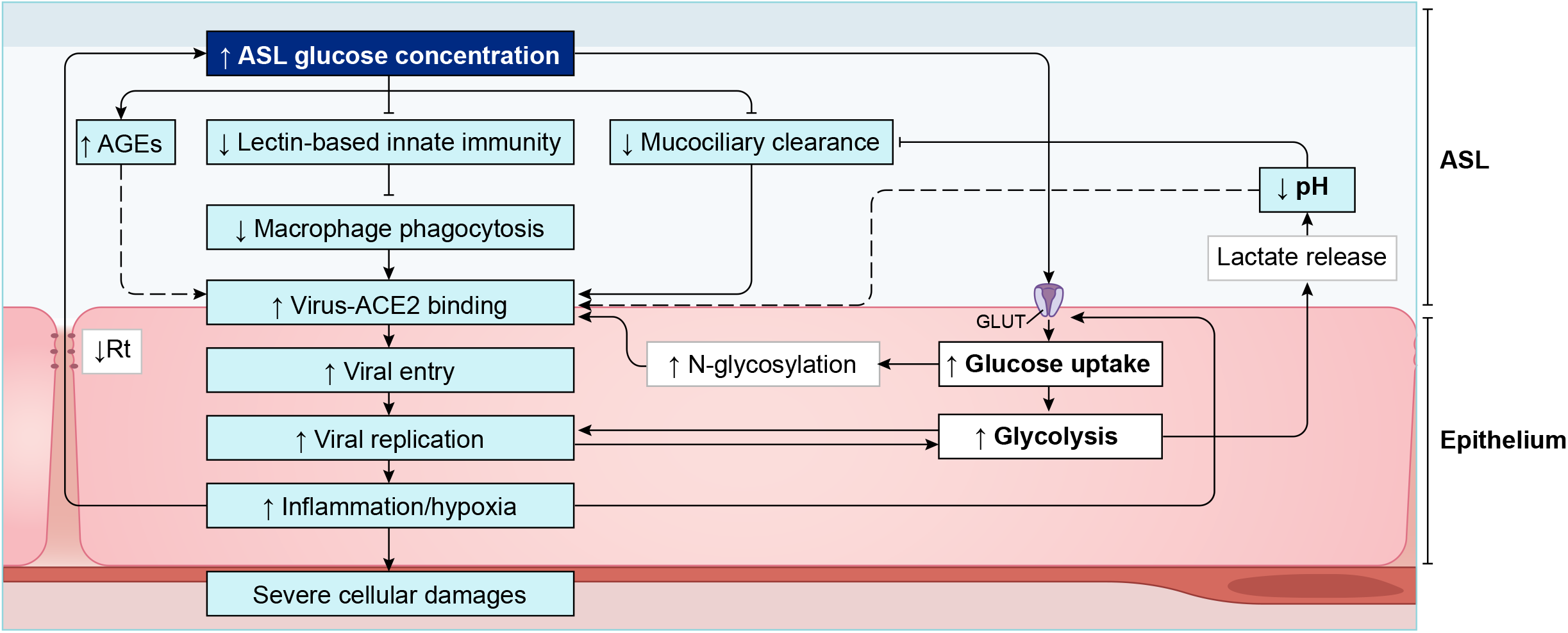
Graphical representation of the effects of high glucose in ASL on SARS-CoV-2 primary infection and viral replication. AGEs = Advanced glycation end products, Rt = epithelial resistance, GLUT = Glucose transporters.

#### 2.5.3 Modeling of the impact of glucose concentration on the different steps of SARS-CoV-2 primary infection

To better understand the interplay between the key variables of the numerous glucose-mediated actions implicated in the SARS-CoV-2 primary infection and to attempt to quantitatively evaluate the impact of elevations in blood glucose levels on COVID-19 severity, we built a computational model to simulate numerous glucose-mediated actions and predict the severity of the infection with different viral loads (see Methods).

##### 2.5.3.1 Modeling of SARS-CoV-2 binding with its receptor ACE2

A schematic of the model is presented in Figure 21A. Briefly, lectin traps a fraction of the virus in the ASL before it reaches the receptors. Higher glucose concentrations act competitively for binding to lectin, leaving less lectin available for trapping the virus and allowing more viruses the chance to reach the receptor. In the presence of a constant concentration of lectin, binding of the virus to the receptor depends on both the viral load and glucose concentration in the ASL. After endocytosis, the virus uses epithelial glucose to replicate, leading to the production of lactate, which when released in the ASL, further lowers the pH of the ASL.

**Figure 21:**
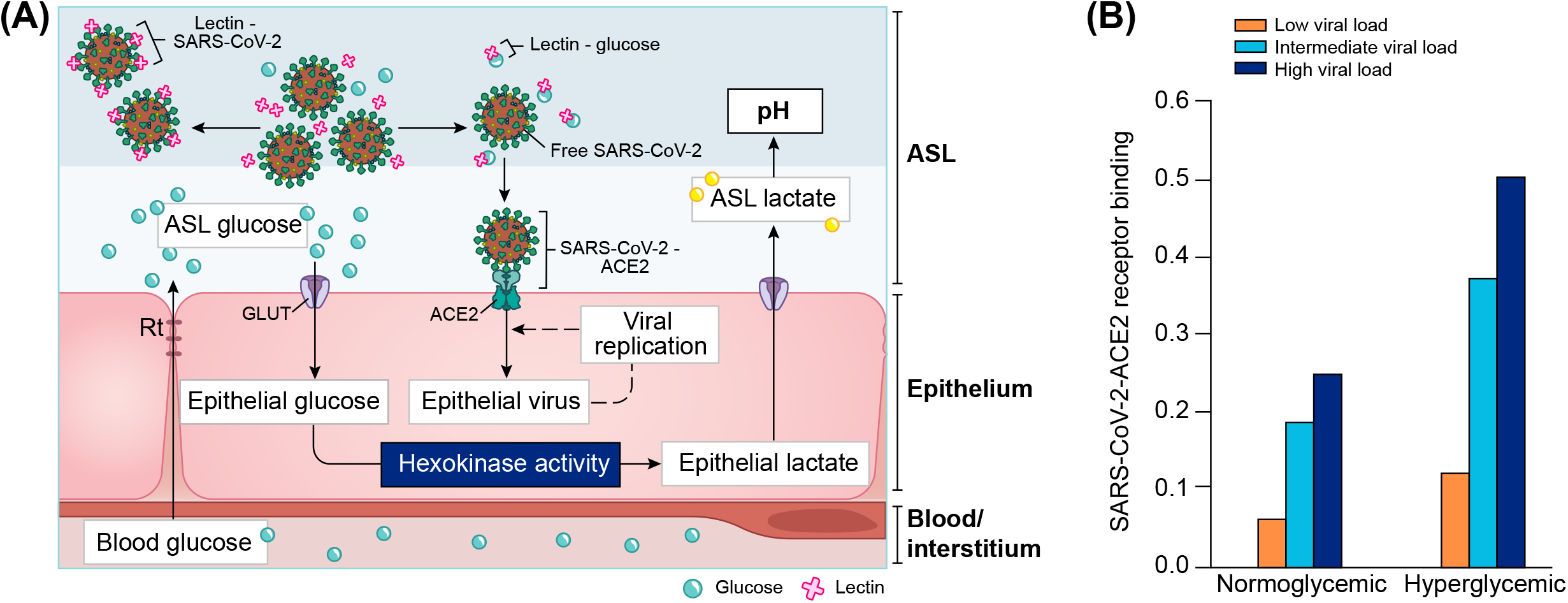
Computational modeling of glucose-dependent SARS-CoV-2 infection. (**A)** Schematic representation of the parameters used in the different SARS-CoV-2 primary infection computational models. GLUT = Glucose transporters (1, 2, 10); Rt = paracellular resistivity (1/Rt = paracellular conductivity in model) (see methods for details). (**B)** Simplified modeling of SARS-CoV-2 - ACE2 binding in a normoglycemic or hyperglycemic patient, as a function of the viral load (represented by three different viral contents at the time of infection, see Definition of viral loads in methods). Normoglycemic = 0.4 mM ASL glucose, hyperglycemic = 1.2 mM ASL glucose.

We first simulated the binding of SARS-CoV-2 to the receptor ACE2 as a function of three different viral loads at the time of infection in a normoglycemic or hyperglycemic condition (See Methods). The model illustrates the extent to which SARS-CoV-2 may bind to ACE2 depending on both the viral load (see methods for description of viral loads) and the glucose concentration in ASL. Increased glucose in the ASL due to hyperglycemia increases receptor binding for all viral loads (Figure 21B). In the hyperglycemic case, binding to the receptor is nearly doubled for all viral loads, suggesting that the receptor binding at any viral load is amplified under hyperglycemic conditions. Additionally, a high viral load results in near maximal receptor binding if patients are hyperglycemic.

##### 2.5.3.2 Simulating SARS-CoV-2 cell entry

We next explored the efficiency of SARS-CoV-2 endocytosis after a simulated “sneeze” that delivers virions in a pulse-like manner with a time course of concentration decay (see Methods). We used three “sneezes” of varying viral content and explored cell-entry after receptor binding; again, in a normoglycemic or hyperglycemic condition. The rise time is a function of inhaling the virus and the decay time represents clearance from the lung (Figure 22A).

**Figure 22:**
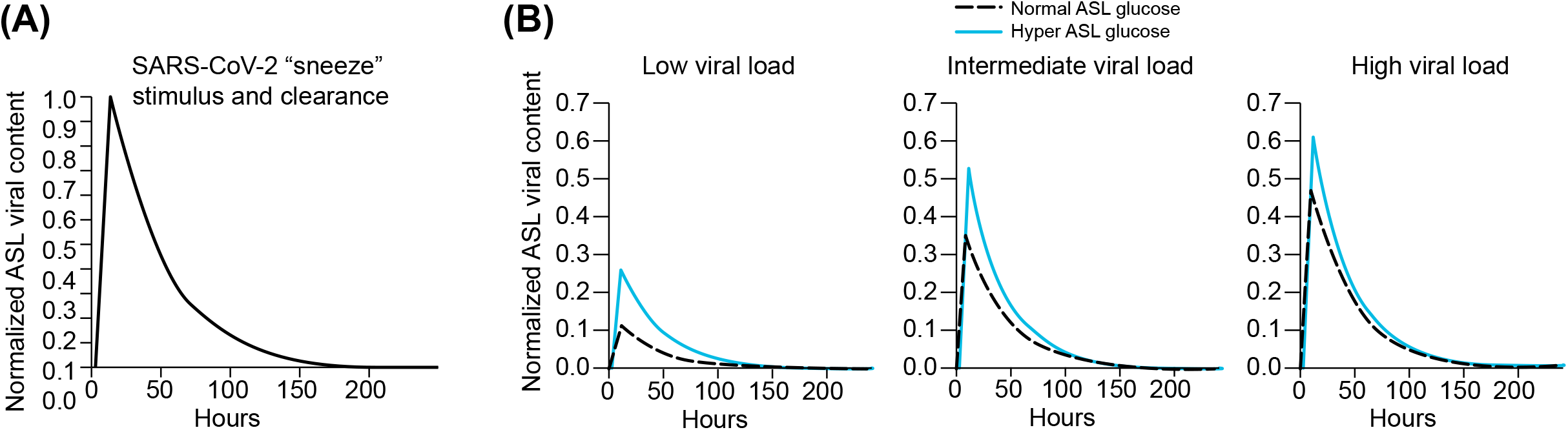
Simplified computational model of SARS-CoV-2 endocytosis as a function of glucose concentration in ASL and viral load at time of a sneeze stimulus. **(A)** Theoretical model of the time course of an arbitrary viral load of SARS-CoV-2 (sneeze stimulus) in the lung epithelial cells. The rise time is a function of inhaling the virus and the decay time represents clearance from the lung (**B)** Simplified model of SARS-CoV-2 endocytosis in function of glucose concentration in the ASL (dashed line = control = 0.4mM, solid blue line = hyperglycemic = 1.2 mM), after infection with a low, intermediate or high viral load of SARS-CoV-2 at time of infection.

The model suggests that SARS-CoV-2 endocytosis in epithelial cells is substantially increased by high glucose in the ASL in all cases, but with the greatest effect seen for low viral loads (Figure 22B, blue lines). The decreasing effect of elevated glucose concentration as the viral load increases (left vs right panels) is due to saturation of the endocytic process.

##### 2.5.3.3 Modeling of SARS-CoV-2 replication rate

In the next step, we examined the viral replication rate which takes into account receptor binding, endocytosis and subsequent intracellular viral load, which depends on glucose concentration and the viral load in the ASL (Figures 23 A,B and Methods). The model suggests that the viral replication rate for low viral loads in the hyperglycemic case is equivalent to the rate of replication induced by high viral loads in the normoglycemic condition. It also suggests that the hyperglycemic condition can further amplify the replication rates induced by any viral load by three to four times compare to normal condition (Figure 23B).

**Figure 23:**
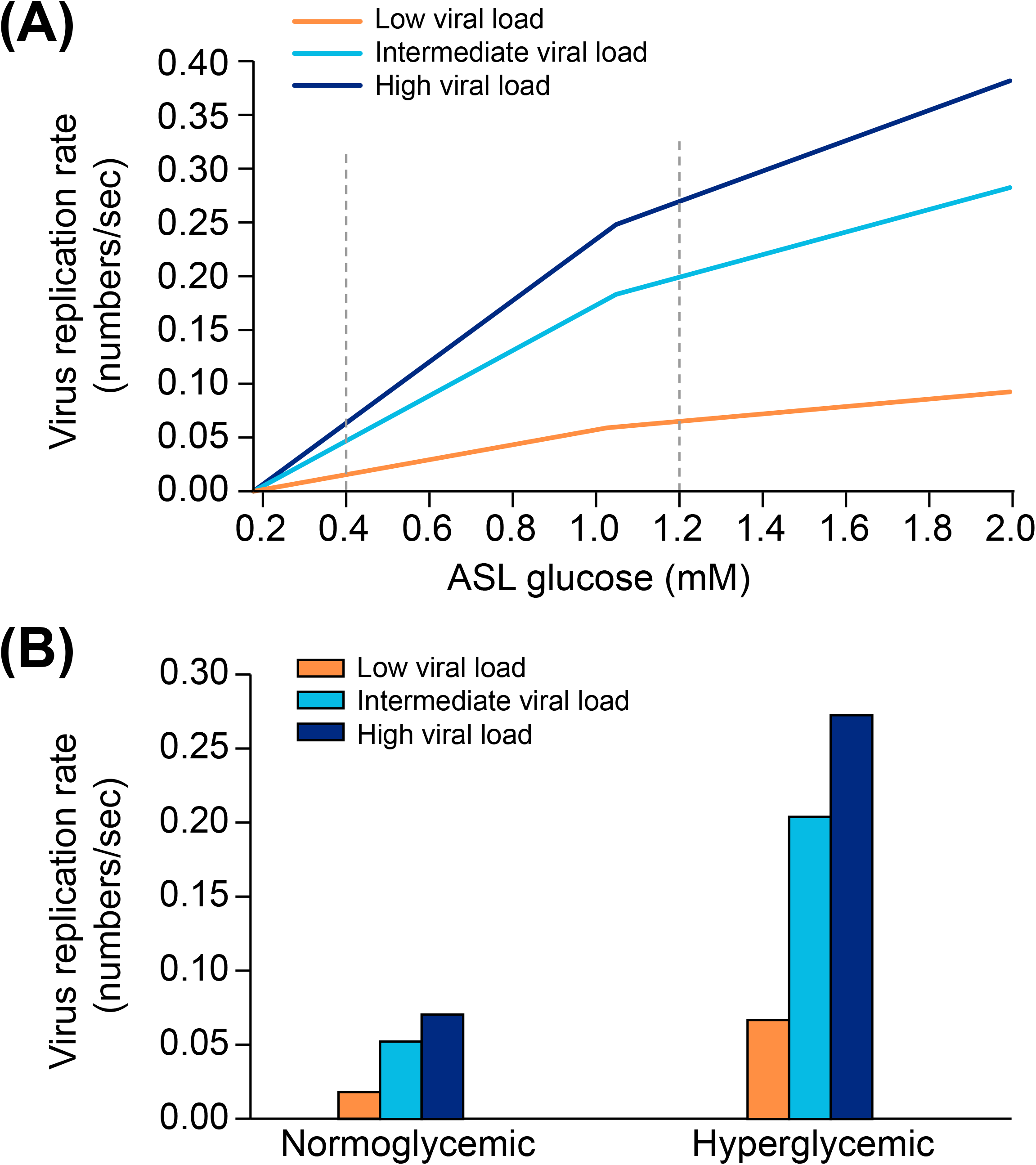
Simplified computational model of SARS-CoV-2 replication rate as a function of glucose concentration in ASL and viral load at time of a sneeze stimulus. (**A)** Dynamic of the viral replication as a function of a sweep over a range of glucose concentrations in ASL. (**B)** Values of viral replication rate in a normoglycemic (0.4 mM ASL glucose) or hyperglycemic case (1.2 mM ASL glucose) after a low, intermediate or high viral load at time of infection (normo- and hyperglycemic values are indicated by the dashed lines in panel A).

##### 2.5.3.4 Modeling SARS-CoV-2 viral numbers

Finally, we simulated the hypothetical viral number produced in epithelial cells after a “sneeze” stimulus which takes into account receptor binding, endocytosis and viral replication rate, again as a function of both glycemic conditions and viral loads (Figure 24A). We then used the resulting value as a biomarker of primary infection severity (Figures 24B and Methods). We took the number of virions in the epithelium for each viral load in a normoglycemic (0.4 mM ASL glucose) and a hyperglycemic condition (1.2 mM ASL glucose), and set a hypothetical threshold for primary infection severity (number of virions generated per epithelial cell that would induce severe epithelial damage) at around 1×10^4^ virions (Figure 24B). While the hypothetical severity threshold was slightly crossed in the normoglycemic condition at the intermediate viral load, condition it was already reached at the low viral load in the hyperglycemic condition. Furthermore, the degree of the effect of hyperglycemia on severity of outcome depends on the viral load, as the severity threshold is only slightly cross in the normoglycemic condition in case of high viral load, whereas it is dramatically exceeded in the hyperglycemic condition already in case of intermediate viral load (Figure 24B).

**Figure 24:**
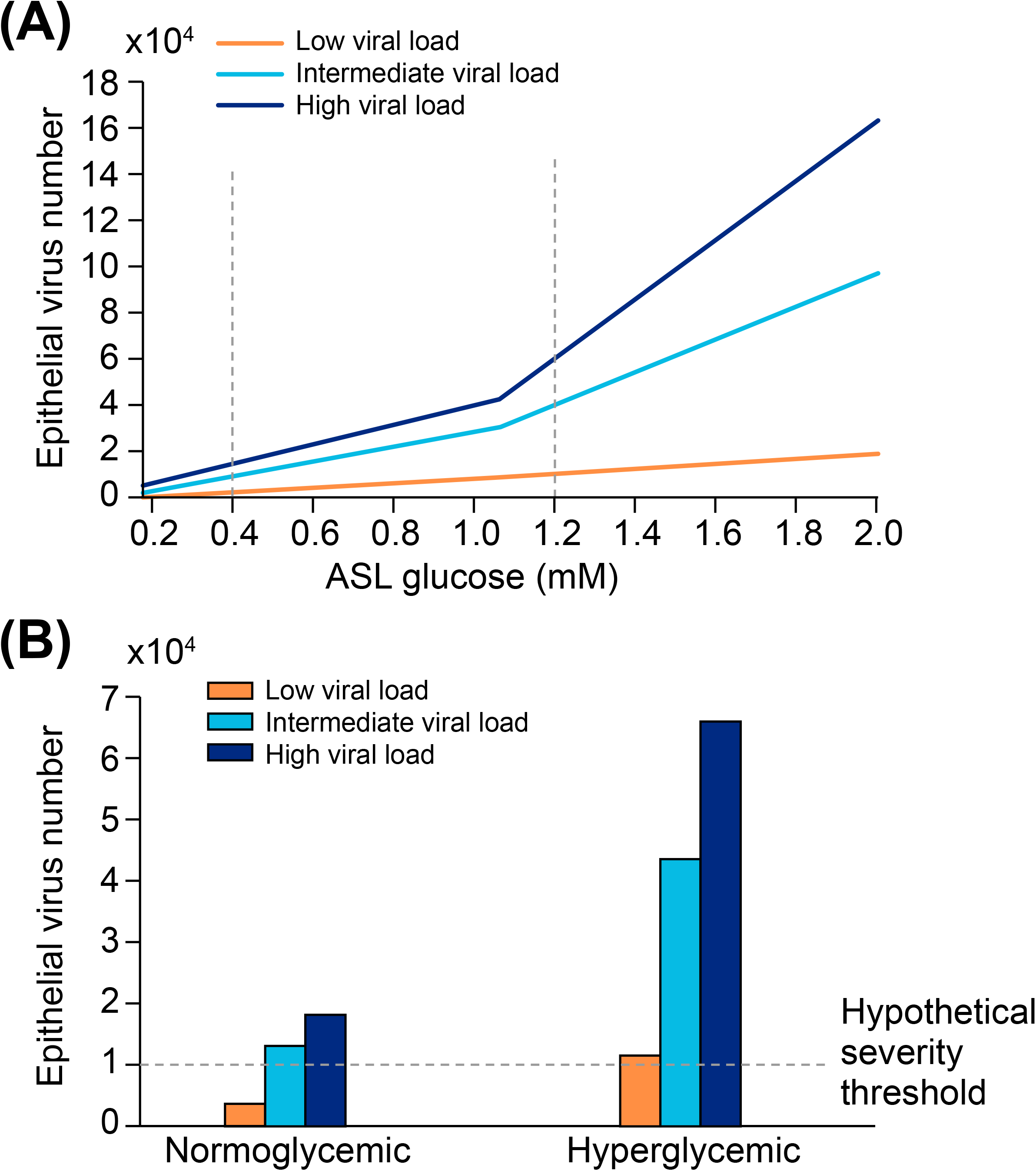
Simplified computational model of the number of virions in the epithelium as a function of glucose concentration in the ASL and viral load at time a sneeze stimulus. (**A)** Dynamic of the viral propagation as a function of a sweep of a range of glucose concentrations in the ASL. (**B)** Number of virions in a normoglycemic (0.4 mM ASL glucose) or hyperglycemic (1.2 mM ASL glucose) case after a low, intermediate or high viral load at time of infection (normo- and hyperglycemic values are indicated by the dashed lines in panel A). We set a hypothetical “severity threshold” for an absolute value >1×10^4^ virions.

To summarize, we modeled the various interactions of glucose in the lung reported in the literature to test the feasibility of the effects of a range of glucose concentration in the ASL on viral receptor binding, endocytosis and replication in lung epithelial cells. The model suggests that while under normal conditions, a high viral load is required to cause severe epithelial damage, under hyperglycemic conditions, even low viral loads could start causing damage. This emphasizes both the importance of the viral load in the infection process and the susceptibility conferred by elevated glucose levels. Consequent viral binding, endocytosis and replication is predicted in hyperglycemic conditions already with intermediate viral loads, and becoming even extreme with high viral loads. While these results could vary somewhat quantitatively due to limitations in the model resulting from simplification steps or unknown mechanisms and parameters, the direction of the results is unlikely to change since all reported effects of glucose only facilitate the infection process.

These first steps of the infection are defining moments for how the disease progresses further. In the next chapter, we show how, if the escalation is not controlled at this stage, high blood glucose also facilitates the subsequent development of the disease and its complications.

#### 2.5.4 Elevated glucose contributes to severe complications of COVID-19

Patients with DM, obesity, hypertension or the elderly are generally more sensitive to respiratory viruses such as influenza or respiratory syncytial viruses (RSV) (291, 292). The common impairment of non-specific innate immune defenses of the lung due to high glucose in ASL that we have previously described, could explain the general susceptibility of these patients to respiratory viruses. However, the reason why SARS-CoV-2 infection leads to a worsening of the disease with severe symptoms, such as ARDS, multi-organ failures, or pulmonary embolism in some cases is still elusive. Specificity of viruses resides, among others, in their binding receptor, i.e. ACE2 in the case of SARS-CoV viruses. ACE2 is an effector of the RAAS system that converts angiotensin II (Ang II) to angiotensin 1-7 (293). Following the binding of SARS-CoV-2 to its receptor, the virus-receptor complex is internalized, leading to the inactivation of ACE2, and consequently to an extracellular increase of Ang II concentration (188, 294, 295) – first locally then systemically. Ang II accumulation has numerous physiological consequences such as an increase in endothelial permeability, vasoconstriction, inflammation and thrombosis (188, 296–298), and further causes glycemic dysregulation and increased insulin resistance (299, 300). Indeed, plasma levels of Ang II are increased in SARS-CoV-2 infected patients compared to healthy individuals, which has been associated with the viral load and lung injury (301), suggesting that inactivation of ACE2 is specifically implicated in the disease severity.

##### 2.5.4.1 Elevated glucose drives the immune response into a cytokine storm and ARDS

As mentioned above, high glucose in the ASL is mainly responsible for a weak migration capacity and activation of the innate immune cells at the site of infection, delaying the secretion of the pro-inflammatory mediators, necessary for a well-timed effective immune response. Indeed, previous studies have shown that hyperglycemia impairs the diapedesis capacity (recruitment from the blood) of immune cells (236, 238, 240, 302) delaying the immune response. This is also in agreement with the delayed recruitment of monocytes and secretion of type 1 IFNs observed in severe cases of SARS-CoV-1 and SARS-CoV-2 infections (303–305). Moreover, it was shown that SARS coronaviruses have evolved an elegant way when infecting host cells, to inhibit their production of type 1 IFNs (306, 307) which are key mediators of the antiviral response, an effect that itself could be explained by high glucose and related increased glycolysis and lactate production by infected cells (308). This overall lag in the pro-inflammatory signal, that could be attributed to elevated glucose consequences, favors viral propagation, leading to greater epithelial damage associated with an increased level of DAMPs, ROS and pro-inflammatory cytokines secretion by the many infected cells. These effects, combined with a late and excessive infiltration of M1 monocytes, M1 macrophages and T cells at the site of infection, cause an exaggerated local inflammation. Indeed, late but excessive infiltration of macrophages, monocytes and neutrophils has been observed in COVID-19 patients (68). Moreover, the subsequent anti-inflammatory signal (M1 to M2 shift) necessary to resolve the inflammation is not triggered on time leaving the immune cells in a pro-inflammatory state. Impaired apoptosis and clearance of infected cells by macrophages also adds to a prolonged secretion of inflammatory cytokines by infected cells— an effect that could be further aggravated by the inhibition of the SP-D and SP-A in elevated glucose conditions and a failure to trigger the clearance of apoptotic cells to resolve the inflammation (219, 224).

According to this cascade, a growing body of evidence suggests an overactivation of many immune cells in hyperglycemic conditions. In high glucose conditions, DC cells, monocytes, M1 macrophages, effector CD4+ and CD8+ T lymphocytes, once recruited, show a hyper-responsiveness with an exaggerated expression of cytokines, mainly IL-6 and IL-1 (145, 236, 238, 240, 302). This exaggerated response is amplified by the SARS-CoV-2 specific inactivation of ACE2 with the resulting local increase in Ang II levels that further aggravates the pro-inflammatory phenotype (IL-6 and ROS (309, 310)).

To recap our analysis, we propose that in hyperglycemic conditions, the local overproduction of ROS and cytokines by infected epithelial cells, combined with the hyperactivation of the immune cells and the general imbalance of the pro- and anti-inflammatory signals, and aggravated by the inactivation of ACE2, most likely lays the foundations for the cytokine storm syndrome (CSS) observed upon SARS-CoV-2 infection in severe cases (68). Importantly, the immune cells themselves express ACE2 and are therefore also targeted by the SARS-CoV-2 virus. Infected circulating immune cells and increased apoptosis of T-lymphocytes can lead to lymphopenia, adding to the overall dysfunction of the immune response (68, 311). To model these conditions, we represented the main events involved in the course of the immune response upon SARS-COV-2 infection in a healthy patient (Figure 25A), and the main impact due to high glucose in high-risk patients (Figure 25B), a representation that is in perfect alignment with other reports (62, 313).

**Figure 25.**
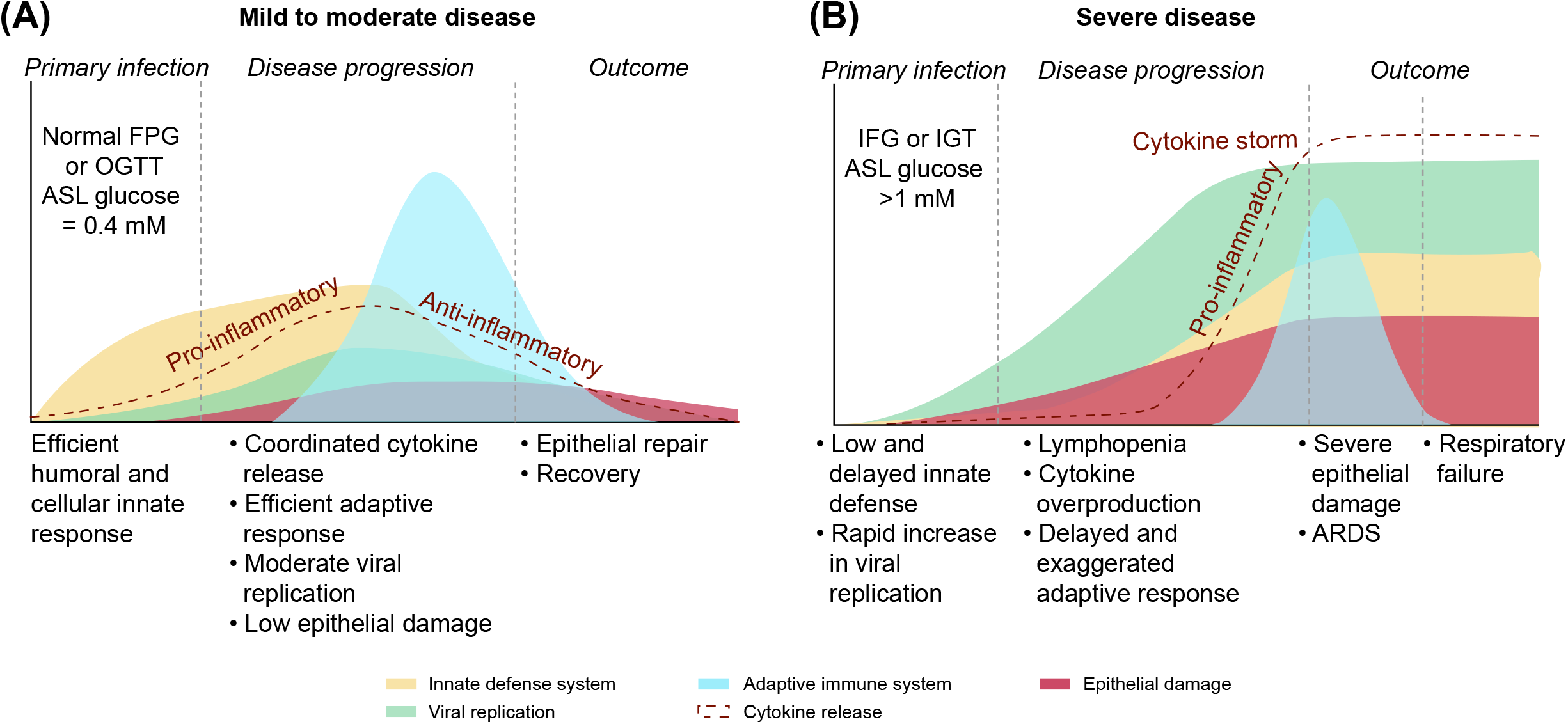
Schematic of the immune response in a patient with normal glycemia or with hyperglycemia or IGT. **(A)** The efficient humoral and cellular innate defense of the lung (antimicrobial agents, phagocytes, DC cells and NK cells) leads to a high phagocytotic rate of the viruses, resulting in a low primary viral replication rate and limiting epithelial damage. The pro-inflammatory signal targeted by the innate immune cells and the infected epithelial cells allows the recruitment of the adaptive immune system (T-cells and B-cells) for further viral clearance. The pro-inflammatory cytokines production is well balanced by the anti-inflammatory secondary response to resolve inflammation. Viruses are finally cleared, and epithelial damages are limited. **(B)** In case of high glucose in ASL due to high blood glucose and/or increased epithelial permeability, the humoral and cellular innate defenses of the lung are strongly impaired, reducing the primary viral clearance and delaying the pro-inflammatory signal. The inefficient primary viral elimination response leads to a rapid increase in viral replication as well as important epithelial cellular damage, combined with a delay in the signal for the adaptive response. Additionally, high blood glucose is linked to exaggerated pro-inflammatory cytokine production as well as an impaired anti-inflammatory secondary response, together responsible for the cytokine storm. The cytokine overproduction and the important cellular damages are leading to development of ARDS and eventually respiratory failure.

With uncontrolled viral propagation and cell damage, the overproduction of cytokines aggravates the damage of the alveolar epithelium and the thin pulmonary vascular endothelium. Excessive fluid accumulates in the alveolar spaces causing pulmonary edema, impairment of gaseous exchange, spiraling into the acute respiratory distress syndrome (ARDS) that is characteristic of the severe forms of COVID-19 (68, 313, 314). At this stage, oxygenation or mechanical ventilation is necessary. ARDS often leads to hypoxemia, respiratory failure and in critical cases, the death of the patient.

ARDS is a severe complication observed also in other respiratory viral infections such as influenza (315). However, the overall incidence of ARDS caused by the seasonal IAV is only around 2.7 cases per 100,000 person-years (316), whereas it is 15%-30% in COVID-19 (317). This huge difference is likely due to the specificity of SARS-CoV-2 for its receptor ACE2 and the consequent higher levels of Ang II. First, as previously described, the Ang II accumulation adds to the uncontrolled pro-inflammatory status, aggravating the cytokine overproduction (IL-6, ROS) characteristic of the cytokine storm. Second, Ang II was shown to inhibit alveolar fluid clearance, to dysregulate ENaC expression that worsens alveolar edema (318). Third, it was shown that Ang II leads to the overexpression of RAGE (319), the pro-inflammatory receptor of AGEs. The higher level of AGEs due to high glucose present in DM, aging or hyperglycemic patients, combined with the Ang II-dependent overexpression of RAGE will lead to a subsequent hyperactivation of the AGEs-RAGE signaling pathway with overproduction of ROS and IL-6 (251, 320), that may add to the sustained pro-inflammation of the lung, responsible for the ARDS.

To summarize thus far, in addition to all the glucose-mediated effects, the specific inactivation of ACE2 may contribute to the higher proportion and more severe form of ARDS upon SARS-CoV-2 infection, compared to other respiratory viral infections, in this group of patients.

##### 2.5.4.2 Elevated glucose contributes to the development of multi-organ failure

ARDS is one of the leading causes of death upon SARS-CoV-2 infection. However, SARS-CoV-2 shows the peculiarity to degenerate into other deadly complications such as multi-organ failures, which has been observed in 47% of the severe cases (35, 39, 40, 321). As mentioned before, the local cytokine storm is responsible for the destruction of both the alveolar and vascular epithelia. Consequently, the inflammatory components present at the infection site (i.e. cytokines, oxidative species, antimicrobials peptides, as well as the virions particles) diffuse and circulate in the bloodstream damaging the vasculature itself and the peripheral organs. Importantly, hyperglycemia, aging and hypertension are all conditions associated with pre-existing endothelium impairments, such as a generalized increase in endothelial permeability (79), further facilitating the transport of these cytotoxic agents from the blood to the peripheral organs. The excessive amount of circulating antimicrobial components, pro-inflammatory cytokines and ROS can cause damage and inflammation to many organs. Additionally, all organs expressing ACE2 become potential additional targets for the circulating SARS-CoV-2 virions such as the heart, kidneys, the gastrointestinal tract, the brain or the vasculature (41, 322). As previously detailed for AECII cells, the binding, replication and dissemination of SARS-CoV-2 may also be facilitated in the peripheral organs in hyperglycemic patients. Indeed, SP-D is expressed and is part of the innate immune defense of other organs such as the gastrointestinal tract and kidneys (172, 226, 323, 324). For this reason, the innate defense of these organs could also be directly impaired by high glucose, consistent with the gastrointestinal symptoms and high rate of kidney failure reported (325, 326).

More importantly, the pancreatic β−cells express significant levels of ACE2 (200) and may become damaged and inflamed upon SARS-CoV-2 infection (327). The dysfunction of the β−cells may cause a reduction in insulin release and secondary acute hyperglycemia, as it was observed in the preceding 2003 SARS-CoV-1 outbreak (328), and more recently also proposed for SARS-CoV-2 (329, 330), aggravating the hyperglycemia and enabling the multiple damaging effects of high blood glucose. Accumulation of Ang II itself can also lead to β−cells apoptosis (331), further amplifying the glycemic dysregulation, insulin resistance, driving a positive feedback paracrine loop and vicious cycle of adverse effects in the disease’s progression.

##### 2.5.4.3 Elevated glucose favors thrombotic events

In addition to pneumonia, ARDS and multi-organs failures, a high proportion of COVID-19 patients were diagnosed with thrombotic events at a much higher rate than in other types of lung infection (332). Indeed, alveolar capillary microthrombi are nine times more frequent in COVID-19 patients than in people with influenza (333), and occur in 80% to 100% of severe cases (334, 335). These patients present blood clots disseminated throughout the lungs associated with elevated levels of thrombotic markers such as fibrinogen, thrombin, plasmin and D-dimer (47, 334). The blood clots are responsible for pulmonary embolism, heart attack and stroke (336), and could also contribute to a dramatic drop in blood oxygen levels in severe cases of COVID-19.

It is reported that this coagulopathy arises from a thrombo-inflammation mechanism (337, 338); the infection and destruction of endothelial cells (ECs) expressing the receptor ACE2 (339, 340) trigger an intricate cascade of inflammatory and pro-coagulant events. Under normal physiological conditions, quiescent endothelial cells (ECs) preserve vascular homeostasis, ensure barrier integrity and function, prevent inflammation, and inhibit coagulation by expressing blood clot-lysing enzymes and producing the glycocalyx, a protective layer of glycoproteins and glycolipids with anticoagulant properties. The infection of the ECs by SARS-CoV-2 is responsible for a massive endothelial pyroptosis, a highly inflammatory form of cell apoptosis (334, 341), associated with the disruption of the glycocalyx, the exposure of the basement membrane and activation of pro-coagulant factors (e.g. P-selectin, von Willebrand factor, leukocyte adhesion molecules and fibrinogen) (342). Additionally, pulmonary ECs are antigen presenting cells and assumed to play a role in the immune surveillance against respiratory pathogens. Consequently, infected ECs also release ROS, proinflammatory chemokines and cytokines such as IL-6 (343). The resulting pulmonary endotheliitis contributes to the innate immune hyperactivation, promotes inflammatory cell infiltration (such as by neutrophils) and exacerbates the cytokine storm (334, 340). Importantly, as previously mentioned, the SARS-CoV-2 invasion also especially provokes the internalization of ACE2. The resulting accumulation of Ang II in the endothelium amplifies the pro-inflammatory status and induces a local pulmonary vasoconstriction that, combined with the pro-coagulant and pro-inflammatory effects of the virus, degenerates to a severe thrombotic phenotype (Figure 26). The alveolar microcirculatory thrombosis may degenerate to a systemic disseminated intravascular coagulopathy, associated with a pro-hemorrhagic pattern that exacerbates organ injury and increases the risk of mortality (333, 338, 339).

**Figure 26.**
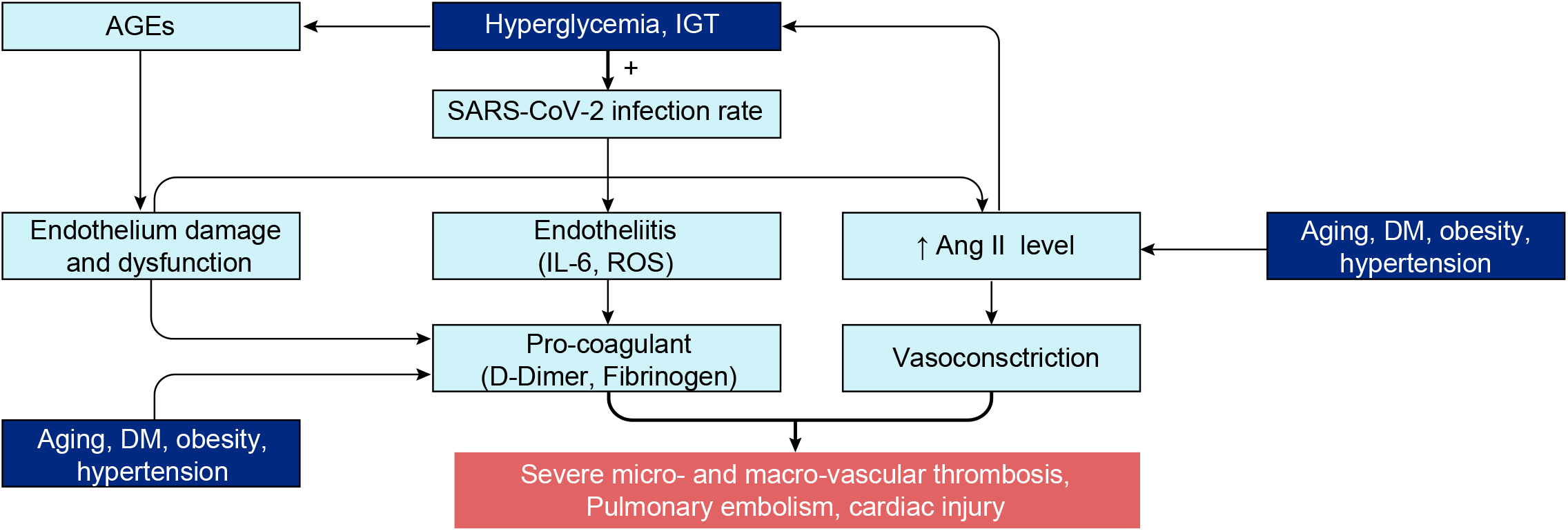
Schematic of SARS-CoV-2 infection in endothelial cells in patients at risk, and the subsequent mechanisms leading to the thrombotic events.

Patients presenting with glucose metabolism dysregulation may particularly be at risk for these thrombotic complications. First, we have highlighted that hyperglycemia or IGT favors a higher viral replication rate, which we assume would also occur in ECs. Second, chronic or acute hyperglycemia is itself a risk factor for coagulation, (344) with increased level of prothrombotic factors and endothelial dysfunctions (e.g. blood viscosity, coagulation and vascular construction) (345–347). Hyperglycemia-induced increase of AGEs, as well as glycation of fibrinogen and collagen, are largely involved in the multiple mechanisms of hyperglycemia-dependent endothelial dysfunction (348–350). Third, increased levels of plasmin fibrinogen or prothrombin was demonstrated in all subsets of patients at risk (347, 351–354). Fourth, aging, hyperglycemia, DM, obesity and especially hypertension, are all conditions associated with a pre-existing increased level of Ang II (299, 355–359). Hence, these patients in particular, may present an excessive concentration of Ang II upon SARS-CoV infection, and then are susceptible to exaggerated vascular vasoconstriction. In addition, the overexpression of Ang II amplifies the glucose dysregulation in these patients, leading to an unstoppable vicious cycle where symptoms become ever more severe.

In summary, patients presenting with a pre-existing procoagulant condition and increased Ang II levels, are more prone to develop severe coagulation disorders and thrombotic events upon SARS-CoV-2 infection, especially if accompanied with hyperglycemia as summarized in Figure 26.

##### 2.5.4.4 Elevated glucose increases the risk of secondary pulmonary infection

Glucose in the ASL is an important and direct nutrient for bacteria or others pathogens (208) and therefore supports secondary bacterial infections, as established in DM (119, 270), cystic fibrosis (360), COPD (361) or patients in the ICU (see Discussion). Secondary bacterial infection during pulmonary disease management is an important cause of mortality, especially in ICUs in European countries (362)). Even if apparently less frequent than in case of IAV infections, bacterial infection is one of the complications in patients critically ill with COVID-19 (363, 364), that could also be linked to elevated blood glucose or IGT in patients at risk.

### 2.6 Therapeutic approaches and research strategies

The evidence that patients with elevated blood glucose or IGT are more prone to severe primary infection and COVID-19 complications and death in the literature is overwhelming. Elevated glucose can not only explain much of the variance in COVID-19 severity as a correlative biomarker, but because virtually every action of glucose in biochemical, metabolic and homeostatic pathways seems to serve only to facilitate the infection, it could also be a primary determining factor in the severity of the disease. Controlling glucose levels could therefore reduce the severity of the disease and consequently also the mortality rate.

Obtaining an unbiased representation of the findings in such a vast database of relevant literature was only possible with the aid of text mining, machine learning and knowledge engineering approaches. The knowledge graph provided an overview of the contents of the dataset, revealed the high-level structure of the information it contains, and helped guide us through and down to the deepest levels of knowledge, where accessing the specific papers referenced allowed human verification of the findings. The hypothesis that arose called for supplementing the review by analyzing data found across different papers, performing computational modeling to test the feasibility of some of the actions of glucose, and producing atomistic reconstructions to better appreciate some of the physical and biophysical parameters involved. This supplemental analysis further supported the hypothesis that elevated glucose is a primary risk factor for the severity of COVID-19.

Interventions that reduce the availability of glucose would allow tackling multiple facets of the primary viral infection: improving the innate defense of the ASL, decreasing the viral replication capacity inside the cells, impairing N-glycosylation process that would compromise the immune evasion facet of the virus and possibly increase the immune recognition, improving timely orchestration of the immune system. Improving glucose metabolism would also diminish the risk of developing secondary SARS-CoV-2 specific complications such as coagulopathy. The literature actually already contains tests of this hypothesis. In fact, the mortality rate is lower among diabetic patients where glycemia is well controlled (82) and recent studies show that patients with uncontrolled hyperglycemia or newly diagnosed DM (i.e. untreated) are even more at risk than those with known DM (i.e. treated) (54, 83). The hypothesis can be refuted, in part or in full, by finding severe COVID-19 cases where comprehensive measurements fail to detect even normally sub-clinical glucose dysregulation.

#### 2.6.1 Glucose lowering drugs

According to the hypothesis that well-controlled glycemia is critical for determining the outcome of COVID-19, the most standard strategy is to use glucose lowering drugs, widely available and low cost. A wide variety of these drugs is available, with different mechanisms of actions that could all present pros and cons for management of COVID-19 (comprehensively reviewed in (86, 365)). For example, ACE agonists would be contra-indicated because they would further increase the inhibition of the ACE pathway and increase expression of ACE2 (366). Also, most of the glucose lowering drugs risk inducing hypoglycemia, which is not recommended. Insulin is widely used for managing glucose levels during hospitalization in ICU, but typically only for diabetic patients (367, 368) where it does seem to reduce mortality, length of stay in ICU, and ventilator dependence with COVID-19 (369). However, its correct adjustment in ICU is extremely challenging (370, 371). Insulin protocols for any COVID-19 patient would therefore have to be explored, developed and clinically tested.

According to the data we gathered for this review, metformin may be an effective glucose-lowering drug for COVID-19. Metformin is an old drug and the first line therapy for diabetes management. Apart from its safer glucose-lowering effect (i.e. reducing glucose without provoking hypoglycemia) metformin has several other interesting effects that may be advantageous for the management of COVID-19. Firstly, metformin is known to restore the permeability function of tight junctions by increasing the transepithelial electrical resistance as well as the expression of the tight junction proteins claudin 1 and occludin (210, 212). Indeed, it has been shown that metformin reduces the airway glucose permeability and the hyperglycemia-induced *Staphylococcus aureus* load, independently of its effect on blood glucose (372). Secondly, this drug has some anti-inflammatory effects, reducing plasma CRP levels, a biomarker for the poor prognosis for the disease (373). Importantly, this anti-inflammatory effect was also observed in the airway epithelium (374). Thirdly, metformin decreases expression of HbA1c and presents cardiovascular protective, vasodilative and anti-thrombotic effects (375–378). Finally, metformin inhibits formation of AGEs ((379, 380) as well as cytosolic and mitochondrial ROS production induced by AGEs in endothelial and smooth muscle cells (381). The combination of these various actions may be protective against the SARS-CoV-2 infection, especially in patients at risk. It is important to mention that metformin is contra-indicated in case of respiratory failure or severe hypoxemia because a side effect, even if rarely reported, is lactic acidosis. Concerning indication of metformin, two studies showed beneficial effects in diabetic users compare to non-users with a reduction in COVID-19 mortality (382) or in heart failure and inflammation (383), which was confirmed by a further meta-analysis (384). However, Cheng et al, 2020 (383) showed that metformin was associated with an increased acidosis. However, they showed that it occurred only in severe cases treated with high dose metformin and in patients that presented with pre-existing renal dysfunction. We found one study where metformin usage seemed to be associated with higher risk of severe COVID-19 in diabetic patients (385), emphasizing that even strong cases for repurposing a drug, such as is the case for metformin, should be performed with great caution and only after extensive randomized clinical trials. More importantly, clinical trials should be also performed on healthy people and non-diabetic cases.

#### 2.6.2 Lowering carbohydrates in diet

The strong link between COVID-19 severity and diabetes and obesity has led to consideration of nutritional interventions in the treatment of the disease (386), as for example the use of low-carbohydrate diets or ketogenic diet (low-carb, high fat diet) The basic principle is to diminish the intake of carbohydrates, providing fat instead of carbohydrates for the body to switch on ketosis and produce ketones as the primary energy source (387, 388). Indeed, there is growing evidence of therapeutic benefits of a ketogenic diet for severe pathologies (389) such as cancer (390, 391), diabetes (392, 393), and pharmaco-resistant epilepsy (394, 395), and for the prevention of Alzheimer disease (396, 397) and other neurodegenerative disorders (398). It is also the first line therapy for the management of the Glut1DS (Glut1 deficiency syndrome) rare disease (399) and there is evidence that a ketogenic diet decreases comorbidities linked to hyperglycemia (400, 401). Since viruses are high glucose consumers, just like cancers (140, 402, 403), diminishing the indispensable primary source of energy for the virus may be an effective intervention. Importantly, it was recently shown to be a safe intervention for patients even in ICU (404, 405).

One study recently showed that the ketogenic diet (KD) activates protective gamma delta T (□δT) cell responses against influenza virus infection (406). These □δT cells are IL-17-producing T cells that play an important antiviral protection in the lung (see Figure 13), maintaining epithelial integrity, regulating homeostasis and providing a first line of defense against pathogens and injury (407). Importantly, □δT cells were shown to be reduced in COVID-19 patients (408). Goldberg et al showed asignificant increase in the frequency and absolute number of □δT cells in the lungs of KD-fed mice; this increase was required for the KD-mediated protection against influenza disease, resulting in lower viral titers and overall better preservation of airway tissue integrity. Interestingly, in 2010, Taylor et al (407) reported that □δT cells were reduced and impaired by hyperglycemia in a mouse model of obesity. In a similar way, KD could also have the potential to help the immune defense against pulmonary viral infection, but this has to be confirmed for SARS-CoV infection.

Supporting this hypothesis, a clinical trial on KD for intubated critical care COVID-19 was initiated (https://clinicaltrials.gov/ct2/show/NCT04358835). The aim of the study was to measure the benefit of KD on gas exchange, inflammation, and duration of mechanical ventilation in intubated patients with COVID-19 infection (on 15 patients at start). Additionally, two recent reports advised on the use of a carbohydrate-restricted diet for the management of the disease (409, 410) and a randomized controlled trial on KD has been developed (see (409)). Cooper et al., claim that the ketogenic diet would be more beneficial than insulin therapy because large fluctuations in blood glucose concentrations are primarily driven by dietary sources, and it would also avoid the adverse effects of hyperinsulinemia. Lowering carbohydrate consumption could therefore manage both hyperglycemia, hyperinsulinemia and may additionally help manage hypertension (411).

#### 2.6.3 Guidelines for COVID-19 biomarkers at admission

##### 2.6.3.1 Importance of systematic glucose metabolism measurement (FPG, PPG, HbA1c, insulin)

The numerous lines of evidence in the literature for elevated blood glucose as a correlative risk factor is overwhelming and makes a strong case for far more thorough monitoring during the management of COVID-19. As previously mentioned, increased FPG becomes an important marker of mortality and morbidity and should be systematically measured. Importantly, a single normal FPG value is not sufficient to exclude acute hyperglycemia or IGT. For this reason, regular FPG and PPG measurements should be systematically obtained. We emphasize that the 2h OGTT test is not recommended as it requires the ingestion of a high amount of glucose, that could be detrimental and escalate the disease progression. We propose that measuring HbA1c and insulin should also be included to detect any glucose metabolism dysregulation (e.g. diabetes, prediabetes, hyperglycemia, IGT, IFG, hyperinsulinemia). Indeed, it appears that the proportion of undiagnosed prediabetic patients is high in severe cases (329). On this line, HbA1c was recently found as a predictor of COVID-19 severity (412). Finally, even moderate dysregulation, and not only severe hyperglycemia, should be taken into consideration as it could be the starting point for an unstoppable viral infection.

##### 2.6.3.2 Alternative biomarkers

###### 2.6.3.2.1 Ang II plasma level

Ang II is strongly associated with dysglycemia, and patients who are at risk for severe COVID-19 disease (e.g. diabetic, obese, elderly or hypertensive) present with an increased basal plasma level of Ang II, that is further amplified by SARS-CoV-2 infection. Ang II possesses inflammatory and vasoconstrictive effects that appear to play a critical role in the COVID-19 disease severity. Hence, measuring Ang II at admission or/and during the course of the disease, could be an additional biomarker for risk stratification or as a prognostic indicator.

###### 2.6.3.2.2 SP-D plasma level

SP-D is a key player in the development and regulation of the innate immune defense of the lung against SARS infection (219, 225). Serum levels of SP-D are elevated in patients with SARS related pneumonia and has been suggested as a marker of alveolar damage in this condition (413). SP-D is mainly synthetized by AECII cells of the lung and is released into the blood during certain types of lung injury. Furthermore SP-D plasma level is considered to be a putative biomarker for pulmonary disease (324, 414), such as acute airway inflammation (415), or exacerbation of COPD (416). Importantly, it was also described as a biomarker of cardiovascular diseases (417, 418) or atherosclerosis (324). Hence, comparing SP-D plasma level in non-critical versus critical patients could indicate if SP-D could be an additional biomarker of COVID-19 disease severity.

##### 2.6.3.3 Glucose management in ICU

The high glucose content in parenteral feeding used in some ICU cases could be more detrimental than beneficial for COVID-19 patients since high glucose favors all stages of the infection. The accepted range of FPG in ICU is (1.45-2g/L; (73)) which is much higher than the normal range. In fact, the common thinking is that “high blood glucose concentrations are believed to be a normal physiologic reaction in stressed patients and that excess glucose is necessary to support the energy needs of glucose-dependent organs.” This strategy may be indicated for some other diseases, but is not supported for SARS-CoV and other viral infections.

The exact target of blood glucose concentration in ICU remains a matter of debate as reports yield contradictory conclusions (419), mostly due to the heterogeneity of the studies, but the beneficial effects of lower glucose in parenteral feeding is consistently supported by multiple studies. For example, Patino et al, (420) have demonstrated that patients receiving hypoenergetic–hyperproteic total parenteral nutrition regimens on a surgical ICU have a more physiological clinical course, with less metabolic stress than those receiving high-energy loads. Later in 2001, (368) Van den Berghe et al, reported that intensive insulin therapy, to maintain blood glucose at or below 110 mg per deciliter, reduces morbidity and mortality among critically ill patients in the surgical intensive care unit. Hypoglycemia has to be avoided, however, in the case of SARS-CoV-2 infection, a tight control of glucose metabolism should be mandatory for ICU patients, as recently proposed (369, 370). Although it is challenging to manage glucose levels in ICU patients, some effective protocols for tight glucose control in these conditions are emerging (88).

## 3 DISCUSSION

CORD-19, a valuable literature dataset, was made open-access to stimulate collaboration and accelerate solutions in the global crisis caused by the SARS-CoV-2 virus. While there are numerous questions one could ask using such a large dataset, we chose to ask why some people get more affected than others. It is difficult, if not impossible, for any expert to synthesize the multifaceted expertise across numerous disciplines contained in hundreds of thousands of scientific studies. We therefore developed machine-learning models to mine the dataset and constructed knowledge graphs to help navigate this literature resource. A simple analysis of the entities mentioned in the papers revealed glucose as one of the biological variables that was most frequently mentioned. We then constructed specific knowledge graphs to focus on all findings that consider glucose in the context of respiratory diseases, coronaviruses, and COVID-19. This allowed us to explore the potential role of glucose across many levels, from the most superficial symptomatic associations to the deepest biochemical mechanisms implicated in the disease. We chose not to construct knowledge graphs to directly reveal any specific type of association(s) between entities to avoid biasing our search. Navigating through all known stages of the disease and biological levels where some association glucose was detected allowed identifying and retrieving the papers and construction of this review. While we cannot completely exclude that some studies have been overlooked, findings from most of the relevant studies are represented in this review.

We also used the knowledge graphs to find the stronger and more consistent claims, tested the feasibility of the parameters reported in multiple papers using computational modeling, and attempted to obtain an atom-level realistic view of the virus and some of the compounds it interacts with. We conclude that the literature strongly supports a case for compromised glucose metabolism that causes elevation of glucose levels in tissue, blood and extracellular fluids as a single pathology that can facilitate virtually every step in the life-cycle of SARS-CoV-2, and that induced elevations of glucose by stress during hospitalization, treatment drugs, and in intravenous infusions can contribute to disease severity. Reduced glucose metabolic capacity could be a common pathology that can contribute to age-dependency of the disease and explain why the specific comorbidities render these groups more vulnerable to the infection. Subclinical pathology of glucose metabolism may also be one of the reasons why some young and apparently healthy people can contract a more severe form of the disease. On the other hand, high glucose metabolic capacity can explain, at least in part, why the youth are less affected and why some elderly can experience a milder form of the disease. Elevated glucose naturally does not act alone. It acts in concert with numerous other pathophysiological pathways to facilitate the primary infection and replication of SARS-CoV-2. For example, the effects of elevated glucose can act synergistically with the virus’s inactivation of the ACE2 receptor to drive a more severe form of COVID-19. Indeed, hyperglycemia or impaired glucose tolerance can cause multiple physiological disturbances that are all linked to the severity of the disease such as an impaired innate immune system, impaired lung epithelial resistance, subsequent hyperactivation and dysregulation of the immune system, increased vascular permeability, and a procoagulant state. Patients presenting with compromised glucose metabolic capacity struggle to contain and eliminate the virus and to prevent the progression of the infection and the occurrence of complications. Patients with high glucose metabolic capacity such as healthy people at any age and in particular young people, have primary lung defenses that are sufficient to contain and expel the virus before significant infection, slow the infection of and replication in cells, and lower the risk of fatal complications. The effects of elevated blood glucose can also act synergistically if lung epithelial tight junctions are compromised causing a positive feedback in the increase of glucose in the ASL, a disruption of glucose homeostasis and a subsequent breakdown of the lung primary defenses.

While there is some evidence for lower expression levels of ACE2 and TMPRSS2 in the lower airways of young people (421), it is not yet clear whether the level of ACE2 receptors can explain all the differences in the severity of COVID-19 (see ref). The results may however be confounded by the integrity of the primary lung defenses which determines access to most of the ACE2 receptors.

The importance of elevated glucose levels compromising the very first defense of the lung as the key barrier to contain the virus and prevent an avalanche of infection and complications is underappreciated in the CORD-19 database. Similarly, the potential importance of elevated glucose levels acting to provide the virus ideal conditions to coat the spike protein with glycans that can confer its pathogenicity and immune evasion is not clearly recognized, and it required piecing together numerous lines of evidence across multiple sources to reveal how elevated glucose could be involved in the complications of the disease, such as driving the immune response into a cytokine storm and participating in the dysregulation of coagulation and thrombotic features. Management of COVID-19 to some extent considers management of glucose as an important component, but if this hypothesis is correct, glucose management may need to become a core strategy.

Tight control and management of glycemia in COVID-19 patients may be critical in order to lower the first phase of infection and decrease the escalation of the disease. Managing glycemia in ICUs, where more than 80% of the patients were reported to present with hyperglycemia, also seems critically important. Even if glycemia is checked during hospital admission, only FPG of more than 7 mmol/L is considered serious, while even a moderate increase in FPG could be a risk factor. Furthermore, FPG reflects mostly the resting glucose levels and may not reveal sufficiently abnormal glucose metabolic capacity to clear glucose. Impaired glucose tolerance (IGT) should therefore be specifically tested, but the 2h-OGGT, the usual gold standard test for IGT involves ingestion of a large bolus of glucose could drive the progression of the disease. HbA1c measurements may serve as an alternative biomarker for IGT. Insulin levels should be systematically measured to detect undiagnosed diabetes, pre-diabetes or insulin resistance. In addition, the monitoring of glycemia during the course of the disease should at least be as important as the monitoring of the more common biomarkers such as IL-6, CRP, D-dimer or ferritin. Interventions to control glycemia should seriously be considered. Ang II and serum SP-D could be additional biomarkers to assess the risk of complications such as the cytokine storm and disseminated intravascular coagulation, and could also help in estimating the time course of the disease.

Even at a late stage of the pandemic, approaches to detect and manage abnormal glucose metabolism and administer appropriate glucose-lowering drugs or diets, are indicated to help weaken the infection. Metformin, an old, safe and FDA approved drug, is an interesting glucose management drug that also has multiple other effects that could be beneficial in the management of COVID-19. Metformin not only reduces blood glucose levels and clearance following a bolus of glucose, but also has anti-inflammatory properties as well as cardiovascular protective effects (i.e. anti-thrombotic). Diabetics on metformin seem to be at lower risk of severe disease, but studies on the potential beneficial effects in healthy and diverse groups and in groups presenting with the other comorbidities of COVID-19 are lacking.

The literature makes a strong case for using a ketogenic diet (KD) in the management of COVID-19, but there are challenges to any clinical implementation. It is contra-indicated for some groups such as those with type 1 DM (409, 410). The diet is difficult to set-up properly to ensure nutritional requirements. Beneficial effects of this diet, that has been found in the management of other diseases, are also usually expected over the long-term (389). The time needed for the body to enter ketosis also varies for different groups and the reasons are unclear. In addition, a transition to ketosis is often associated with flu-like symptoms (keto flu) (422), which may interfere with the innate immune response to the virus.

In summary, evidence that elevated blood glucose, arising from clinically or subclinical pathology in glucose metabolism or from induced hyperglycemia due to hospitalization, drug treatments and intravenous infusions in ICU should be considered as a biomarker that correlates with and hence is predictive of severity of COVID-19, as well as evidence that elevated glucose can cause an acceleration of virtually every step of the SARS-CoV2 infection, is overwhelming. Rigorous clinical studies are called for to determine whether elevated glucose is in fact the predominant driver of disease severity.

## METHODS

*All references are cited in the text or listed in Supplementary materials*.

### A) KNOWLEDGE GRAPH DESIGN AND IMPLEMENTATION

#### 1) Entities Extraction

##### Literature Database

The CORD-19 database (COVID-19 Open Research Dataset) is a freely available dataset of full text articles on COVID-19, SARS-CoV-2, and related coronaviruses, launched in March 2020 by the White House (https://www.kaggle.com/allen-institute-for-ai/CORD-19-research-challenge) and regularly updated (Wang et al, 2020). For our analysis, we used the CORD-19v47 including over 240,000 articles, with over 100,000 being full-text.

##### Named Entity Recognition (NER)

Our text mining pipeline for extracting information from the CORD-19 database starts by using machine learning models for named entity recognition (NER). These models are based on scispaCy models [Neumann, 2019] that we fine-tuned on a manually annotated subset of sentences from the CORD-19 dataset in order to recognize nine custom entity types of interest: “cell compartment”, “cell type”, “chemical”, “symptom / disease”, “drug”, “organ / system”, “organism”, “biological process / pathway”, and “protein”.

##### Entity linking

Extracted words in papers do not necessarily correspond to unique entities. In standard text search, this leads to ambiguity because an entity may be spelled differently in different papers or even within the same paper. Entity linking addresses this problem by resolving extracted entities to unique identifiers taken from a knowledge base while taking into account lexical variations as well as synonyms, aliases and acronyms. The resolved identifier can therefore be used to unambiguously reference an entity on subsequent text mining and knowledge graph tasks.

In this study, the National Cancer Institute Thesaurus (NCIt) ontology [Fragoso G, 2004] was used as the knowledge base to which extracted entities were linked. Such linking also gave us access to the semantics of the entities with their human-readable definitions and hierarchically structured semantic types (e.g, the entity *angiotensin-1* is a *AGT gene product*, which is a subtype of *peptide hormone’*, which is a *protein*). The entity types obtained during the extraction phase were therefore enriched with ontology types which allowed labeling of the resulting concepts into nine unique entity types summarized in the table below.

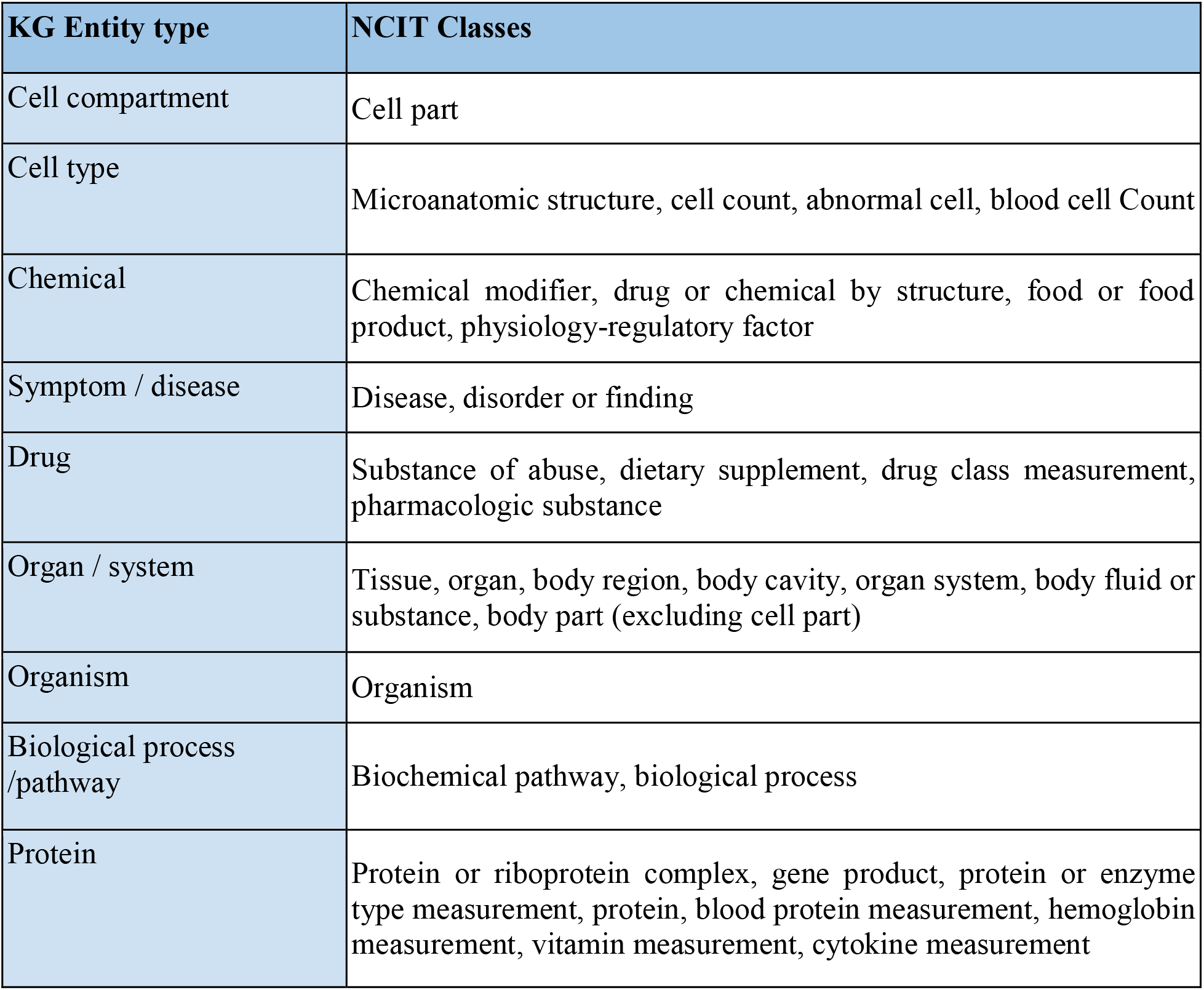

#### 2) Query-based literature search

Amongst the hundreds of thousands of articles contained in the CORD-19 database, we wanted to be able to focus our analysis on subsets of publications. An information retrieval tool for literature search was therefore implemented, allowing users to query the CORD-19 database both for various simple filtering criteria (e.g. “publication date” or “journal”) and for relevance with respect to a given query. We used a machine learning model based on BioBERT [Lee, 2020] and fine-tuned it on the CORD-19 dataset in order to produce sentence embeddings vectors. For any given sentence, its embedding vector encodes its semantic information, so that semantically similar sentences are mathematically represented by similar vectors. Then, we use the cosine distance to compute similarity between pairs of sentences embedding vectors, which enables us to rank the content of the CORD-19 with respect to the query and retrieve the most relevant articles. The criteria set for the search in the current study are the following:

- Query: “Glucose as a risk factor for COVID-19”
- Granularity: “Articles”
- Number of top articles: “3,000”
- Date range: “2000-2020”
- Journal Type: “All

#### 3) Knowledge graph construction method

We examined co-mentions of entity pairs on a paper- and paragraph-level, adding an edge between a pair of entities if they co-occur in at least one paper. We then assigned several weight metrics to each co-mention edge including raw co-mention frequency, positive pointwise mutual information (PPMI) and normalized pointwise mutual information (NPMI) [Bouma, G. (2009)] calculated based on the co-occurrence in the same papers and the same paragraphs. In this context, the presence of an edge between a pair of entities can be interpreted as the presence of some association between the pair of concepts they represent, and the corresponding edge weights quantify the strength of such association. To assign weights on the nodes of the constructed graph that would reflect the importance of entities in the extracted dataset, we have computed nodes’ *weighted degree centrality* using the previously described edge weights (given by raw frequency, PPMI and NMPI).

##### - CORD-19 knowledge graphs

Following the methods described above, we constructed two knowledge graphs. The first knowledge graph is based on the entire CORD-19 dataset. Out of more than 400,000 entities extracted and linked during data preparation, the 10,000 most frequently mentioned entities were selected and used as nodes of the knowledge graph. The constructed graph is very dense containing over 44 million edges (resulting in a density of 0.87, i.e. 87% of all possible pairs of co-mentions) out of which 12 million edges have non-zero paragraph-level co-occurrence (making the density of the paragraph-based co-mention network 0.25).

The second knowledge graph was built using the results of the query-based literature search (see section 2). Out of more than 20,000 extracted entities, the 1,500 most frequently mentioned entities were selected and used as nodes of the knowledge graph. Only the edges that correspond to the non-zero paragraph-level co-occurrence were considered. As a result, we generated approximately 700,000 edges, giving a total density of 0.62.

#### 4) Community detection

To partition the knowledge graph constructed based on the entire CORD-19 dataset into clusters of strongly associated entities, we performed node community detection using the Louvain algorithm [Danon, 2005] on paragraph-level co-occurrences. A community represents a cluster of nodes whose connections are stronger within the community than with the rest of the network. The algorithm detected five different communities of entities. Having examined the most frequent entities in each community, we have mapped the communities to the following five topics: biology of viruses, diseases and symptoms, immune response, infectious disorders, chemical compounds (Supplementary Figure 3). Note that despite the fact that our network is given by a highly dense graph, such community detection is still possible when taking into account edge weights (in our case, the NPMI values of the co-occurrence edges). The resulting partition gives us a modularity value of 0.21.

#### 5) Minimum spanning tree analysis

To sparsify our knowledge graph and gain insight into the most important and relevant associations between entities, we computed the minimum weight spanning tree with the weight being assigned to the inverse of the edge NPMI based on paragraph-level co-occurrence (therefore, a higher pointwise mutual information between entities implies a smaller distance). The minimum spanning trees on Figure 4 and Supplementary Figure 4 were generated from the knowledge graph built using the results of the query-based literature search.

#### 6) Best mutual information pathways (BMPIs) search

To gain insight on the most important and informative sets of entities relating ‘glucose’ and ‘COVID-19’ or ‘SARS-COV-2’ in the literature, we have used the approach called “best mutual information pathways” (BMIPs). Such pathways are constructed using the shortest weighted paths from the source to the target entity (e.g. from ‘glucose’ to ‘COVID-19’ or ‘SARS-COV-2’), where the weight corresponds to the paper-level NPMI (i.e. the higher NPMI associated with an edge, the smaller the ‘distance’ between the corresponding source and target entities).

To be able to navigate and explore the literature, we aimed to identify concepts that relate a source to a target entity. Therefore, we focused on finding a set of such shortest paths, rather than a single shortest path. Classical algorithms exist to find the N shortest paths between two nodes in a weighted graph (such as [Yen, Jin Y (1971)]). However, due to the density of our graph, such algorithms perform poorly in terms of execution time. For the same reason it is extremely rare that one of the shortest paths between two nodes consists of more than two hops. We therefore adopted a naïve strategy that exploits this property. We first find all the indirect shortest paths (discarding the direct edge from the source to the target) in terms of number of hops ignoring the edge weight (usually such paths consist of two hops, which greatly reduces the space of all possible paths). Then, the algorithm ranks these paths by computing the cumulative distance score and chooses the ones with the largest score. Such distance scores are simply given by the sum of the inverse of the NPMI associated with the path’s edges. Finally, to further explore the space of co-mentioned entities in depth, we can run the path search procedure in a *nested* manner. For each edge encountered on a path *e_1_, e_2_, …, e_n_* from *e_1_* to *e_n_,* we further *expanded* it into n shortest paths between each pair of successive entities (i.e. paths between *e_1_* and *e_2_, e_2_* and *e_3_*, etc.). For example, the graph in Figure 3 is obtained by aggregating the nodes and the edges encountered by, first, searching for the 20 BMIPs and, second, by further expanding each of the encountered edges into their five BMIPs.

#### 7) Knowledge graphs visualization

We have developed graphical interfaces that allow performing interactive entity curation, exploration and analysis of co-occurrence graphs, that were used to produce graphs and BMPIs in the figures. Semantic issues that were not identified by the ontology linking process were fixed during this manual curation process. Then, all knowledge graphs-related figures were produced using the Gephi software (https://gephi.org/).

The sizes of the nodes are proportional to their weighted degree centralities. The color of the nodes corresponds to different entity types (except for Supplementary Figure 3 where colors correspond to different communities). The thickness values of the edges correspond to NPMI values, while their length is arbitrary.

#### 8) Source code

The source code for the pipeline stages relative to Semantic Literature Search and Named Entity Recognition can be found here: https://github.com/BlueBrain/Search/tree/v0.1.0

The ML models, and the data used to train them, can be found here: https://zenodo.org/record/4589007

The code, data and instructions to reproduce the entity linking, knowledge graphs generation and analysis can be accessed from: https://github.com/BlueBrain/BlueBrainGraph/tree/master/cord19kg.

### B) GLUCOSE-DEPENDANT SARS-CoV-2 INFECTION COMPUTATIONAL MODELS

The computational model of the glucose dependents of SARS-CoV-2 infectivity in alveolar epithelial cells was written and implemented in the MATLAB simulation language (https://www.mathworks.com/).

The parameters and governing equations are provided below.

#### 1) Parameters

| Name | Value | Source |
| --- | --- | --- |
| Glucose, blood, normoglycemia (gluc_b) | 5 mM | - |
| Glucose, blood, hyperglycemia (gluc_b_hyp) | 10 mM | - |
| Glucose, epithelium, initial (gluc_epi_init) | 2 mM | Bearham et al, 2019 |
| Glucose, ASL, initial normal (gluc_asl_init) | 0.4 mM | Garnett et al, 2012<br>Pezzulo et al, 2011 |
| Glucose, ASL, initial hyper (gluc_asl_h_init) | 1.2 mM | Garnett et al, 2012<br>Baker et al, 2007 |
| Lactate, epithelial, initial (lac_epi_init) | 1 mM | Fisher and Dodia, 1984 |
| Lactate, ASL, initial (lac_asl_init) | 1 mM | Fisher and Dodia, 1984 |
| Paracellular resistance, normal (Rt_n) | 453 $\Omega \cdot \text{cm}^2$ | Garnett et al, 2012b |
| Paracellular resistance, hyper (Rt_h) | 225 $\Omega \cdot \text{cm}^2$ | Garnett et al, 2012b |
| pH, ASL, initial (pH_asl_init) | 7.0 | Amirkhanian and Merritt, 1995 |
| pH, epithelium, initial (pH_epi_init) | 7.2 | Boron, 2004 |
| $K_m$, SARS-CoV-2 - ACE2 receptor (KmACE2) | 1.6 x 10<sup>-5</sup> mM | Bar-On et al, 2020; Wrapp et al, 2020 |
| $K_m$, Na-coupled glucose transporter (KmSgt1) | 0.3 mM | Baker and Baines, 2018 |
| $K_m$, lectin - glucose (KmLectg) | 1.8 mM | Schwartz et al, 1993 |
| $K_m$, lectin – SARS (KmLectv) | 0.45 mM | Kussrow et al, 2009 |
| $K_m$, MCT4 apical lactate transporter (KmMCTapi) | 1.7 mM | Contreras-Baeza et al, 2019 |
| Lactate dehydrogenase, forward (kLDHeplus) | 0.071 | Jolivet et al, 2015 |
| Lactate dehydrogenase, reverse (kLDHeminus) | 1.59 | Jolivet et al, 2015 |
| Glucose, diffusion constant (difuseK) | $5.65 \times 10^3 \text{ m}^2/\text{sec}$ | |
| Lectin, ASL (lectin) | 5.8 mM | Gaunsbaeket al, 2013 |

#### 2) Definition of viral loads

The total range between low and high viral loads was 10x according to clinical findings [Anand and Mayya, 2020]. The intermediate load was chosen to be closer to the higher viral load in order to simulate a non-specific saturating effect, as if close to the top of a dose-response binding curve; then the intermediate load is 5x the low load, whereas the high load is 2x the intermediate load. This saturating effect would emerge from multiple interacting factors for which there is no clear data, but mostly from the principle of ACE2 receptor availability, which is assumed to be finite in this simulation and does not grow with larger viral loads. As control, simulations were also run with the intermediate viral load being equidistant between low and high (data not shown), that did not affect the qualitative outcome of the models

#### 3) Other Abbreviations

|  |  |
| --- | --- |
| asl | Airway surface liquid |
| sc2 | SARS-CoV-2 time-course in ASL (model “sneeze” stimulus) |
| sc2_asl_effect_n | Effective available sc2 in normoglycemic ASL |
| sc2_asl_effect_h | Effective available sc2 in glycemc ASL |
| sc2_asl | viral load in ASL 1 |
| sc2_aslb | viral load in ASL 2 |
| sc2_aslc | viral load in ASL 3 |
| ACE2V_n | sc2-ACE2 binding during low load stimulus, normoglycemic |
| ACE2V_h | sc2-ACE2 binding during low load stimulus, hyperglycemic |
| ACE2V_nb | sc2-ACE2 binding during intermediate load stimulus, normoglycemic |
| ACE2V_hc | sc2-ACE2 binding during intermediate load stimulus, hyperglycemic |
| ACE2V_nb | sc2-ACE2 binding during high load stimulus, normoglycemic |
| ACE2V_hc | sc2-ACE2 binding during high load stimulus, hyperglycemic |
| ACE2V_r | Glucose-dependent sc2 endocytosis, low viral load |
| ACE2V_rb | Glucose-dependent sc2 endocytosis, intermediate viral load |
| ACE2V_rc | Glucose-dependent sc2 endocytosis, high viral load |
| MCTapifrac | Activity of apical endothelial lactate transporter |
| MCTbasfrac | Activity of basal endothelial lactate transporter |
| lac<sub>epi</sub> | Epithelial lactate concentration |
| gluc<sub>aslr</sub> | ASL glucose concentration range |
| gluc<sub>brange</sub> | Blood glucose concentration range |
| Jgluc<sub>nr</sub> | Paracellular glucose flux, normoglycemic |
| Jgluc<sub>hr</sub> | Paracellular glucose flux, hyperglycemic |
| Rt<sub>x</sub> ; x =n, h or range | Paracellular resistance, n=normoglycemia, h= hyperglycemia |
| junction<sub>conduct</sub> | Paracellular conductance (= 1/Rt) |
| Sglt1 | Sodium-dependent glucose transporter |
| vireprate<sub>epi</sub> | virus replication rate in epithelial cell, low load |
| vireprate<sub>epib</sub> | virus replication rate in epithelial cell, intermediate load |
| vireprate<sub>epic</sub> | virus replication rate in epithelial cell, high load |
| vinumber<sub>epi</sub> | Number of virion units in epithelial cell, low load |
| vinumber<sub>epib</sub> | Number of virion units in epithelial cell, intermediate load |
| vinumber<sub>epic</sub> | Number of virion units in epithelial cell, high load |

#### 4) Governing Equations

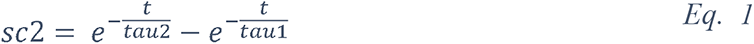

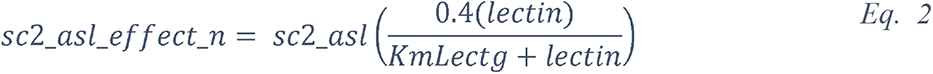

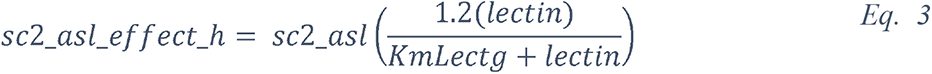

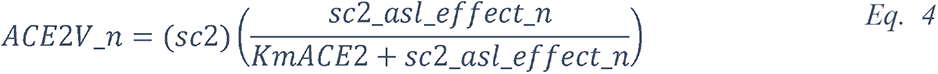

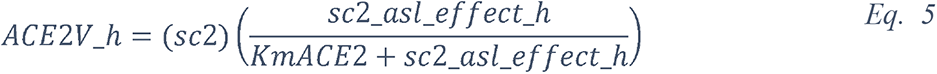

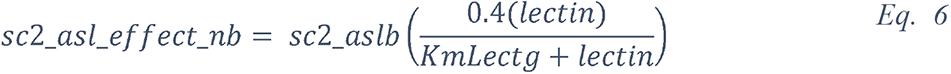

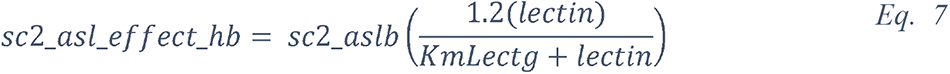

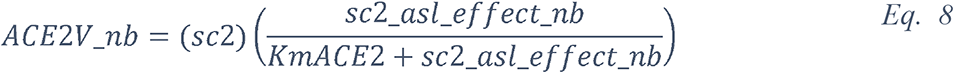

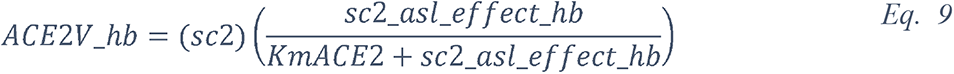

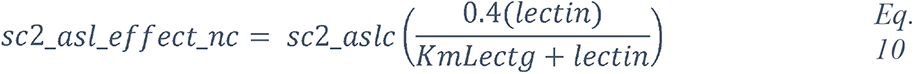

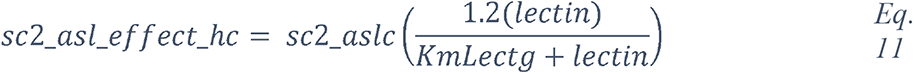

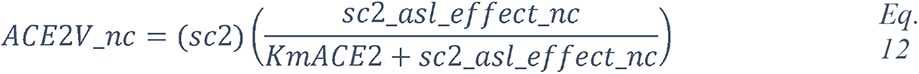

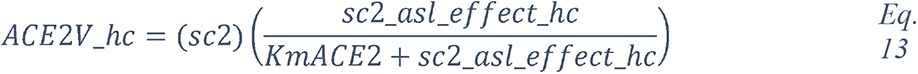

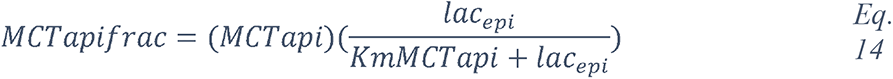

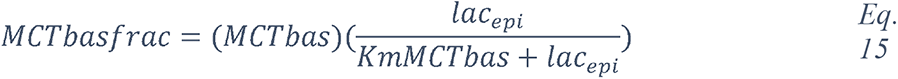

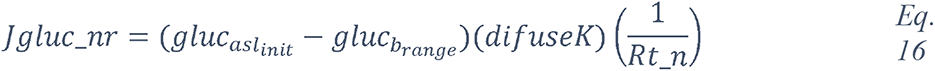

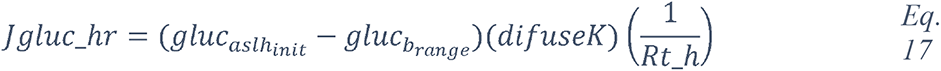

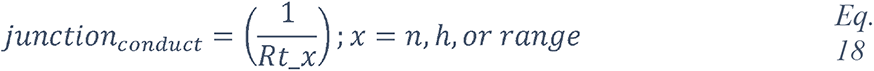

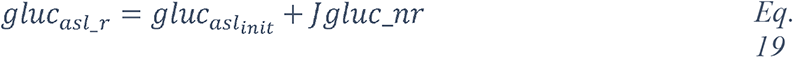

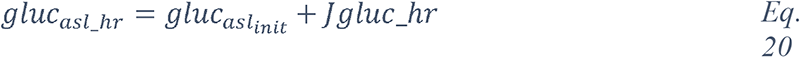

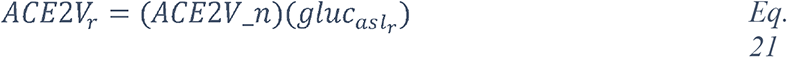

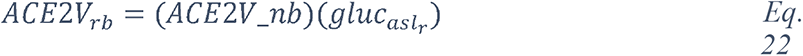

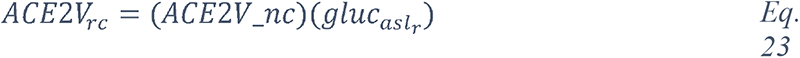

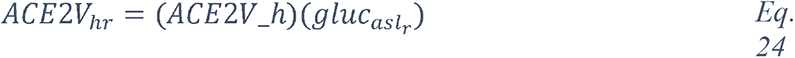

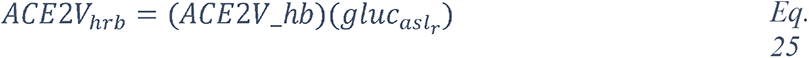

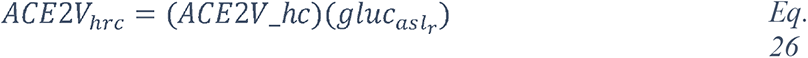

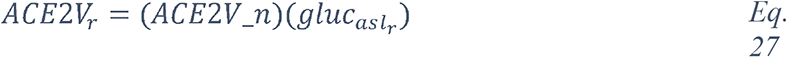

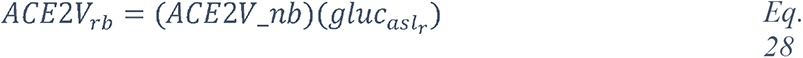

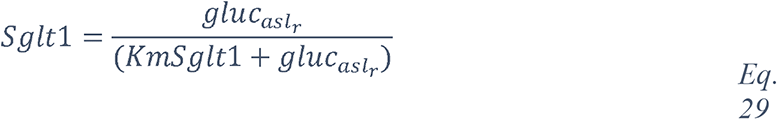

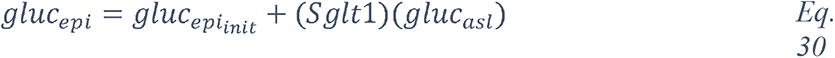

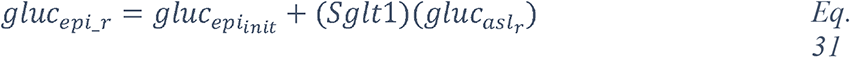

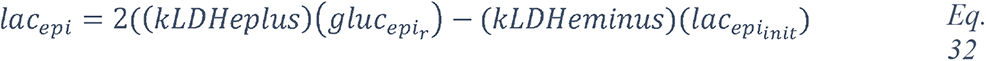

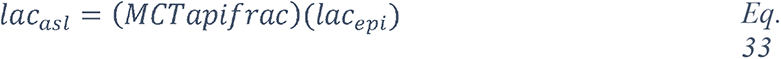

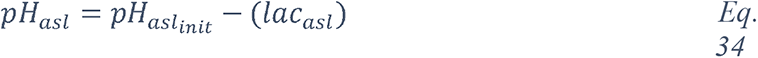

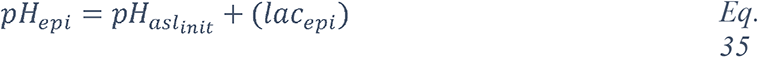

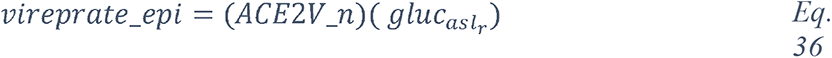

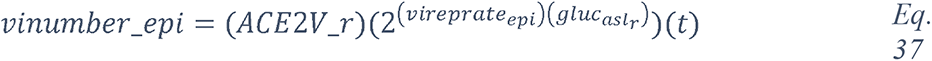

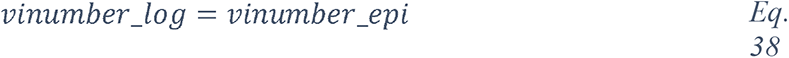

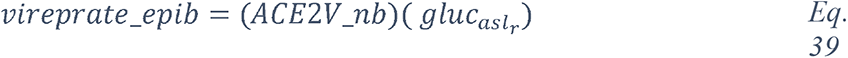

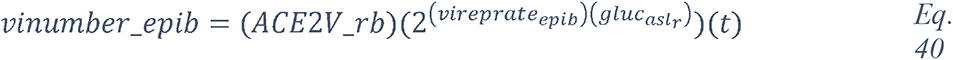

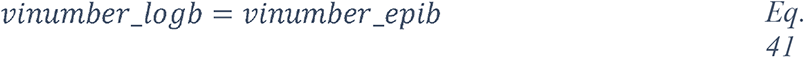

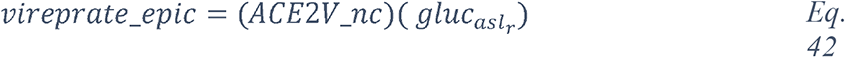

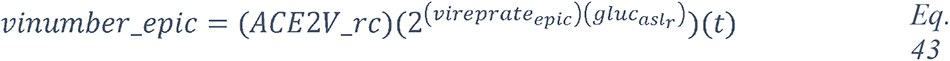

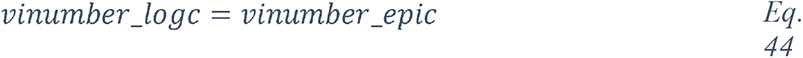

### C) BIOEXPLORER DESIGN AND IMPLEMENTATION

The Blue Brain BioExplorer (BBBE) application is built on top of Brayns (https://github.com/BlueBrain/Brayns), the BBP rendering platform. The role of the application is to use the underlying technical capabilities of the rendering platform to create large scale and accurate 3D scenes from Jupyter notebooks. The code for the model will be open sourced upon publication.

#### 1) Source code

The complete documentation is available here : https://bluebrain.github.io/BioExplorer/

The source code is available here: here: https://github.com/BlueBrain/BioExplorer

#### 2) Components ID and dimension

##### Macrophage and membranes

The 3D model of the macrophage represented in Figure 16 was obtained from https://www.turbosquid.com/3d-models/3d-lymphocytes-neutrophil-basophil-1168937

The viral and host cell membranes were generated from phospholipids structures created following the process described in the VMD (https://www.ks.uiuc.edu/Research/vmd/) Membrane Proteins Tutorial (http://www.ks.uiuc.edu/Training/Tutorials/). Then, an assembly of phospholipids elements is generated by the BioExplorer, with a given shape, and a given number of instances of phospholipids.

#### Molecular components

Dimension and PDB-ID of each molecular component represented in Figures 11, 12, 16, 19 and in the Movie are described in the panel below:

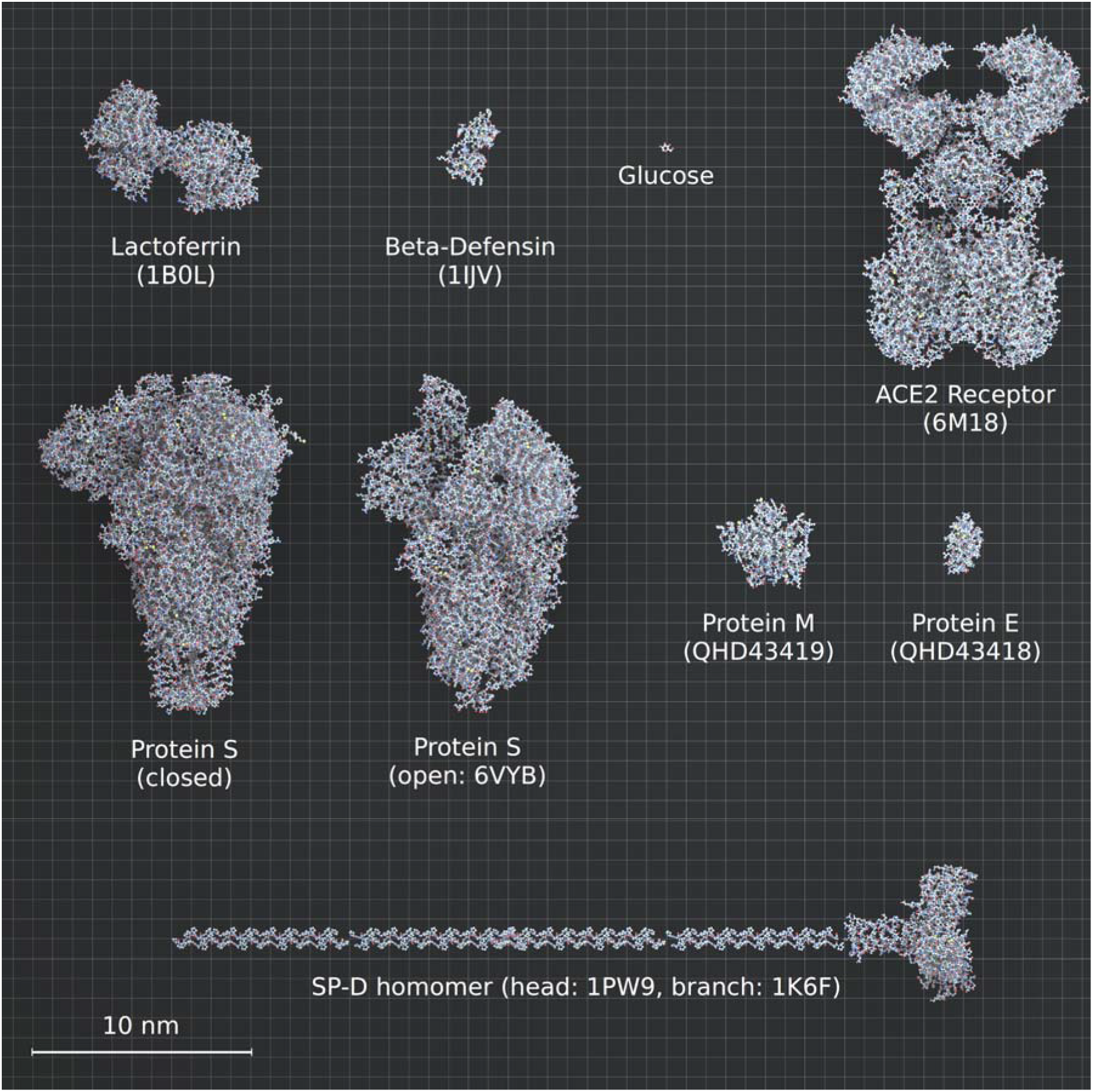

The spike in closed conformation was self-generated using “I-Tasser Protein structure prediction” and “Modeller” from Uniprot ID P0DTC2. The model for ACE2 corresponds to ACE2/B°AT1 complex (Yan et al, 2020). The same models were used to generate surfactant protein-D and surfactant protein-A, only the conformation was different. Dimension and conformation of SP-D and SP-A were obtained from literature (Watson et al, 2018; Hsieh et al, 2018).

#### 3) Component numbers and concentrations (Related to Figures 16, 19 and Movie)

Each viral particle is 90 nm diameter with 62 spikes, 42 “E proteins” and 50 “M proteins” per particle (Neuman et al, 2006; Bar-On et al, 2020).

The images represent a cube with 800 nm side, *i.e.* a volume of 0.512 um^3^.

The number of each component in the 0.512 um^3^ volume, in the two different conditions (0.4 or 1.2 mM glucose), is indicated in the following table:

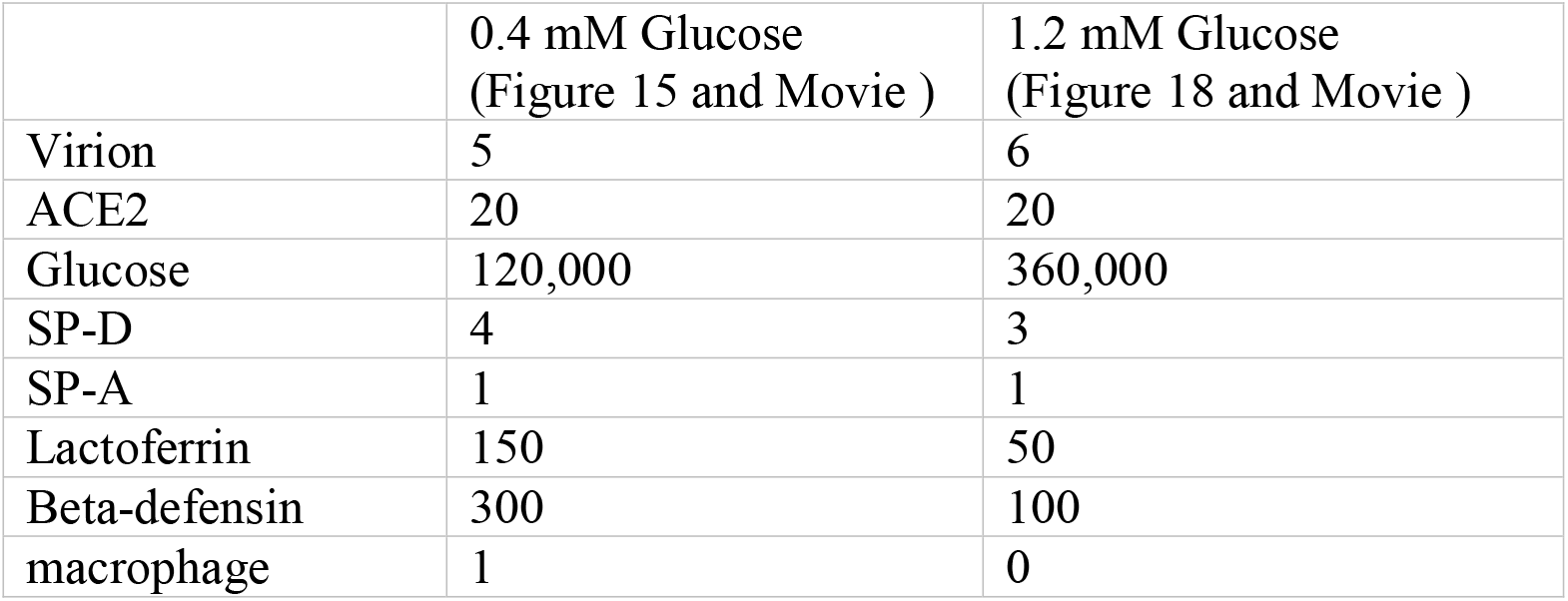

#### 4) Glycans

##### a. Formula

Glycan trees are retrieved from Glycam Builder (http://glycam.org/Pre-builtLibraries.jsp) with the corresponding formula for each of the four types of N-glycans (No diversity was included in the present model).

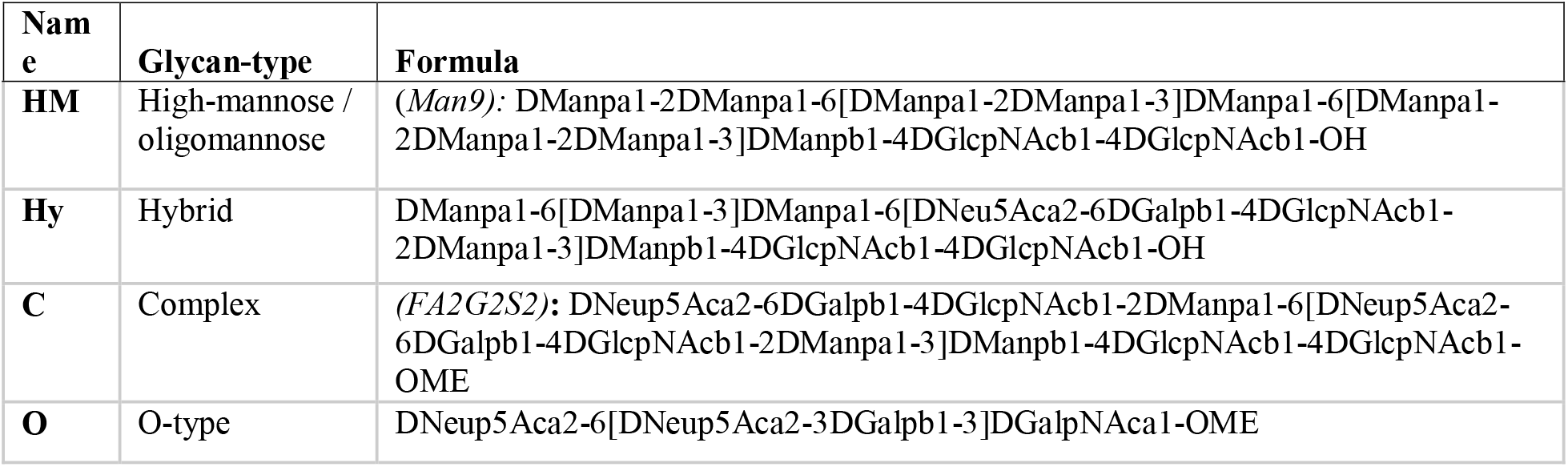

##### b. Types and positions

The glycan-type (HM, complex or hybrid) with the highest representation reported for each specific site was considered, hence no microheterogeneity is included in our representation.

- Viral Spike* (From Watanabe *et al,* 2020 and Uniprot ID P0DTC2):

| N-site (aa position) | Tree Type |
| --- | --- |
| 17 | C |
| 61 | HM |
| 74 | C |
| 122 | HM |
| 149 | C |
| 165 | C |
| 234 | HM |
| 282 | C |
| 331 | C |
| 343 | C |
| 603 | HM |
| 616 | C |
| 657 | C |
| 709 | HM |
| 717 | HM |
| 801 | HM |
| 1074 | HM |
| 1098 | C |
| 1134 | C |
| 1158 | C |
| 1173 | C |
| 1194 | C |
| T-site (aa position) |  |
| 323 | O |
| 325 | O |

\*The above-mentioned glycans are all present on the spike in closed conformation. However, glycans at positions 17, 74, 149, 165, 1158, 1173 and 1194 are missing in the representation of the spike in its open conformation, that is inherent to the model provided by PDB (ID:6VYB).

- Human ACE2 (From Shajahan *et al,* 2020 and Uniprot ID Q9BYF1):

| N-site (aa position) | Tree Type |
| --- | --- |
| 53 | C |
| 90 | C |
| 103 | C |
| 322 | C |
| 432 | C |
| 546 | Hy |
| 690 | C |
| T/S-site (aa position) |  |
| 155 | O |
| 730 | O |

- Viral M protein (From Fung *et al,* 2018 and Uniprot ID P0DTC5):

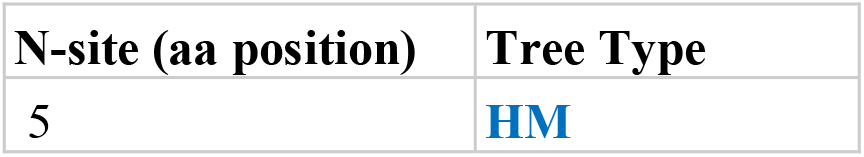

| N-site (aa position) | Tree Type |
| --- | --- |
| 5 | HM |

- Viral E protein (From Fung *et al,* 2018 and Uniprot ID P0DTC4):

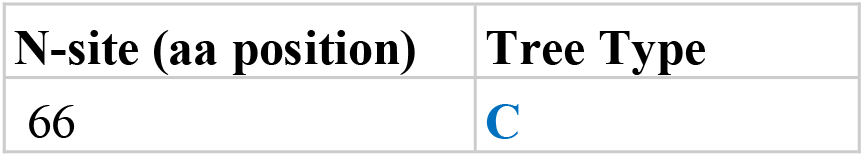

## Supporting information

Logette et al, supp material

## Data Availability

The links to access the simulation codes used to generate the datasets presented in this study are provided in the related section of methods, and can be run using the instructions in the "README.md" files. All simulation codes will be publicly open sourced upon publication.

https://github.com/BlueBrain/BlueBrainSearch

https://github.com/BlueBrain/BlueBrainGraph

https://github.com/BlueBrain/BioExplorer

## Abbreviations

ACE2: angiotensin-converting enzyme 2
AGEs: advanced glycation end products
Ang II: angiotensin II
ARDS: acute respiratory distress syndrome
ASL: airway surface liquid
BMI: body mass index
CFR: case fatality rate
CRD: carbohydrate recognition domain
FPG: fasting plasma glucose
GLUT: glucose transporter
ICU: intensive care unit
IFG: impaired fasting glucose
IGT: impaired glucose tolerance
KD: ketogenic diet
OGTT: oral glucose tolerance test
PPG: postprandial glucose
ROS: reactive oxygen species

## Authors contributions

Investigations, E.L., C.L. and H.M.; Data collection and literature review, E.L., C.L. and M.B.; Blue Brain BioExplorer design and implementation, C.F. and E.L.; Computational infection models, J.S.C. and D.K.; Design and implementation of Machine Learning models, F.C., S.S., E.D., J.K. and P.A.F. Design and implementation of a Knowledge Graph building process, F.S., E.O., A.K.K., P.A.F., S.K. and E.S.; Figures design, C.M.; Writing, E.L., C.L. and H.M.; Supervision, E.L. and H.M. All authors contributed to the article and approved the submitted version.

## Acknowledgments

We thank Polina Shichkova for technical support on protein modeling for Blue Brain BioExplorer, the BBP infrastructure team for providing help to host web server and storage and Karin Holm for editing of the manuscript and support for the submission process.

## Data Availability Statement

The links to access the simulation codes used to generate the datasets presented in this study are provided in the related section of methods, and can be run using the instructions in the “README.md” files.

