## Supplementary material for "Elevated Blood Glucose Levels as a Primary Risk Factor for the Severity of COVID-19": Logette et al, supp material

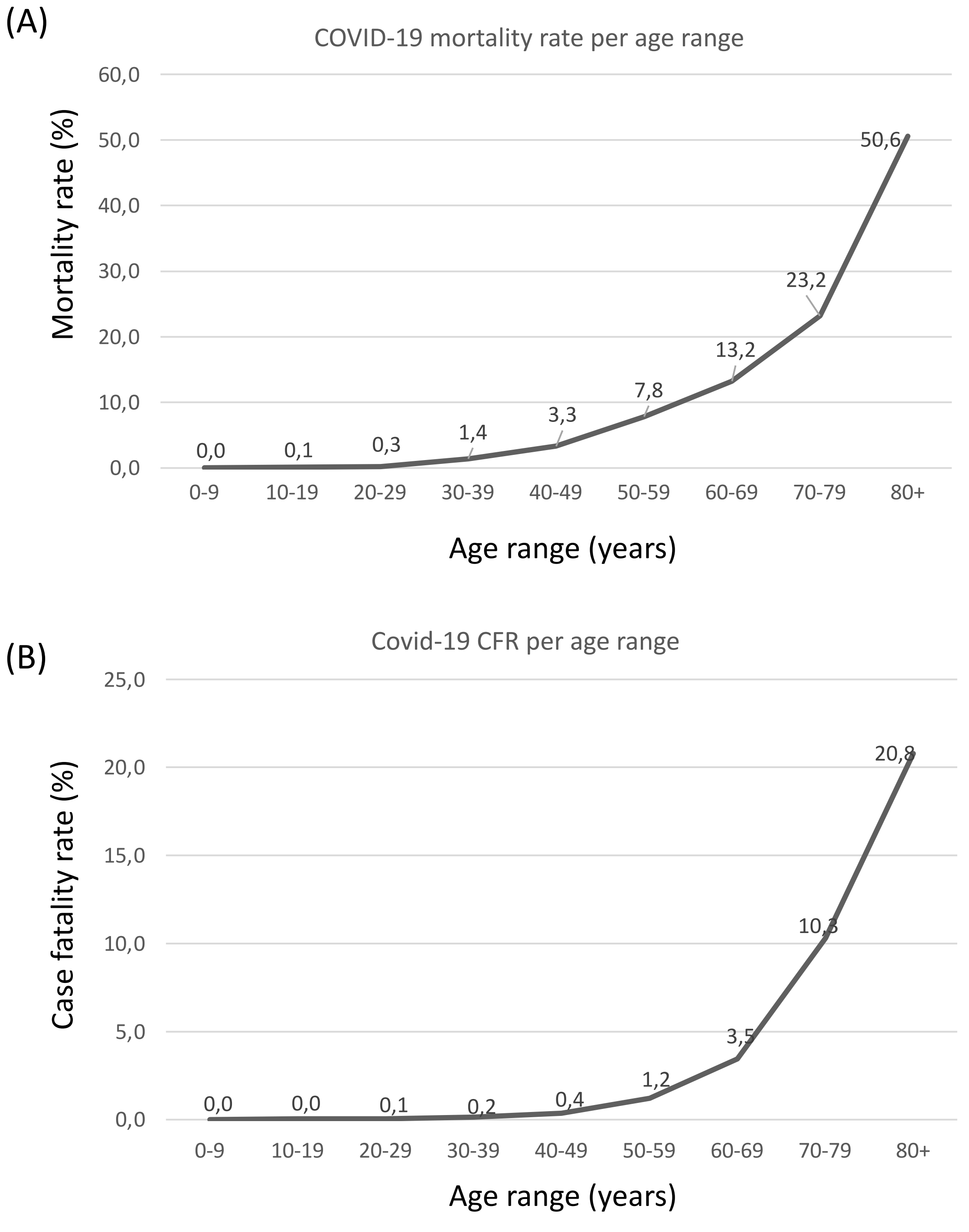


Supplementary Figure 1: (A) Average COVID-19 mortality rate per age range (i.e the percentage of COVID-19 deaths per age range among the total number of COVID-19 deaths). (B) Average COVID-19 case fatality rate (CFR) per age range (i.e the percentage of COVID-19 deaths among the COVID-19 positive cases per age range). Data source are reported in Supplementary Tables 1A and B respectively.

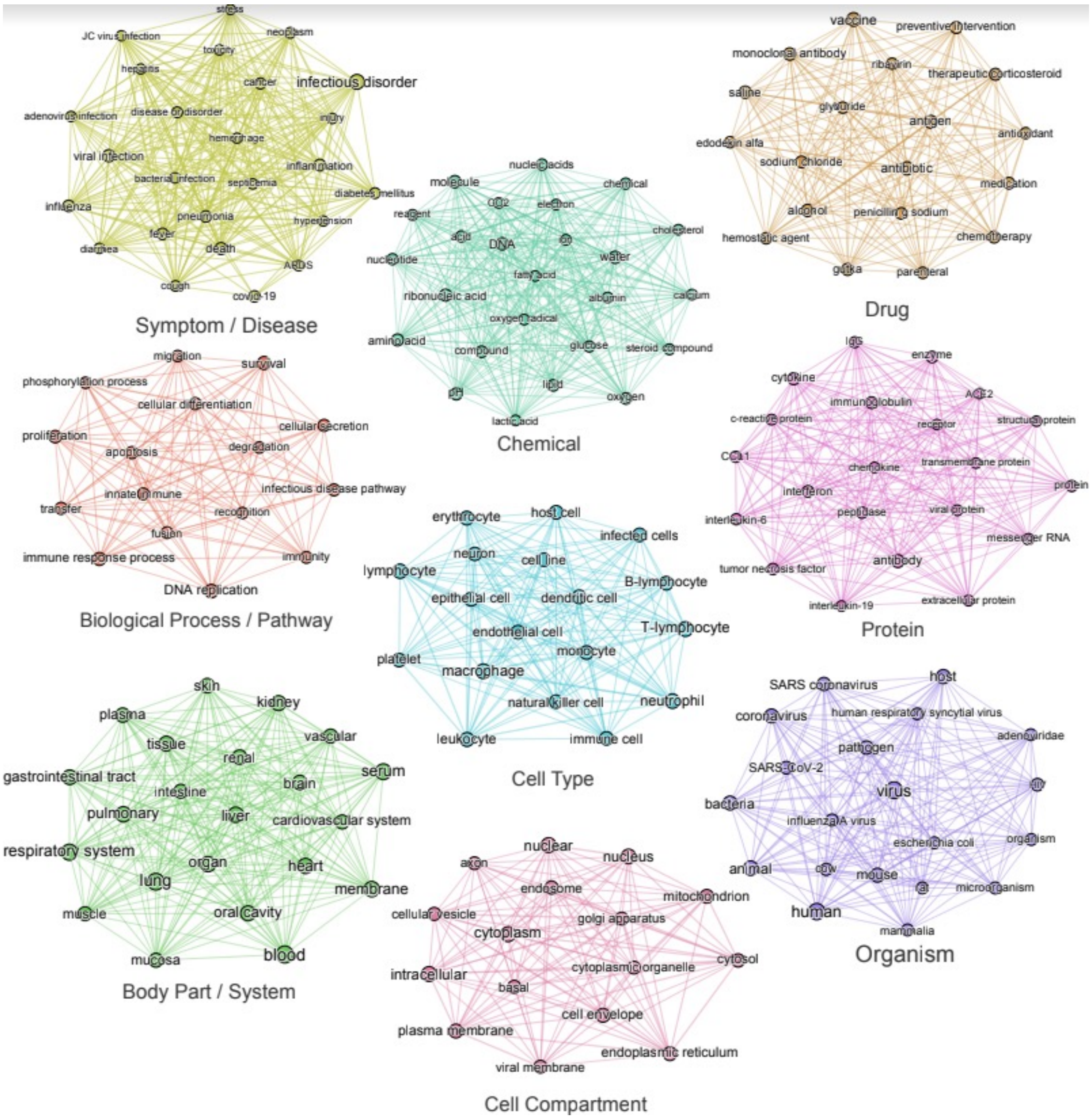


Supplementary Figure 2: Co-mention network overview with zoom on single-entity type subnetworks. Labeled subfigures represent samples of the most frequent entities for each respectively colored single-entity type subnetwork from Figure 2.

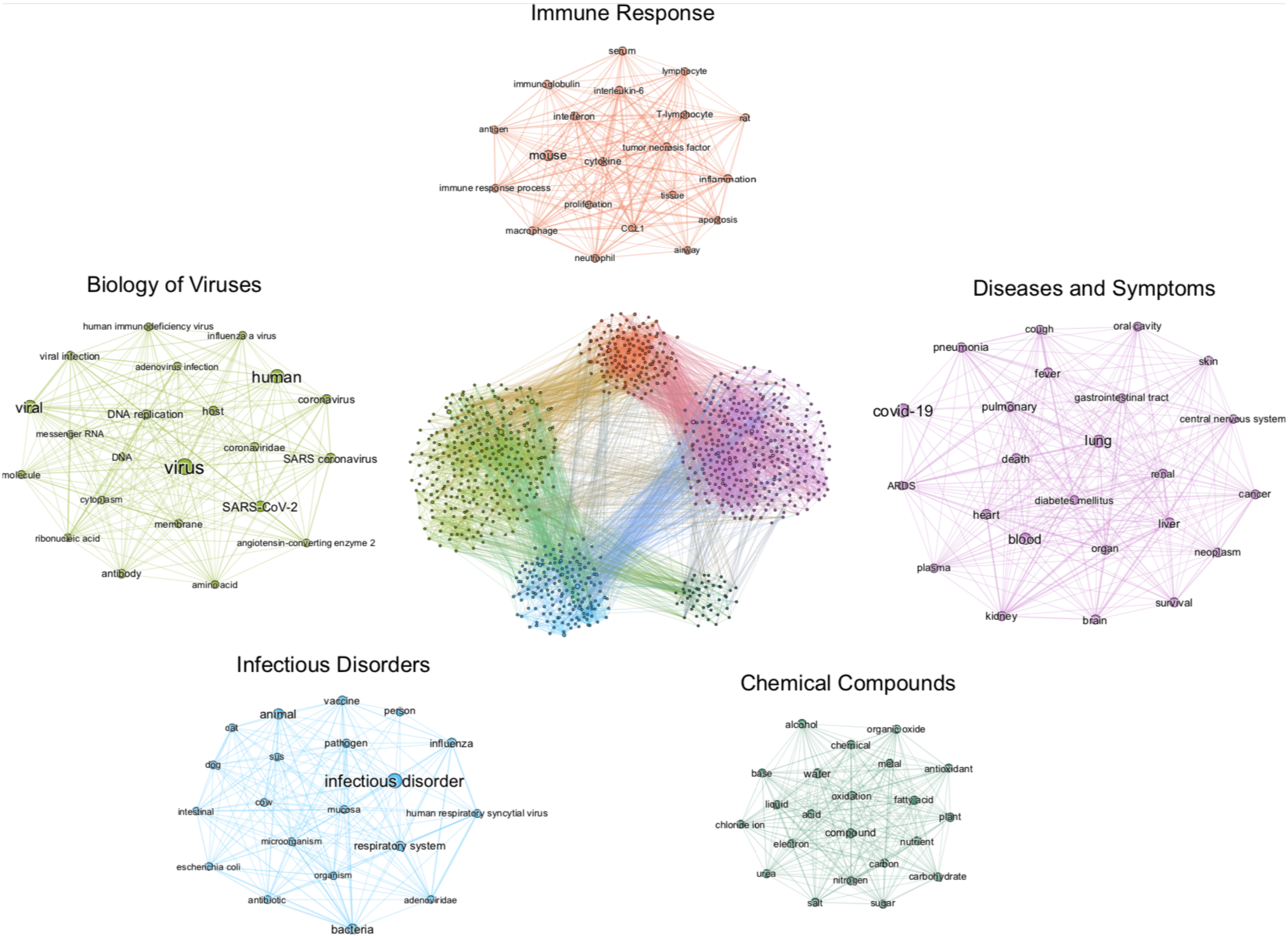


Supplementary Figure 3: Co-mention network communities overview. The figure in the center displays a sample of the knowledge graph containing ~1’000 nodes representing the most frequent entities and edges whose NPMI weight > 0.2. Node sizes are proportional to the weighted degree of nodes, node colors represent different communities detected using the Louvain algorithm and edge thickness is proportional to the NPMI weight of a given entity co-occurrence. Labeled subfigures represent samples of the most frequent entities for each respectively colored community.

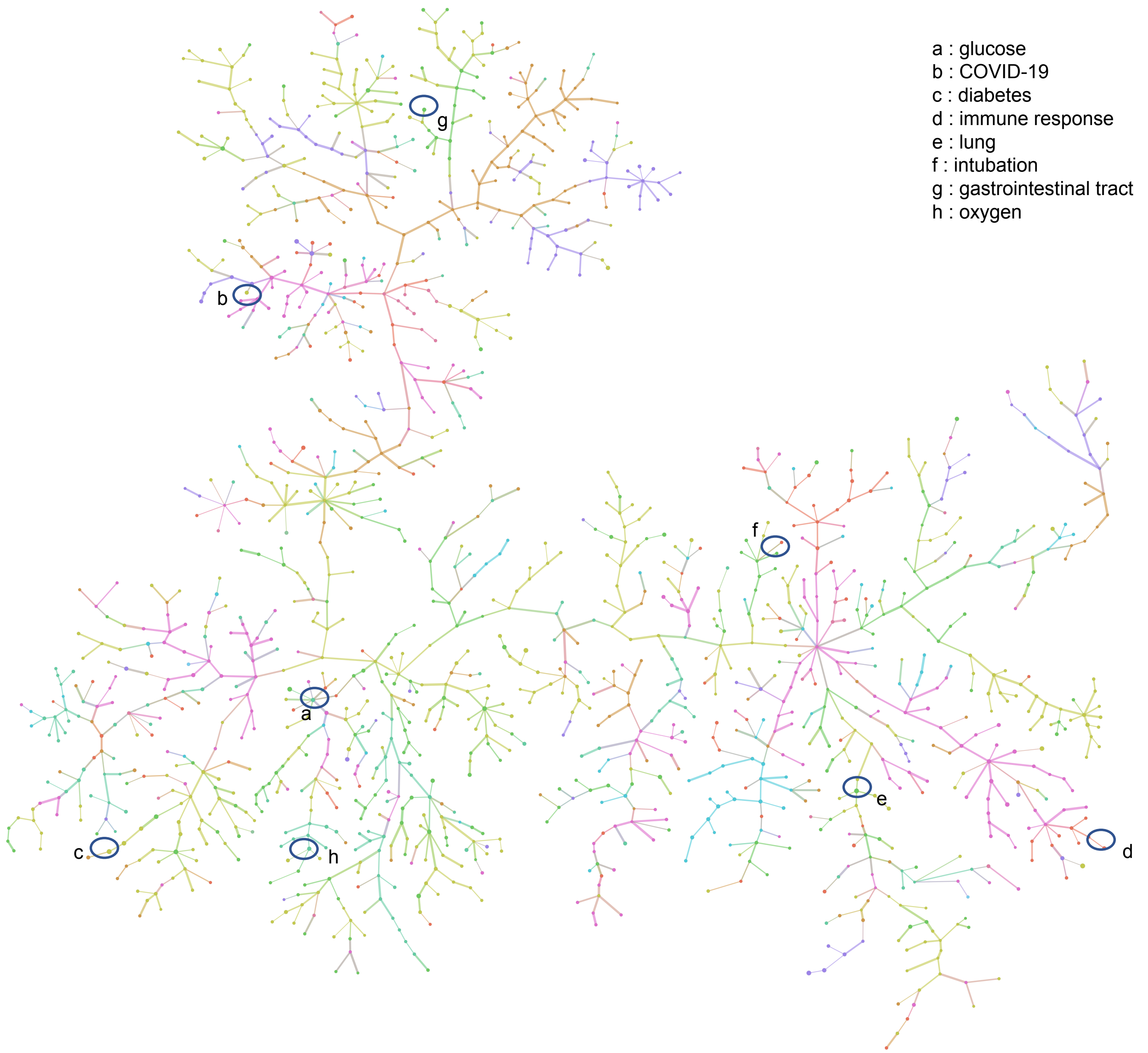


Supplementary Figure 4: Spanning-tree constructed from the 1,500-most frequent entities extracted from the 3,000-most relevant papers in the CORD-19v47 database, according to the query "Glucose is a risk factor for COVID-19". Positions of few key entities are indicated. A high resolution pdf version is available (Figure S4 high res).

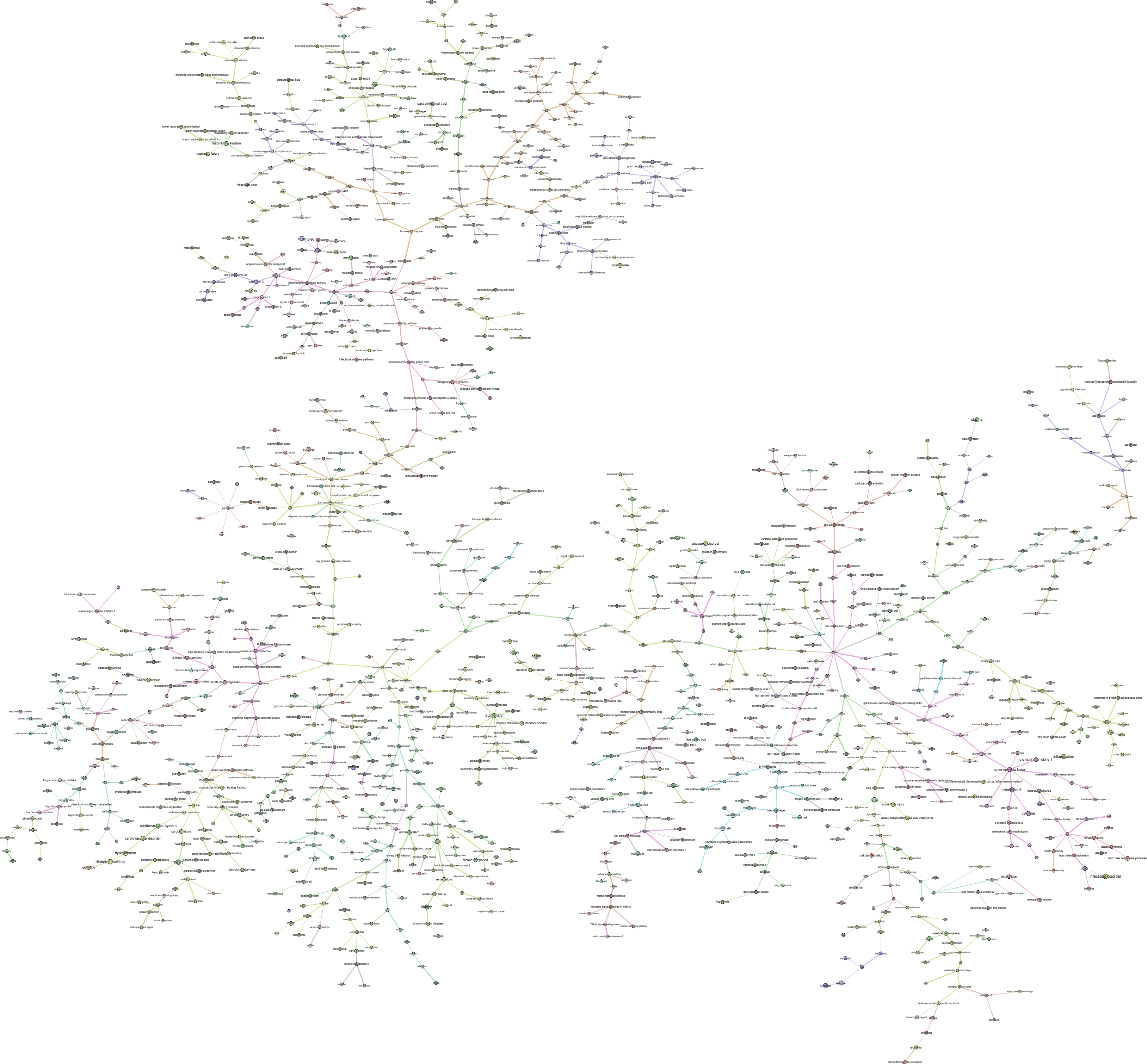

respiratory system
respiratory failure
influenza
cerebrospinal fluid
gastrointestinal tract
hemorrhage
hepatitis c virus
viral infection
membrane
coronavirus
sars coronavirus
pneumonia
staphylococcus aureus
streptococcus pneumoniae
pseudomonas aeruginosa
microorganism
escherichia coli
klebsiella pneumoniae
depression
infectious disease pathway
therapeutic corticosteroid
central nervous system
hemoglobin
erythrocyte
cardiovascular system
cardiovascular disorder
hypertension
diabetes mellitus
oxidative stress
mitochondrion
c-reactive protein
insulin resistance
pulmonary
chronic obstructive pulmonary disease
kidney
chronic kidney disease
acute renal failure
disease or disorder
apoptosis
proliferation
cellular differentiation
human immunodeficiency virus
tumor necrosis factor
interleukin-6
inflammation
infectious disorder
immune response process
macrophage
acute respiratory distress syndrome
cerebral hemisphere
human
animal
chemotherapeutic perfusion

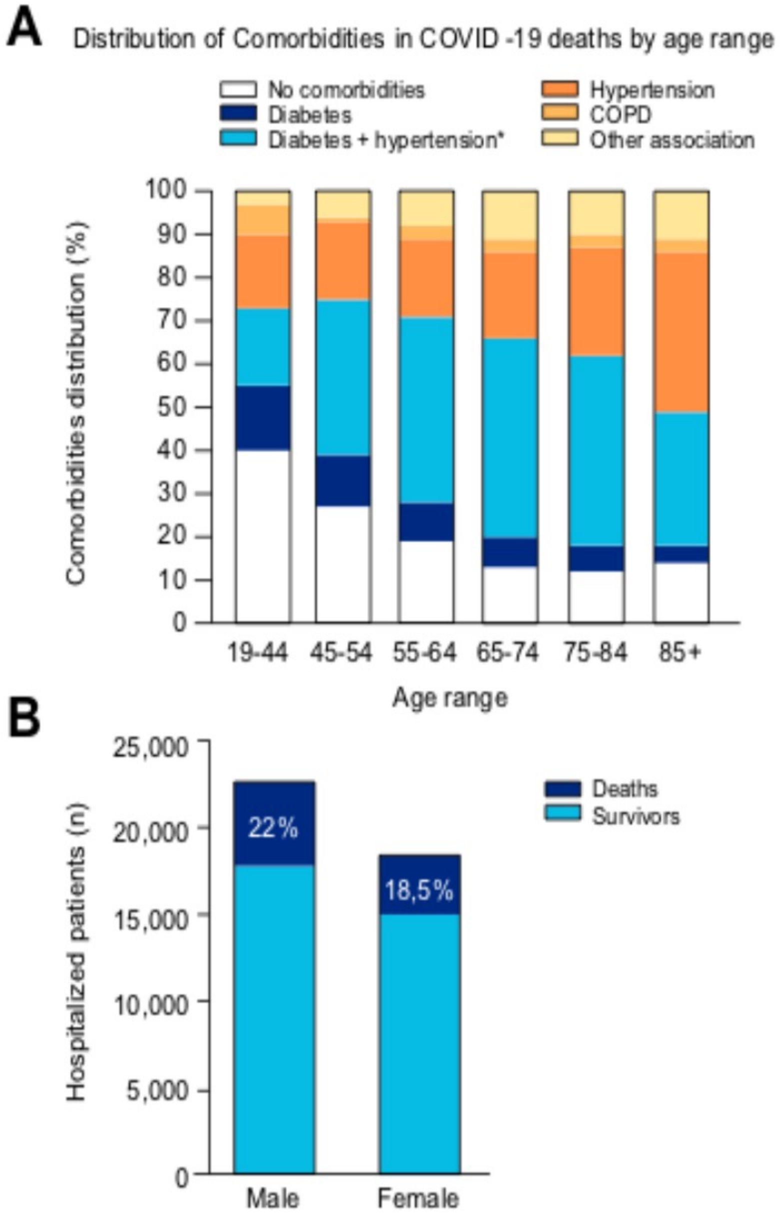


Supplementary Figure 5: A) Prevalence of comorbidities among COVID-19 related deaths. B) Distribution of death among total case of hospitalization according to gender. Data are from US populations as of April 30, 2020 and detailed in Supplementary Tables 3A and B respectively.

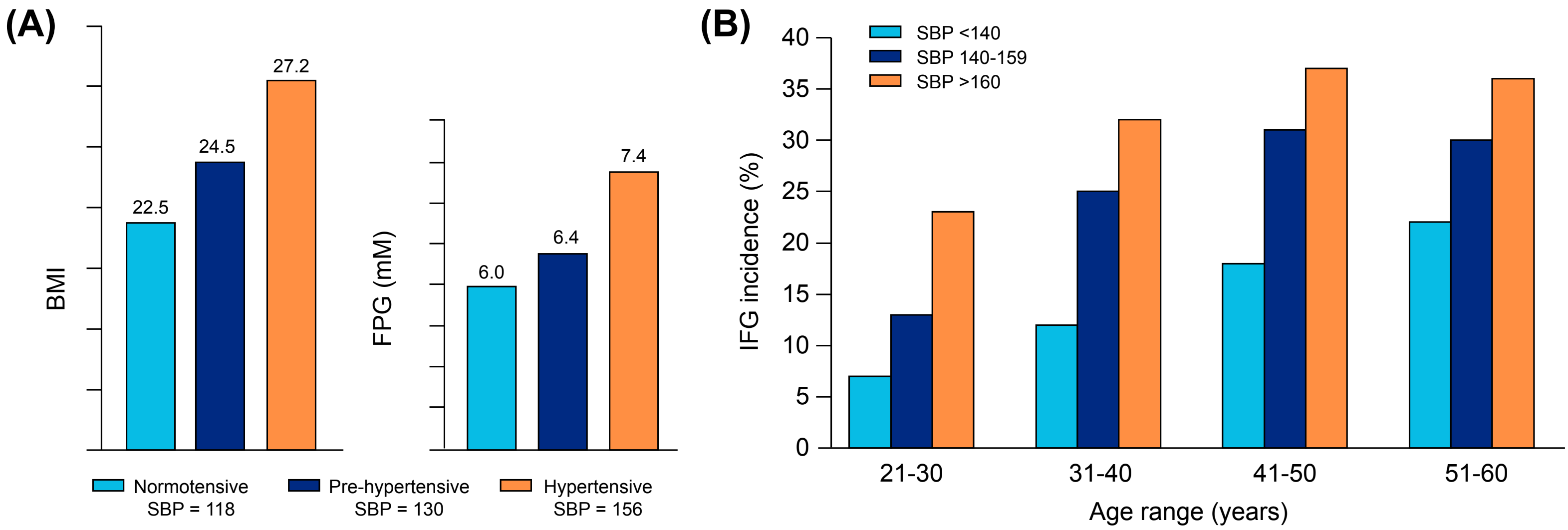


Supplementary Figure 6: Blood glucose in hypertension. (A) Correlation between hypertensive status according to SBP value (SBP=systolic blood pressure in mmHg), and BMI (top) or FPG (bottom). Adapted from Agrawal et al, 2017. (B) Prevalence of impaired fasting glucose (IFG) according to age range and SBP (mmHg). Adapted from Henry et al, 2002. (see Supplementary References)

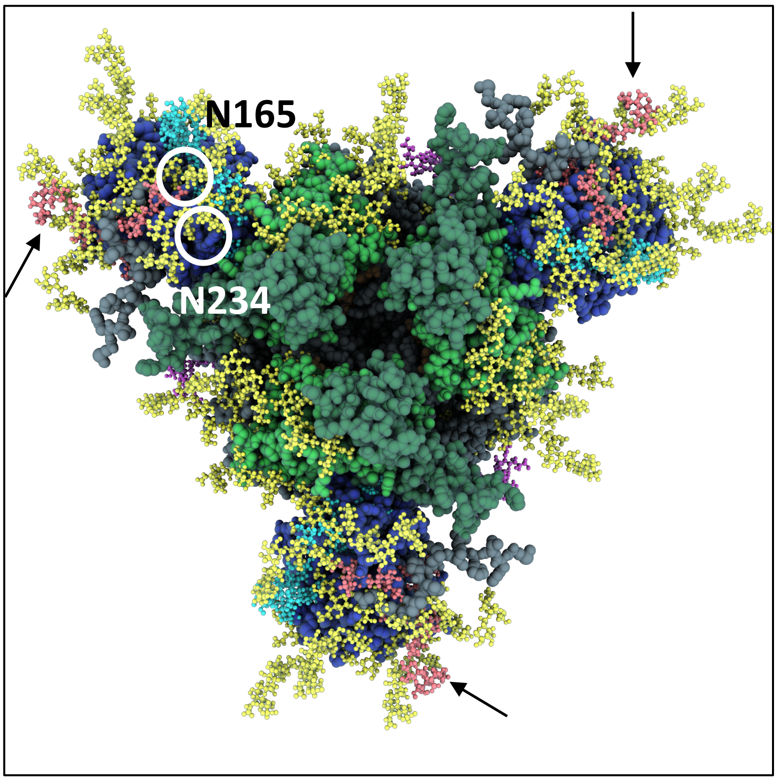


Supplementary Figure 7: Position of the potential new glycan site on the SARS-CoV-2 P.1 variant. The additionnal glycan-tree at N20 is colored in red and indicated with arrows on each S monomer of the spike. Position of N165 and N234 glycan-sites are circled on one single monomer.

A

| Age range | South Korea | Germany | South Africa | Sweden | Italy | Switzerland | Japan | Ukraine | Global |
|---|---|---|---|---|---|---|---|---|---|
| **0-9** | 0.00 | 0.04 | 0.20 | 0.04 | 0.01 | 0.02 | 0.00 | 0.00 | **0.04** |
| **10-19** | 0.00 | 0.02 | 0.30 | 0.01 | 0.01 | 0.00 | 0.00 | 0.80 | **0.14** |
| **20-29** | 0.00 | 0.12 | 1.30 | 0.16 | 0.05 | 0.00 | 0.10 | 0.30 | **0.25** |
| **30-39** | 0.40 | 0.24 | 6.10 | 0.30 | 0.22 | 0.13 | 0.31 | 3.60 | **1.41** |
| **40-49** | 0.80 | 0.79 | 12.60 | 0.73 | 0.87 | 0.29 | 1.12 | 9.40 | **3.33** |
| **50-59** | 4.59 | 3.08 | 24.40 | 2.41 | 3.40 | 1.58 | 3.26 | 19.90 | **7.83** |
| **60-69** | 11.98 | 8.47 | 26.20 | 6.44 | 9.76 | 5.98 | 9.31 | 27.70 | **13.23** |
| **70-79** | 31.74 | 20.91 | 17.70 | 20.94 | 25.25 | 20.32 | 26.29 | 22.40 | **23.19** |
| **80+** | 50.50 | 66.29 | 11.30 | 68.93 | 60.43 | 71.69 | 59.61 | 15.80 | **50.57** |
| **total** | **100.01** | **99.96** | **100.10** | **99.96** | **100.00** | **100.01** | **100.00** | **99.90** | **99.99** |

B

| Age range | South Korea | China | Spain | Italy | Switzerland | Poland | Global |
|---|---|---|---|---|---|---|---|
| **0-9** | 0.00 | 0.00 | 0.01 | 0.00 | 0.00 | 0.00 | **0.00** |
| **10-19** | 0.00 | 0.20 | 0.02 | 0.00 | 0.00 | 0.05 | **0.05** |
| **20-29** | 0.00 | 0.20 | 0.12 | 0.00 | 0.00 | 0.00 | **0.05** |
| **30-39** | 0.05 | 0.20 | 0.32 | 0.10 | 0.12 | 0.15 | **0.16** |
| **40-49** | 0.10 | 0.40 | 1.06 | 0.20 | 0.08 | 0.30 | **0.36** |
| **50-59** | 0.42 | 1.30 | 3.22 | 0.60 | 0.65 | 1.00 | **1.20** |
| **60-69** | 1.27 | 3.60 | 5.00 | 3.00 | 3.35 | 4.50 | **3.45** |
| **70-79** | 6.68 | 8.00 | 14.30 | 10.20 | 11.59 | 11.20 | **10.33** |
| **80+** | 19.45 | 14.80 | 21.10 | 20.90 | 28.45 | 20.05 | **20.79** |

**Supplementary Tables 1 (Related to Supplementary Figures 1): A)** COVID-19 mortality rate per age range as of June 2020 in eight different countries. Global rates are plotted in Supplementary Figure 1A. **B)** COVID-19 case fatality rate (CFR) per age range as of June 2020 in six different countries. Global rates are plotted in Supplementary Figure 1B. References are listed in Supplementary References.

A

| Rank | Entity | Frequency | Entity type | Rank | Entity | Frequency | Entity type |
|---|---|---|---|---|---|---|---|
| 1 | COVID-19 | 110,145 | Symptom / Disease | 51 | amino acid | 14,227 | Chemical |
| 2 | virus | 75,012 | Organism | 52 | pH | 14,107 | Chemical |
| 3 | infectious disorder | 73,574 | Symptom / Disease | 53 | gastrointestinal tract | 13,510 | Organ / System |
| 4 | coronavirus | 67,945 | Organism | 54 | immunoglobulin | 13,198 | Protein |
| 5 | human | 61,816 | Organism | 55 | antibiotic | 13,034 | Drug |
| 6 | viral | 52,801 | Organism | 56 | outbreak | 12,766 | Symptom / Disease |
| 7 | SARS-COV-2 | 49,386 | Organism | 57 | cow | 12,656 | Organism |
| 8 | SARS coronavirus | 41,463 | Organism | 58 | transfer | 12,625 | Biological Process / Pathway |
| 9 | blood | 33,970 | Organ / System | 59 | tissue | 12,542 | Organ / System |
| 10 | person | 33,893 | Organism | 60 | nucleic acids | 12,530 | Chemical |
| 11 | coronaviridae | 32,976 | Organism | 61 | ARDS | 12,392 | Symptom / Disease |
| 12 | animal | 31,998 | Organism | 62 | brain | 12,307 | Organ / System |
| 13 | lung | 31,752 | Organ / System | 63 | cardiovascular system | 11,864 | Organ / System |
| 14 | respiratory system | 31,221 | Organ / System | 64 | interferon | 11,687 | Protein |
| 15 | death | 31,114 | Symptom / Disease | 65 | skin | 11,686 | Organ / System |
| 16 | host | 27,219 | Organism | 66 | renal | 11,630 | Organ / System |
| 17 | bacteria | 26,082 | Organism | 67 | human respiratory syncytial virus | 11,356 | Organism |
| 18 | influenza | 26,000 | Symptom / Disease | 68 | T-lymphocyte | 11,189 | Cell Type |
| 19 | antibody | 24,447 | Protein | 69 | plasma | 11,132 | Organ / System |
| 20 | fever | 24,295 | Symptom / Disease | 70 | MERS coronavirus | 11,063 | Organism |
| 21 | vaccine | 24,073 | Drug | 71 | disease or disorder | 11,035 | Symptom / Disease |
| 22 | pneumonia | 23,998 | Symptom / Disease | 72 | compound | 10,907 | Chemical |
| 23 | viral infection | 23,797 | Symptom / Disease | 73 | nucleotide | 10,780 | Chemical |
| 24 | pathogen | 23,441 | Organism | 74 | neoplasm | 10,750 | Symptom / Disease |
| 25 | mouse | 23,122 | Organism | 75 | enzyme | 10,567 | Protein |
| 26 | heart | 22,583 | Organ / System | 76 | cytoplasm | 10,364 | Cell Compartment |
| 27 | DNA replication | 21,577 | Biological Process / Pathway | 77 | lower | 10,342 | Organ / System |
| 28 | pulmonary | 21,261 | Organ / System | 78 | nuclear | 10,305 | Cell Compartment |
| 29 | serum | 20,300 | Organ / System | 79 | antigen | 10,263 | Drug |
| 30 | water | 19,746 | Chemical | 80 | immunity | 10,236 | Biological Process / Pathway |
| 31 | survival | 18,915 | Biological Process / Pathway | 81 | peptidase | 10,156 | Protein |
| 32 | DNA | 16,948 | Chemical | 82 | adenoviridae | 10,148 | Organism |
| 33 | infectious disease pathway | 16,882 | Biological Process / Pathway | 83 | adenovirus infection | 10,111 | Symptom / Disease |

| | | | | | | | |
|---|---|---|---|---|---|---|---|
| 34 | membrane | 16,708 | Organ / System | 84 | hypertension | 1,094 | Symptom / Disease |
| 35 | liver | 16,688 | Organ / System | 85 | lymphocyte | 10,064 | Cell Type |
| 36 | inflammation | 16,554 | Symptom / Disease | 86 | diarrhea | 10,039 | Symptom / Disease |
| 37 | oral cavity | 16,522 | Organ / System | 87 | macrophage | 10,028 | Cell Type |
| 38 | cough | 16518 | Symptom / Disease | 88 | proliferation | 10,006 | Biological Process / Pathway |
| 39 | ribonucleic acid | 16,397 | Chemical | 89 | ACE2 | 9,933 | Protein |
| 40 | cancer | 15,767 | Symptom / Disease | 90 | neutrophil | 9,920 | Cell Type |
| 41 | human immunodeficiency virus | 15,733 | Organism | 91 | interleukin-6 | 9,883 | Protein |
| 42 | organ | 15,679 | Organ / System | 92 | nasopharyngeal | 9,851 | Symptom / Disease |
| 43 | kidney | 15,600 | Organ / System | 93 | microorganism | 9,808 | Organism |
| 44 | cytokine | 15,584 | Protein | 94 | airway | 9,703 | Organ / System |
| 45 | immune response process | 15,014 | Biological Process / Pathway | 95 | child | 9,650 | Organism |
| 46 | diabetes mellitus | 14,984 | Symptom / Disease | 96 | mammalia | 9,643 | Organism |
| 47 | molecule | 14,846 | Chemical | 97 | messenger RNA | 9,614 | Protein |
| 48 | influenza A virus | 14,686 | Organism | 98 | eye | 9,588 | Organ / System |
| 49 | SARS | 14,372 | Symptom / Disease | 99 | respiratory system disorder | 9,528 | Symptom / Disease |
| 50 | oxygen | 14,289 | Chemical | 100 | injury | 9,438 | Symptom / Disease |

B

| Rank | Entity | Rank | Entity type |
|---|---|---|---|
| 1 | water | 10 | nucleotide |
| 2 | DNA | 11 | viral RNA |
| 3 | RNA | 12 | chemical |
| 4 | molecule | 13 | lipid |
| 5 | oxygen | 14 | reagent |
| 6 | amino acid | 14 | acid |
| 7 | pH | 16 | CO2 |
| 8 | nucleic acids | 17 | glucose |
| 9 | compound | … | |

**Supplementary Tables 2 (Related to Table 1): A)** Ranking of the 100 most frequently mentioned terms in the CORD-19v47 database following COVID-19-related entity types recognition. (i.e. 'COVID-19' is mentioned in 110,145 distinct papers). **B)** Ranking of top mentioned terms from the entity type "*chemical*" in the CORD-19v47 database.

Note that in order to filter out mentions of glucose as a chemical appearing in experimental procedures, we have excluded sections entitled 'Methods' and 'Materials and Methods' from our analysis.

A

| Age range | 0-18 | 19-44 | 45-54 | 55-64 | 65-74 | 75-84 | 85+ |
|---|---|---|---|---|---|---|---|
| Total death n | 0 | 241 | 468 | 1,181 | 2,010 | 2,356 | 2,087 |
| no comorbidities n (%) | 0 | 97 (40) | 126 (27) | 225 (19) | 261 (13) | 283 (12) | 292 (14) |
| Diabetes n (%) | 0 | 36 (15) | 56 (12) | 106 (9) | 141 (7) | 141 (6) | 84 (4) |
| Diabetes + Hypertension* n (%) | 0 | 43 (18) | 169 (36) | 508 (43) | 925 (46) | 1036 (44) | 647 (31) |
| Hypertension n (%) | 0 | 41 (17) | 84 (18) | 213 (18) | 402 (20) | 589 (25) | 772 (37) |
| COPD n (%) | 0 | 17 (7) | 5 (1) | 35 (3) | 60 (3) | 71 (3) | 63 (3) |
| Other combinations n (%) | 0 | 7 (3) | 28 (6) | 94 (8) | 221 (11) | 236 (10) | 229 (11) |

B

| | Male | Female |
|---|---|---|
| Total case (n) | 22,566 | 18,441 |
| Total death (n) (%) | 4,937 (22%) | 3,406 (18,5%) |

**Supplementary Tables 3 (Related to Supplementary Figures 5). (A)** Prevalence of comorbidities among COVID-19 related deaths as plotted in Supplementary Figure 5A. (**B)** Distribution of death among total cases of hospitalization according to gender as plotted in Supplementary Figure 5B. Data from US populations as of April 30, 2020. Data source: https://ehrn.org/prevalence-of-comorbidities-in-covid-19-related-hospitalizations-and-deaths/

**A**

| | FPG (mM) | | 2h PPG (mM) | | HbA1c (%) | |
|---|---|---|---|---|---|---|
| | Control | T2DM | Control | T2DM | Control | T2DM |
| Duraisamy et al, 2010 | 4.5 | 9.01 | 5.32 | 13.05 | 5.22 | 8.97 |
| Bhowmik et al, 2018 | 4.7 | 9.5 | 5.4 | 13.9 | 5.3 | 7.6 |
| Yang et al, 2014 | 5.26 | 9.79 | 5.45 | 14.53 | 4.23 | 8.19 |
| **Mean** | **4.82** | **9.4** | **5.4** | **13.8** | **4.92** | **8.3** |

**B**

| | Control | | Hypertensive | |
|---|---|---|---|---|
| | FPG mM | *n* | FPG mM | *n* |
| Mittal, 2014 | 4.6 | *50* | 5.47 | *70* |
| Tarray et al , 2014 | 4.92 | *50* | 5.31 | *50* |
| Heianza et al , 2015 | 5.24 | *8,486* | 5.44 | *1,098* |
| **Mean** | **4.92** | | **5.41** | |

**Supplementary Tables 4 (Related to Figures 9): (A)** Values of FPG, 2h-PPG and HbA1c in control (n=2,116) versus diabetic patients (n=439) according to values reported in 3 different studies. Mean values are plotted in Figure 9A. (**B)** Values of FPG in control (n=8,586) versus hypertensive patients (n=1,218) according to values reported in three different studies. Mean values are plotted in Figure 9B. References are listed in Supplementary References.

A

| Age | FPG (mM) | | | | **Mean** |
|---|---|---|---|---|---|
| <19 | 5.2 | NA | 4.9 | 4.78 | **4.96** |
| 20-29 | 5.1 | NA | 5 | 4.83 | **4.98** |
| 30-39 | 5.05 | 4.44 | 5.1 | 4.97 | **4.89** |
| 40-49 | 5.5 | 4.61 | 5.15 | 5.22 | **5.12** |
| 50-59 | 5.7 | 4.78 | 5.2 | 5.44 | **5.28** |
| 60-69 | 5.7 | 5.06 | 5.3 | 5.50 | **5.39** |
| 70-79 | 5.71 | 5.39 | NA | 5.56 | **5.55** |
| 80-89 | 5.72 | 5.72 | NA | 5.64 | **5.69** |
| >90 | 5.72 | 6.06 | NA | NA | **5.89** |
| | Ko et al, 2006 *(n=6,901)* | Yashin et al, 2009 *(n= 5,128)* | Yates and Laing, 2002 *(n=191)* | Yi et al, 2017 *(n>10,000)* | |

B

| Age | FPG in male (mM) | | **Mean FPG male** | FPG in female (mM) | | **Mean FPG female** |
|---|---|---|---|---|---|---|
| <19 | NA | 4.78 | **4.78** | NA | 4.78 | **4.78** |
| 20-29 | NA | 4.89 | **4.89** | NA | 4.78 | **4.78** |
| 30-39 | 4.44 | 5.11 | **4.78** | 4.39 | 4.89 | **4.64** |
| 40-49 | 4.58 | 5.39 | **4.99** | 4,53 | 5.03 | **4.78** |
| 50-59 | 4.78 | 5.56 | **5.17** | 4.72 | 5.22 | **4.97** |
| 60-69 | 5.17 | 5.56 | **5.36** | 5.03 | 5.42 | **5.22** |
| 70-79 | 5.67 | 5.56 | **5.61** | 5.42 | 5.56 | **5.49** |
| 80-89 | 5.78 | 5.61 | **5.69** | 5.78 | 5.67 | **5.72** |
| >90 | 6.11 | NA | **6.11** | 6.33 | NA | **6.33** |
| | Yashin et al, 2009 *(n= 2,336)* | Yi et al, 2017 *(n>10,000)* | | Yashin et al, 2009 *(n=2,873)* | Yi et al, 2017 *(n>10,000)* | |

C

| Age | 2h PPG (mM) | | | | **Mean** |
|---|---|---|---|---|---|
| 20-29 | 6.11 | 7.22 | 7.28 | NA | **6.87** |
| 30-39 | 6.22 | 7.44 | 7.61 | 5.9 | **6.79** |
| 40-49 | 6.78 | 7.67 | 8.00 | 6.2 | **7.16** |
| 50-59 | 7.22 | 7.22 | 8.39 | 6.5 | **7.33** |
| 60-69 | 7.78 | 8.44 | 8.72 | 7.1 | **8.01** |
| 70-79 | 7.78 | 8.39 | 9.61 | 8 | **8.44** |
| 80-89 | NA | 9.22 | 10.10 | 8.4 | **9.24** |
| | Chia et al, 2018 *(n=2,777)* | Elahi et al, 1982 *(n=186)* | Elahi et al, 2000 *(n=NA)* | Shikomata et al, 1991 *(n=742)* | |

**Supplementary Tables 5 (Related to Figures 8): (A)** Values of FPG in function of age range according to data reported in 4 different studies. Mean values are plotted in Figure 8A. **(B)** Values of FPG in function of age range and gender according to data reported in 2 different studies. Mean values are plotted in Figure 8B. **(C)** Values of 2h PPG after an OGTT test in function of age range according to values reported in 4 different studies. Mean values are plotted in Figure 8C. The number of participants (n) in each study is reported. In some cases, absolute values are not reported and then directly extracted from raw plots. All references are available in the manuscript or in Supplementary References.

### Supplementary References

**References Supplementary Table 1A:** (1–10)

**References Supplementary Table 1B:** (5, 11, 12)

**References Supplementary Tables 4:** (13–18)

**References Supplementary Tables 5:** (19–25)

**References Knowledge Graphs Methods:** (26–31)

**References BioExplorer Methods:** (32–38)

**References Computational Modeling Methods:** (39–44)

1. La répartition par âge des décès liés au coronavirus ressemble à une mortalité classique - Libération, https://www.liberation.fr/direct/element/la-repartition-par-age-des-deces-lies-au-coronavirus-ressemble-a-une-mortalite-classique_112144/

2. • Sweden: coronavirus deaths by age | Statista, https://www.statista.com/statistics/1107913/number-of-coronavirus-deaths-in-sweden-by-age-groups/

3. • Spain: coronavirus mortality rate by age 2020 | Statista, https://www.statista.com/statistics/1105596/covid-19-mortality-rate-by-age-group-in-spain-march/

4. • South Korea: coronavirus death cases by age 2020 | Statista, https://www.statista.com/statistics/1105080/south-korea-coronavirus-deaths-by-age/

5. • Poland: Coronavirus (COVID-19) fatalities by age 2020 | Statista, https://www.statista.com/statistics/1110890/poland-coronavirus-covid-19-fatalities-by-age/

6. • Netherlands: coronavirus deaths by age 2021 | Statista, https://www.statista.com/statistics/1109459/coronavirus-death-casulaties-by-age-in-netherlands/

7. • Italy: coronavirus death rate by age | Statista, https://www.statista.com/statistics/1106372/coronavirus-death-rate-by-age-group-italy/

8. • COVID-19 deaths by age and gender Ukraine 2020 | Statista, https://www.statista.com/statistics/1109638/covid-19-deaths-by-age-and-gender-ukraine/

9. • Coronavirus (COVID-19) deaths in Switzerland by age group 2021 | Statista, https://www.statista.com/statistics/1110092/coronavirus-covid-19-deaths-age-group-switzerland/

10. • Coronavirus (COVID-19) deaths by gender and age Germany 2021 | Statista, https://www.statista.com/statistics/1105512/coronavirus-covid-19-deaths-by-gender-germany/

11. What we know about the victims of the coronavirus pandemic in Switzerland - The Local, https://www.thelocal.ch/20200409/what-we-know-about-the-victims-of-the-coronavirus-pandemic-in-switzerland

12. M Roser; H Ritchie; E Ortiz-Ospina; J Hasell. Coronavirus Pandemic (COVID-19), https://ourworldindata.org/coronavirus

13. Y Yang; L Gao; D-J Wang; F-R Li; Y-H Yan; X-Y Wang; X Liao; Q Huang; H Zhang; Q-R Chen; Q Wang; Y Weng; M-H Song. Comparison of 25-hydroxy vitamin D3, adiponectin and apolipoprotein A5 levels in subjects with different glucose tolerance and their correlation with blood lipid. *Biomed Res* 25, 5 (2014)

14. R Tarray; S Saleem; D Afroze; I Yousuf; A Gulnar; B Laway; S Verma. Role of insulin resistance in essential hypertension. *Cardiovascular Endocrinology & Metabolism* 3, 129–133 (2014)

15. Y Mittal. Fasting Blood Glucose Level in Patients Suffering From Hypertension. *Asian Journal of Biomedical and Pharmaceutical Sciences* 4, 4 (2014)

16. Y Heianza; Y Arase; S Kodama; SD Hsieh; H Tsuji; K Saito; S Hara; H Sone. Fasting glucose and HbA1c levels as risk factors for the development of hypertension in Japanese

individuals: Toranomon hospital health management center study 16 (TOPICS 16). *J Hum Hypertens* 29, 254–259 (2015)

23. D Elahi; D Muller. Carbohydrate metabolism in the elderly. *Eur J Clin Nutr* 54, S112–S120 (2000)

24. D Elahi; DC Muller; SP Tzankoff; R Andres; JD Tobin. Effect of Age and Obesity on Fasting Levels of Glucose, Insulin, Glucagon, and Growth Hormone in Man. *Journal of Gerontology* 37, 385–391 (1982)

25. H Shimokata; DC Muller; JL Fleg; J Sorkin; AW Ziemba; R Andres. Age as independent determinant of glucose tolerance. *Diabetes* 40, 44–51 (1991)

26. JY Yen. Finding the K Shortest Loopless Paths in a Network. *Management Science* 17, 712–716 (1971)

27. M Neumann; D King; I Beltagy; W Ammar. ScispaCy: Fast and Robust Models for Biomedical Natural Language Processing. *Proceedings of the 18th BioNLP Workshop and Shared Task* 319–327 (2019)

28. J Lee; W Yoon; S Kim; D Kim; S Kim; CH So; J Kang. BioBERT: a pre-trained biomedical language representation model for biomedical text mining. *Bioinformatics* btz682 (2019)

29. G Fragoso; S de Coronado; M Haber; F Hartel; L Wright. Overview and utilization of the NCI thesaurus. *Comp Funct Genomics* 5, 648–654 (2004)

30. L Danon; J Duch; A Diaz-Guilera; A Arenas. Comparing community structure identification. *J Stat Mech* 2005, P09008–P09008 (2005)

31. G Bouma. Normalized (Pointwise) Mutual Information in Collocation Extraction. *Proceedings of the Biennial GSCL Conference 2009* (2009)

32. R Yan; Y Zhang; Y Li; L Xia; Y Guo; Q Zhou. Structural basis for the recognition of SARS-CoV-2 by full-length human ACE2. *Science* 367, 1444–1448 (2020)

33. A Watson; MJS Phipps; HW Clark; C-K Skylaris; J Madsen. Surfactant Proteins A and D: Trimerized Innate Immunity Proteins with an Affinity for Viral Fusion Proteins. *J Innate Immun* 11, 13–28 (2019)

34. Y Watanabe; JD Allen; D Wrapp; JS McLellan; M Crispin. Site-specific analysis of the SARS-CoV-2 glycan shield. Microbiology (2020)

35. A Shajahan; S Archer-Hartmann; NT Supekar; AS Gleinich; C Heiss; P Azadi. Comprehensive characterization of N- and O- glycosylation of SARS-CoV-2 human receptor angiotensin converting enzyme 2. Biochemistry (2020)
36. BW Neuman; BD Adair; C Yoshioka; JD Quispe; G Orca; P Kuhn; RA Milligan; M Yeager; MJ Buchmeier. Supramolecular Architecture of Severe Acute Respiratory Syndrome Coronavirus Revealed by Electron Cryomicroscopy. *JVI* 80, 7918–7928 (2006)
37. I-N Hsieh; X De Luna; MR White; KL Hartshorn. The Role and Molecular Mechanism of Action of Surfactant Protein D in Innate Host Defense Against Influenza A Virus. *Front Immunol* 9, 1368 (2018)
38. YM Bar-On; A Flamholz; R Phillips; R Milo. SARS-CoV-2 (COVID-19) by the numbers. *eLife* 9, e57309 (2020)
39. FP Schwarz; KD Puri; RG Bhat; A Surolia. Thermodynamics of monosaccharide binding to concanavalin A, pea (Pisum sativum) lectin, and lentil (Lens culinaris) lectin. *Journal of Biological Chemistry* 268, 7668–7677 (1993)
40. A Kussrow; E Kaltgrad; ML Wolfenden; MJ Cloninger; MG Finn; DJ Bornhop. Measurement of Monovalent and Polyvalent Carbohydrate−Lectin Binding by Back-Scattering Interferometry. *Anal Chem* 81, 4889–4897 (2009)
41. R Jolivet; JS Coggan; I Allaman; PJ Magistretti. Multi-timescale Modeling of Activity-Dependent Metabolic Coupling in the Neuron-Glia-Vasculature Ensemble. *PLoS Comput Biol* 11, e1004036 (2015)
42. MQ Gaunsbaek; KJ Rasmussen; MF Beers; EN Atochina-Vasserman; S Hansen. Lung surfactant protein D (SP-D) response and regulation during acute and chronic lung injury. *Lung* 191, 295–303 (2013)
43. Y Contreras-Baeza; PY Sandoval; R Alarcón; A Galaz; F Cortés-Molina; K Alegría; F Baeza-Lehnert; R Arce-Molina; A Guequén; CA Flores; A San Martín; LF Barros.

Monocarboxylate transporter 4 (MCT4) is a high affinity transporter capable of exporting lactate in high-lactate microenvironments. *Journal of Biological Chemistry* 294, 20135–20147 (2019)

44. S Anand. Size distribution of virus laden droplets from expiratory ejecta of infected subjects. *Scientific Reports* 9 (2020)
